# A Comprehensive Guide to MEGA-PRESS for GABA Measurement

**DOI:** 10.1101/2021.11.24.21266827

**Authors:** AL Peek, TJ Rebbeck, AM Leaver, NA Puts, SL Foster, KM Refshauge, G Oeltzschner, MRS Expert Panel

## Abstract

**Background:** The aim of this guideline is to provide a series of evidence-based recommendations that allow those new to the field of MEGA-PRESS to produce high-quality data for the measurement of GABA levels using edited magnetic resonance spectroscopy with the MEGA-PRESS sequence at 3T. GABA is the main inhibitory neurotransmitter of the central nervous system and has been increasingly studied due to its relevance in many clinical disorders of the central nervous system. MEGA-PRESS is the most widely used method for quantification of GABA at 3T, but is technically challenging and operates at a low signal-to-noise ratio. Therefore, the acquisition of high-quality MRS data relies on avoiding numerous pitfalls and observing important caveats.

**Methods:** The guideline was developed by a working party that consisted of experts in MRS and experts in guideline development and implementation, together with key stakeholders. Strictly following a translational framework, we first identified evidence using a systematically conducted scoping literature review, then synthesised and graded the quality of evidence that formed recommendations. These recommendations were then sent to a panel of 21 world leaders in MRS for feedback and approval using a modified-Delphi process across two rounds.

**Results:** The final guideline consists of 23 recommendations across six domains essential for GABA MRS acquisition (Parameters, Practicalities, Data acquisition, Confounders, Quality/reporting, Post-processing). Overall, 78% of recommendations were formed from high-quality evidence, and 91% received agreement from over 80% of the expert panel.

**Conclusion:** These 23 expert-reviewed recommendations and accompanying extended documentation form a readily usable guideline to allow those new to the field of MEGA-PRESS to design appropriate MEGA-PRESS study protocols and generate high-quality data.

## 1. INTRODUCTION

Gamma-aminobutyric acid (GABA) is the main inhibitory neurotransmitter of the central nervous system (CNS) and plays an important role in regulating healthy brain function. For example, GABA is implicated in sensory processing (Puts *et al*., 2017; Wood *et al*., 2021), learning (Kolasinski *et al*., 2019; Zacharopoulos *et al*., 2021), memory (Gasbarri and Pompili, 2014) and motor function (Kolasinski *et al*., 2019; Zacharopoulos *et al*., 2021).

GABA is of particular interest in clinical conditions of the CNS and altered GABAergic function has been associated with chronic pain (Peek *et al*., 2020), psychological disorders e.g. stress and depression (Schür *et al*., 2016; Godfrey *et al*., 2018), substance addiction (Vengeliene *et al*., 2008) and neurodevelopmental disorders, e.g. autism spectrum disorder (Marotta *et al*., 2020). Evidence for altered GABA function comes through multiple lines of enquiry including animal models (Enna and McCarson, 2006; Sun *et al*., 2018), genetics (Baulac *et al*., 2001; Coghlan *et al*., 2012), post-mortem studies (de Jonge *et al*., 2017), blood plasma (Petty, 1994; Bhandage *et al*., 2019) and *in-vivo* Magnetic Resonance Spectroscopy (MRS) (Schur *et al*., 2016; Peek *et al*., 2020). Given the wealth of evidence, targeting the GABAergic system with therapeutic interventions may therefore prove fundamental to improving patient outcomes in these conditions. However, this requires a better understanding of the role of GABA in humans, which requires the reliable measurement of GABA in the human brain. The only currently available approach to measure GABA *in-vivo* in humans is through tailored MRS.

MRS is a non-invasive brain imaging technique which enables the *in-vivo* quantification of endogenous brain neurometabolites based upon their chemical structure. Conventional proton MRS has been successfully used to quantify numerous neurometabolites, such as glutamate, N-acetylaspartate (NAA) and choline-containing compounds. GABA is also present in the MR spectrum, however, due to its lower concentration and complicated peak pattern, its signal is difficult to reliably separate from more abundant neurometabolites such as creatine (Mullins *et al*., 2014)-particularly at field strengths typical for current clinical MRI scanners. The most widely used technique for measuring GABA levels at 3T is J-difference editing, most famously implemented in the MEscher–GArwood Point RESolved Spectroscopy (MEGA-PRESS) experiment (Mescher *et al*., 1998). MEGA-PRESS consists of two sub-experiments (usually acquired in an interleaved fashion), one applying editing pulses at a frequency of 1.9 ppm to selectively refocus the coupling evolution of the GABA signal at 3 ppm (‘Edit-ON’), while the other allows the free evolution of the spin system throughout the echo time (‘Edit-OFF’). Subtracting the Edit-OFF from the Edit-ON spectrum reveals a difference-edited GABA signal while removing the stronger overlapping signals from creatine-containing compounds. (see de Graaf 2019 for review). The edited signal at 3 ppm is contaminated by co-edited macromolecular signals (estimated to account for about 50% of the edited signal area) and is commonly referred to as GABA+. While the macromolecular contamination can be reduced by adding a second editing pulse at 1.5 ppm (Henry *et al*., 2001), the increased specificity comes at the expense of a much greater sensitivity to experimental instability, particularly thermal drift of the magnetic field strength (Edden *et al*., 2016).

The separability of the GABA signal is significantly improved using MEGA-PRESS, but accurate detection and quantification still require high-quality data. Data quality is determined to a great extent by the choice of acquisition parameters, however, few studies provide sufficient detail of these. For example, in a recent meta-analysis (Peek *et al*., 2020) investigating the use of MRS to measure GABA levels in pain conditions, only two out of fourteen studies reported using parameters that were deemed adequate for quantification of GABA levels. The remaining studies either documented using inadequate parameters or sequences, or altogether failed to fully report the parameters used, a finding resonated in other reviews such as Schur *et al*. 2016. The heterogeneity in MRS acquisition parameters used within the field has been acknowledged as a significant barrier to the reproducibility and comparability of quantitative MRS outcome measures (Mullins *et al*., 2014; Peek *et al*., 2020). In response, multiple expert panels have recently formed to establish consensus guidelines for minimal best practice in acquisition and analysis of MRS data (Mullins *et al*., 2014; Öz *et al*., 2020; Choi *et al*., 2021; Kreis *et al*., 2021). While some aspects covered in these consensus guidelines might apply to GABA measurement using MEGA-PRESS, the specific requirements for its successful application are not addressed in detail.

A further barrier to implementing these consensus documents is that they are typically written by experts with a high level of technical knowledge, leading to some recommendations being difficult for those new to the field to interpret and adequately implement. The growing field of translational research has increasingly seen those from fields outside of magnetic imaging physics wishing to use advanced MRS methods in both clinical and research populations.

Examples include clinician-researchers and higher degree research students in areas such as pain medicine, physiotherapy and psychology. Typically these researchers do not have a background in magnetic resonance physics, and often do not have direct access to the resources or expertise required to interpret and implement technical consensus documents. We have therefore identified a need for an easily accessible and translatable guideline to the adequate use of MEGA-PRESS for the measurement of GABA. However, the substantial heterogeneity in preferred acquisition parameters, even among leading MRS experts, is a challenge for creating widely applicable methodological guidelines.

We therefore used an established translational framework widely used for developing clinical guidelines in order to maximize the objectivity of our recommendations. The National Health and Medical Research Council (NHMRC), the leading governmental authority on medical research in Australia, recommends a multi-stage process for guideline development (NHMRC, 2021). Four key aspects to ensure robustness include: 1) engaging subject, and methodological expertise alongside end-users, 2) evidence synthesis, 3) establishing quality and strength of evidence using the Grading of recommendations, Assessment, Development and Evaluation (GRADE) (NHMRC, 2009), and 4) independent expert review of the recommendations (NHMRC, 1999, 2021). These steps ensure guidelines are credible, useable and ready for implementation into practice.

The result of this study is a robust, translatable, evidence-based, and expert-reviewed guideline that will enable those new to the field to use MEGA-PRESS to acquire high-quality data for the reliable quantification of brain GABA levels. The adherence to a translational framework ensures that the guidelines are evidence-based, rather than a narrative of personal opinions and experiences. Whilst the guideline has been written specifically for the reliable measurement of GABA using MEGA-PRESS at 3T, many of the recommendations will, with certain modifications, also be applicable to similar techniques employing different signal localization (e.g. MEGA-sLASER, MEGA-SPECIAL (Edden *et al*., 2012)), editing schemes (HERMES) (Saleh *et al*., 2016b), and target metabolites (Harris *et al*., 2017).

## 2. METHODS

We followed the NHMRC framework Guidelines for Guidelines (NHMRC, 2021) and utilized the ADAPTE toolkit (The ADAPTE Collaboration, 2009) to develop this guideline. This framework divides the evidence synthesis and recommendation formation workflow into three stages: set up, adaptation and finalisation (The ADAPTE Collaboration, 2009). The stages are summarised in Figure 1.

**Figure 1:**
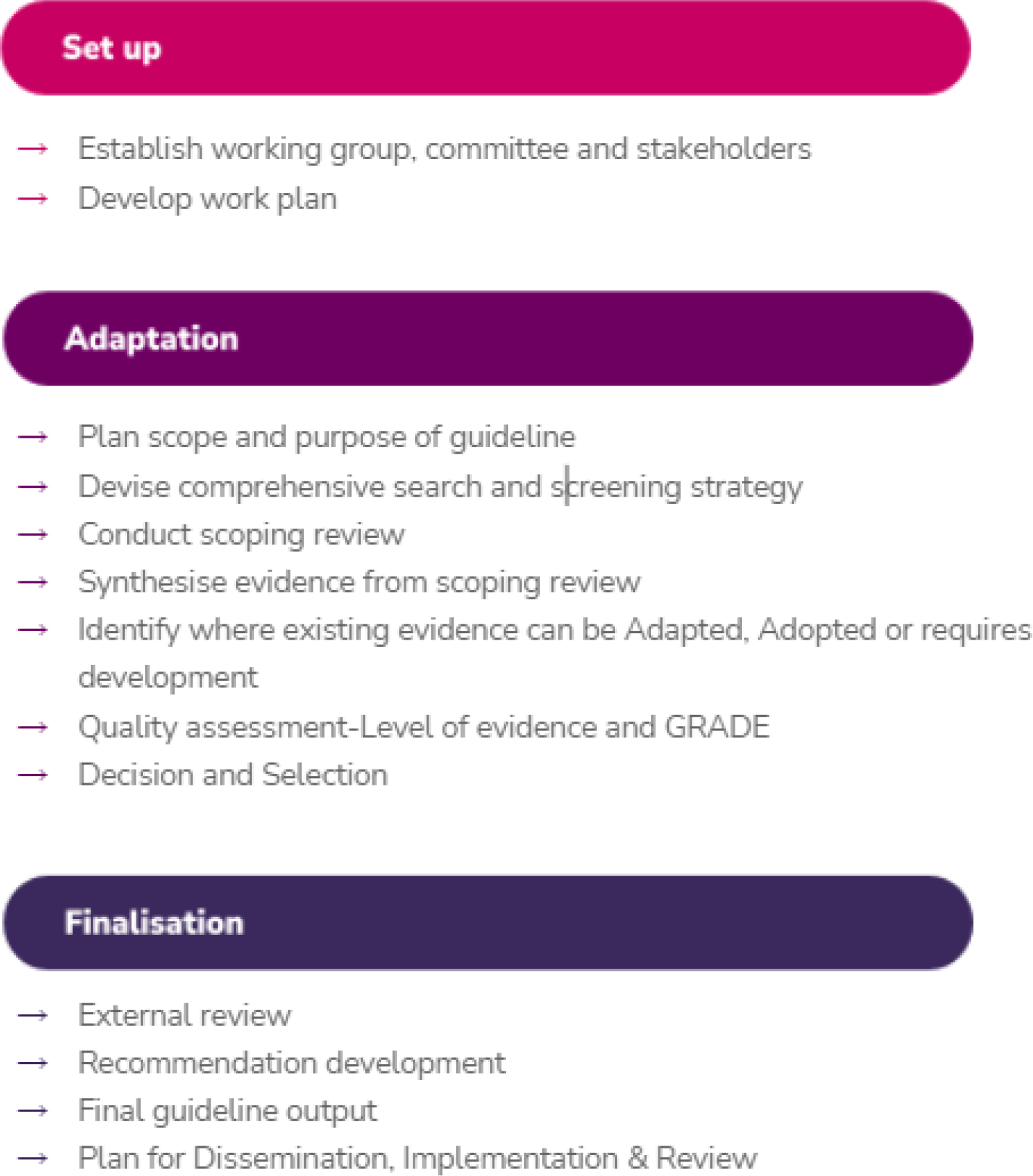
Demonstrating the process followed to develop the guideline based on the ADAPTE process (The ADAPTE Collaboration, 2009)

The purpose of the set up stage was to establish the guideline working party and sub-committees, identify key stakeholders and formulate a work plan (The ADAPTE Collaboration, 2009).

### 2.1.1 Committee establishment and stakeholder engagement

The guideline working party included a core team of six co-authors. The working party consisted of two sub-committees; i) Guideline development/implementation sub-committee (four members with a total of over 40 years of experience in forming clinical/therapeutic guidelines) and ii) MRS sub-committee (three members with a total of over 23 years experience in MRS of GABA). One author was included in both sub-committees to ensure consistency, communication and continuity across meetings. Key stakeholders reflect proposed end-users and those with an interest in the final guideline. Stakeholders were identified and engaged by the working party to be involved in the development process. The key stakeholders were a research radiographer, a PhD student studying MRS, three clinician-researchers who were investigating GABA levels in multiple pain conditions, and two MRS experts who provide training to new MRS users.

### 2.1.2 Work plan

A work plan identifying and recruiting all expertise required for project completion through large international collaborative networks was developed. Stages were identified through NHMRC Guidelines for Guidelines (NHMRC, 2021) and a time-line established. Details of the Adaptation and Finalisation stages are described as follows.

### 2.2 Adaptation

The adaptation stage was the largest of stages and included several steps from systematically identifying literature through a scoping review, through to the formulation of guideline recommendations.

#### 2.2.1 Scope and purpose

The working party met with key stakeholders on two occasions through an iterative discussion process to arrive at the scope and purpose of the Comprehensive Guide to MEGA-PRESS for GABA measurement. The result of the discussions led to the identification of six key domains critical for high-quality data: Parameters, Practicalities, Data Acquisition, Confounders, Quality/Reporting, Post-processing. The working party and stakeholders agreed the following were not within scope: i) providing in-depth review of all differences between vendor-specific user interfaces, hardware, and implementations of the MEGA-PRESS sequence; ii) details regarding post-processing, modelling and quantification methods, except for those aspects with direct implications for the acquisition protocol design for example, the necessity of acquiring a water-reference signal (see 4. *Discussion*). Further it was agreed that the focus would be set on recommendations for measuring GABA at 3T in clinical and research populations using MEGA-PRESS, although some recommendations would translate to other metabolites, field strengths and sequences.

#### 2.2.2 Search and Screening

Evidence to inform the guideline was identified through a systematically conducted scoping review. A search strategy was developed using terms for GABA editing (e.g. MEGA-PRESS, spectral editing, GABA) AND magnetic resonance spectroscopy (e.g. MRS, magnetic resonance spect*) AND terms specific to GABA MRS acquisition stages (e.g. gradients, shim). Three databases were searched (Ovid MEDLINE, Embase and PubMed) and reference lists of included studies were screened by the MRS expert sub-committee for any missing publications. A two-stage approach was used to screen studies for inclusion against the pre-specified inclusion criteria regarding study methods and study design (For further details of review methodology see Supplement 1). Studies were included if they used methods involving single-voxel MRS data acquired in humans, phantoms or using computer simulations. Study designs were included if they were consensus documents, systematic reviews, randomised controlled trials or methodological investigations. Studies were excluded if the methods included animals, used multi-voxel or spectroscopic imaging techniques (beyond the scope of these guidelines), or used designs that were narrative (non-evidence-based) reviews, commentaries or conference proceedings. In the first stage of screening, two reviewers independently screened titles and abstracts to identify studies appropriate for full text review (AP, GO). In the second stage, full texts were screened for inclusion. Data were then extracted independently by two authors using a standardised form for each of the six pre-identified domains. Inconsistencies in screening and disagreement on exclusion/inclusion were discussed and resolved with a third reviewer (NP). The MRS sub-committee reviewed the results of the search and identified any key missing papers.

#### 2.2.3 Results of the scoping review

The initial search retrieved 2664 studies, 21 additional publications were identified following the reference list search of included publications, the MRS-subcommittee review, and following the release of a special issue of *NMR in Biomedicine.* The special issue *“Advanced methodology for in vivo magnetic resonance imaging”* (Choi *et al*., 2021) contained a series of expert consensus guidelines in MRS published after the commencement of the search.

Following removal of duplicates, 1460 studies were screened against the inclusion and exclusion criteria, resulting in exclusion of a further 1283 records, leaving 176 records for full-text screening. Following the exclusion of 87 studies (32 due to study design, 39 due to content e.g. not MEGA-PRESS, or not relevant to 3T, and 16 for both content and design reasons), 90 publications were used to inform the guidelines (For PRISMA Flowchart see Supplement 2). Nine of the 90 publications were consensus documents, one randomized control trial, one seminal textbook describing the theory of *in-vivo* MR spectroscopy, one seminal paper documenting MEGA-PRESS practices, four systematic reviews, three multi-site trials, and seventy-one methodological publications. The publications used to inform each recommendation are listed in Supplement 3.

#### 2.2.4 Evidence Synthesis

The MRS sub-committee summarised evidence from the studies identified by the scoping review under the six pre-identified domains. The MRS sub-committee used an iterative process to establish where recommendations currently existed in consensus documents, and could be later considered for adoption or adaptation or where recommendations would require development. Following the ADAPTE framework for guideline adaptation (The ADAPTE Collaboration, 2009), a recommendation is considered suitable for *Adoption-* when it can be lifted directly from an existing guideline or for *Adaption-* when the recommendation needs to be adjusted to suit the audience or context. Where no evidence exists the recommendations require development *DeNovo* (‘from scratch’) (NHMRC, 2021). This first scoping draft (Draft 1) included 20 recommendations under the six domains. Furthermore, the MRS sub-committee identified five areas that required recommendation development.

#### 2.2.5 Evidence Level Assessment and GRADING the certainty of evidence

An NHMRC *Level of Evidence* was assigned to each study included in the evidence synthesis for each of the recommendations. The Level of Evidence describes the suitability of a study design to address a research question (ranging from Level 1 indicating the most robust design to Level 4 indicating the least robust design) (NHMRC, 2009). Studies involving confounders of GABA levels were assessed using the traditional hierarchy of evidence (NHMRC, 2009) given that such research questions are best answered through systematic reviews of randomised controlled trials (Level 1). For studies reporting MRS principles and acquisition parameters, the MRS sub-committee considered consensus documents the highest level of evidence (Level 1). Hence, to appraise these publications, the traditional NHMRC evidence hierarchy was adapted following the recommendations for hierarchy modification by the NHMRC (NHMRC, 2009). Details of the traditional and modified evidence hierarchy are detailed in Table 1.

**Table 1:**
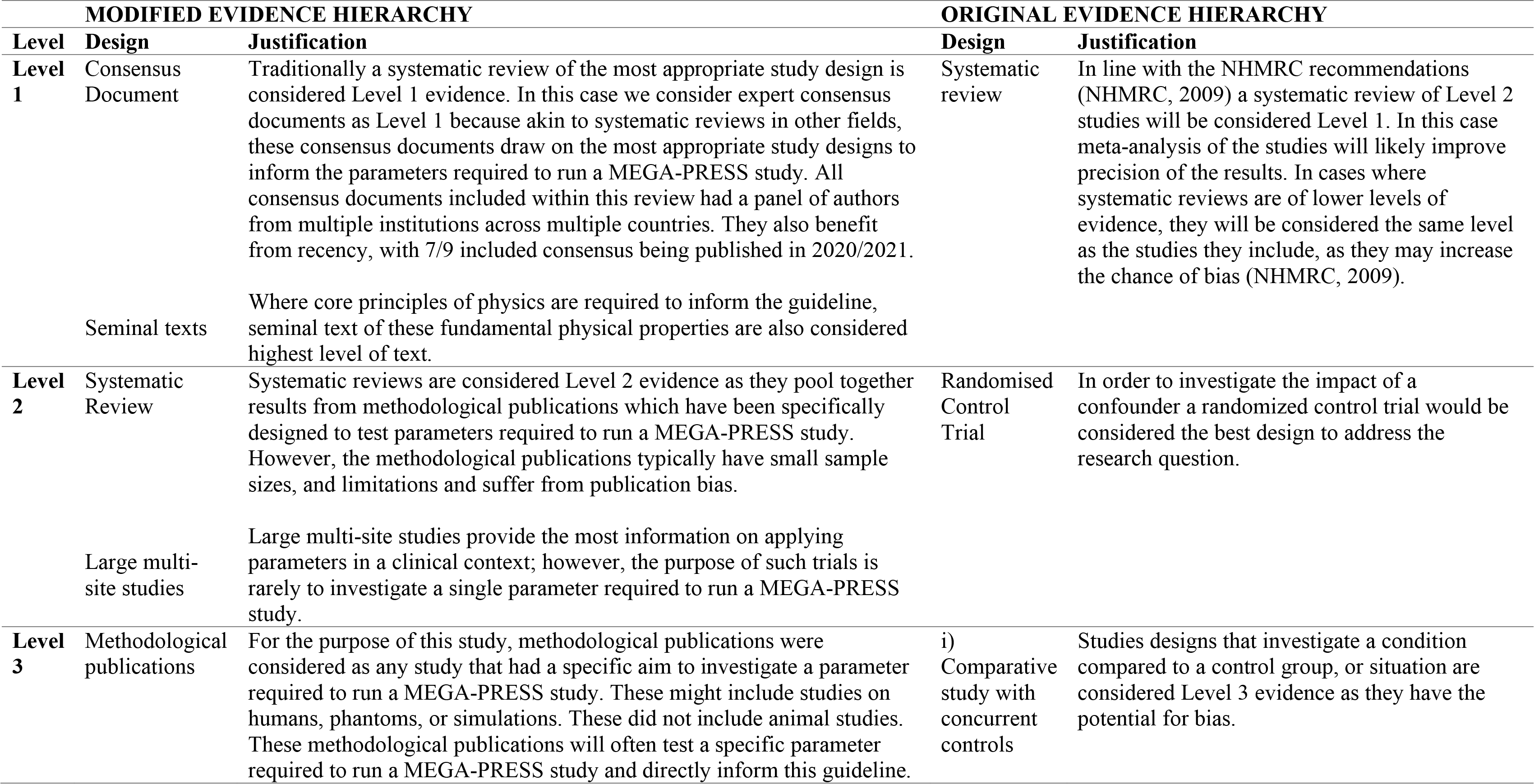
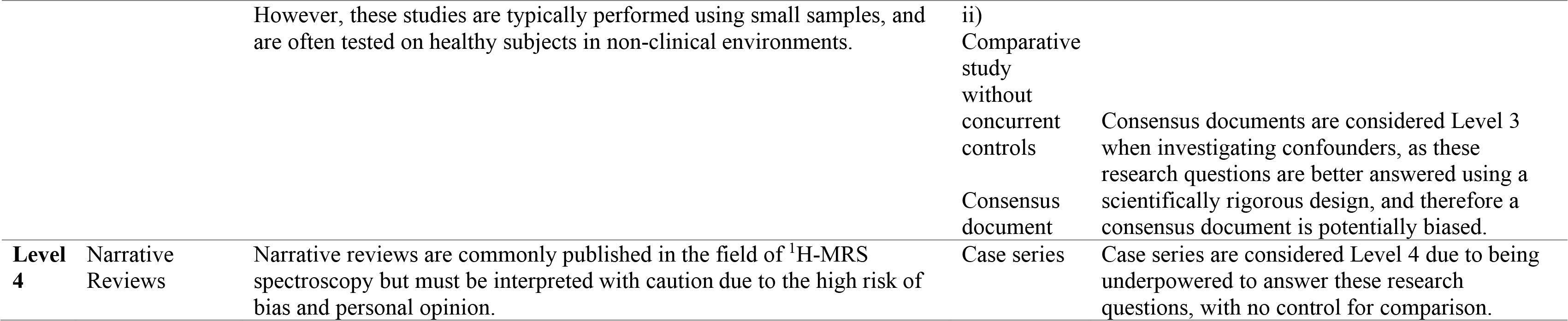
Level of evidence modified from NHMRC (2009) (NHMRC, 1999)

The modified Grading of recommendations, Assessment, Development and Evaluation (GRADE) (NHMRC, 2009) was then utilized to determine the degree of certainty in the body of evidence used to inform each of the recommendations. The GRADE process considers the Level of Evidence and direction of findings to determine the level of confidence that can be placed in the recommendation (NHMRC, 2009). The modified-GRADE ranges from GRADE A where a recommendation is informed by a number of Level 1 studies providing consistent recommendations through to GRADE I where there is insufficient evidence to provide a recommendation (Table 2). The GRADE process was carried out independently by four blinded member of the development/ implementation sub-committee. Disagreements were resolved through discussion.

**Table 2.**
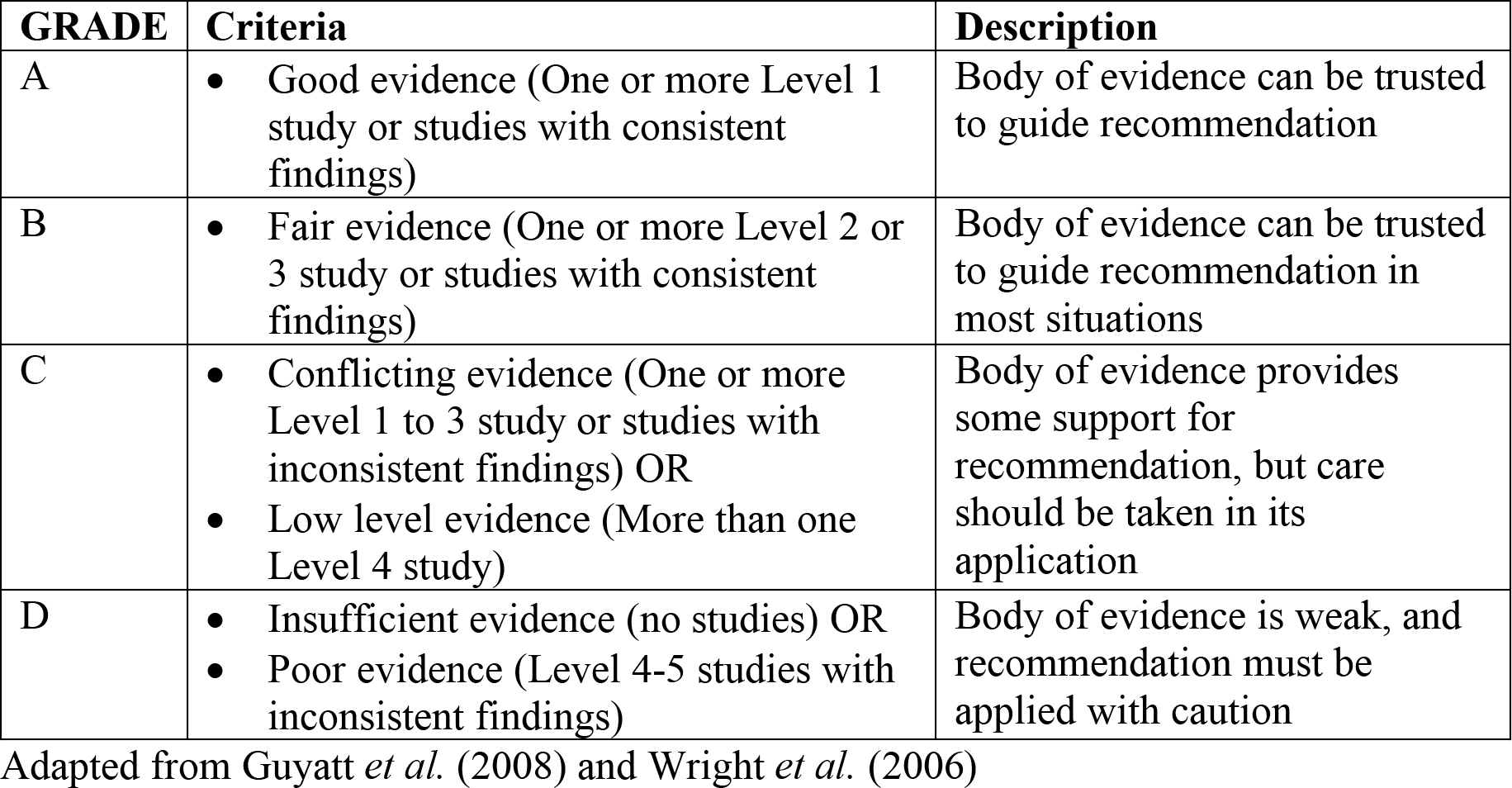
GRADE of Recommendation.

#### 2.2.6 Decision and Selection

The first draft (Draft 1) consisting of 20 recommendations was circulated to the key stakeholders prior to an in-person consensus meeting held at the 5^th^ International GABA Symposium (19^th^-21^st^ November 2019, Park City, UT, United States). The aims of the consensus meeting were: 1) to discuss key information required by those new to the field of MRS, and identify any gaps not addressed through the draft recommendations; 2) to identify and reach agreement where recommendations could be adopted or adapted from existing evidence; 3) to determine the process to develop recommendations for areas currently not supported by evidence.

As a result of the stakeholder meeting, the 20 recommendations were revised and augmented to 26 recommendations. Agreement was reached that 12 recommendations were suitable for direct adoption, nine for adaptation, and five required development (note: four of these five were later adapted from recommendations in newly released consensus documents). It was agreed that the process for development would be led by the MRS sub-committee. The MRS sub-committee would use and customise evidence from other fields or sequences for GABA MEGA-PRESS. Discussion regarding the key information required for those new to the field was agreed upon.

These decisions were then forwarded to the development/implementation sub-committee. This sub-committee wrote recommendations in easily understandable language suitable for those new to the field of MRS. The revised draft was then circulated back to the stakeholders and a finalised draft (Draft 2) was prepared to be circulated for review by an external expert panel.

### 2.3 Finalisation

The finalisation stage included; external expert review, production of this peer review publication, one-page infographic and extended guideline, and agreeing upon the implementation and dissemination plan and schedule for review and update (The ADAPTE Collaboration, 2009).

#### 2.3.1 External review

The finalised draft (Draft 2) was sent for agreement and review by a panel of experts using a modified-Delphi process. The modified-Delphi process is a group consensus strategy, designed to transform opinions into group consensus using an iterative multi-stage process (Hasson *et al*., 2000; Miller *et al*., 2020). The expert panel was established through invitation by the MRS sub-committee. Experts were identified based on their contribution to recent MRS consensus documents, and their contribution to the field of MRS. The panel consisted of 21 expert MRS researchers from 15 universities in eight different countries. In Round 1, experts rated a) their agreement with the content of the recommendation, and b) the suitability of the recommendation for use in a beginner’s guide. Ratings were on a Likert scale of -5 to +5 (where -5 to -1 indicated disagreement, 0 represented a neutral agreement, and 1 to 5 indicated agreement). Experts were also given the opportunity to comment on each of the recommendations and submit suggestions for modifications. The results from Round 1 and 2 expert panel agreement were analysed using percentages.

Recommendations were classified as having ‘expert panel endorsement’ and accepted into the final guideline where at least 80% of the expert panel had agreed to the recommendation. In cases where recommendations did not reach the 80% threshold, they were revised, taking into account the written feedback from the expert panel. These revised recommendations were then re-sent to the expert panel for a second rating (Round 2). The Round 2 expert panel consisted of 20 experts, as one expert was unavailable to review the revised recommendations. Any recommendation not achieving agreement of at least 80% of the expert panel in Round 2 was not given the ‘expert panel endorsement’ label. In these instances, evidence was reviewed by the working party, and the significance of removing the recommendation from the guideline was deliberated until a final verdict on the recommendation was reached.

#### 2.3.2 Recommendation development

The finalised Comprehensive Guide to MEGA-PRESS for GABA Measurement consists of 23 recommendations across the six key domains. Nineteen of the 26 recommendations sent for expert panel review (Draft 2) received expert panel endorsement (over 80% agreement) in Round 1 (Figure 2). Sixteen of these were immediately accepted into the guideline. Three of the nineteen required further refinement (1 due to new evidence being published, one due to not being deemed suitable for those new to the field by the expert panel, one due to being deemed too long by the expert panel). Following expert feedback from Round 1, the recommendations were consolidated and re-grouped from 26 to 23 recommendations.

**Figure 2.**
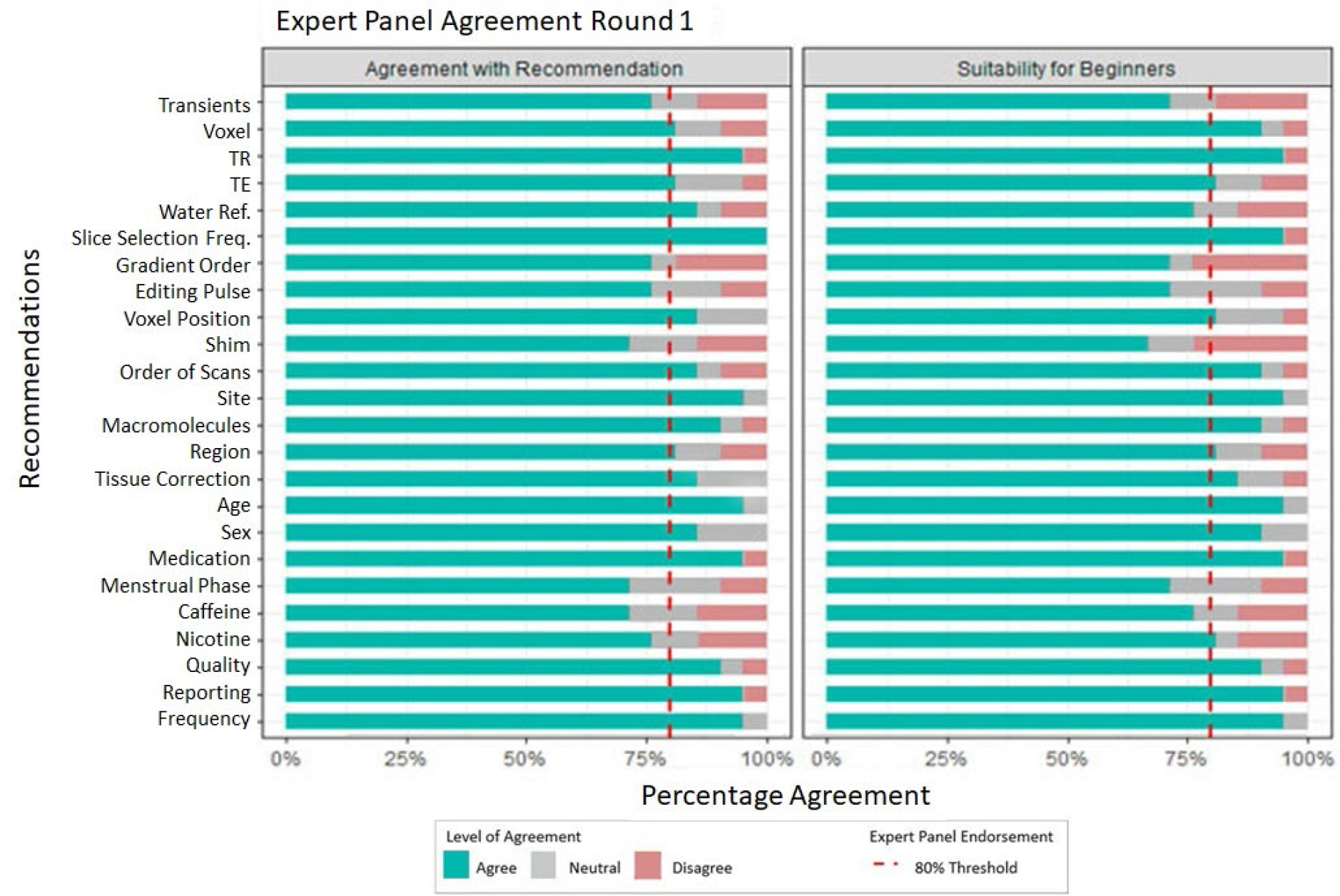
Results from Round 1 of the Expert panel review

Overall, eight recommendations were revised and submitted to the expert panel for Round 2 assessment. Following Round 2, a further six recommendations received expert panel endorsement and were accepted into the guideline (Figure 3). Two recommendations did not receive expert endorsement (‘gradient order’ -75% and ‘water reference’ -55%). In these cases, the MRS sub-committee revisited the evidence for these recommendations and debated the inclusion of these recommendations in the guideline. In both cases the result of the debate was to include the recommendation, without expert panel endorsement, with the addition of further explanatory notes in the consideration section of the extended document.

**Figure 3.**
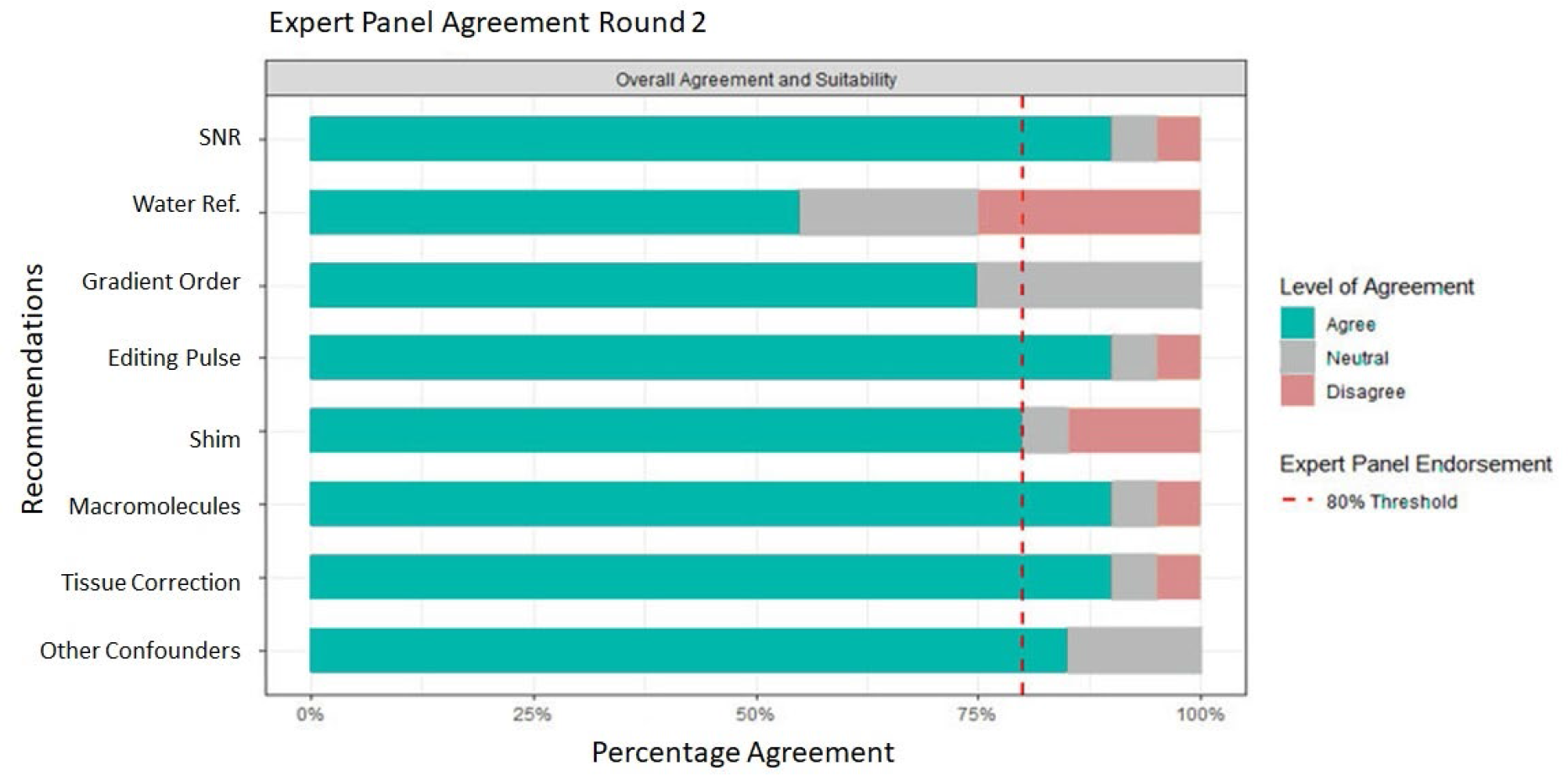
Results from Round 2 of the Expert panel review

#### 2.3.3 Final Guideline outputs

The three outputs from this work include a full guideline (Supplement 4), this peer reviewed publication summarising the recommendations and a one-page infographic summary (Supplement 5).

1. A Comprehensive Guide to MEGA-PRESS for GABA measurement (Full guideline) The full guideline (Supplement 4) is a detailed document providing background information on the subject of each recommendation. The full-length guideline is recommended for consultation when upskilling in the field of MEGA-PRESS, particularly during the study protocol design phase. Each final recommendation included in the guideline is the result of the evidence synthesis and the expert panel feedback. Therefore this guideline consists of the full evidence summary that informed the recommendation, and includes the key considerations added by the expert panel that resulted in the final recommendation.
2. The peer reviewed publication (This manuscript) This peer reviewed publication first outlines the rigor of the methodological process of recommendation development and then provides a summary of the recommendations. This manuscript provides GRADE of evidence, percentage of expert panel agreement and a shortened summary of the evidence synthesis and expert panel feedback that informed the recommendation. This manuscript can be used instead of the full-length guideline when a brief overview of parameters that determine data quality is sufficient.
3. One-page infographic summary The infographic (Supplement 5) provides a quick visual reference guide, summarizes the key messages of the Comprehensive Guide and provides a memory aid to users who have previously read the full guideline. Its purpose is to improve the translation of the guideline into standard practice.

#### 2.3.4 Dissemination, Implementation and Review

The working party designed the dissemination and implementation plan. Dissemination will occur at key annual meetings and conferences where target markets, such as junior researchers, applications-oriented scientists, and educators will be in attendance. This includes the International Society for Magnetic Resonance in Medicine (ISMRM), Society for MR Radiographers & Technologists (SMRT) and Organization for Human Brain Mapping (OHBM). In addition, the guideline will be presented in GABA-MRS-focused workshops and educationals such as the International Symposium on GABA and Advanced MRS and EDITINGSCHOOL, where attendees have a specific interest in GABA MRS.

Pilot implementation will commence at all of the working parties’ collaborative sites (over 25 sites worldwide), where the guideline will be integrated in current operating procedures, and the infographic will be distributed. In addition, members of the guideline working party will integrate it into their supervision and teaching procedures to students (graduate and undergraduate), residents and researchers. The guideline will be reviewed for currency by the working party in 2026 and updated should further high-quality evidence provide recommendations differing to those presented in this guideline.

## 3. RESULTS

The final guideline consisted of 23 recommendations, under six domains essential for GABA MRS acquisition; Parameters, Practicalities, Data acquisition, Confounders, Quality/reporting, Post-processing. Overall 78.3% of recommendations were formed from high quality evidence (Level A or B) and 91.3% received agreement from over 80% of the expert panel (Table 3).

**Table 3.**
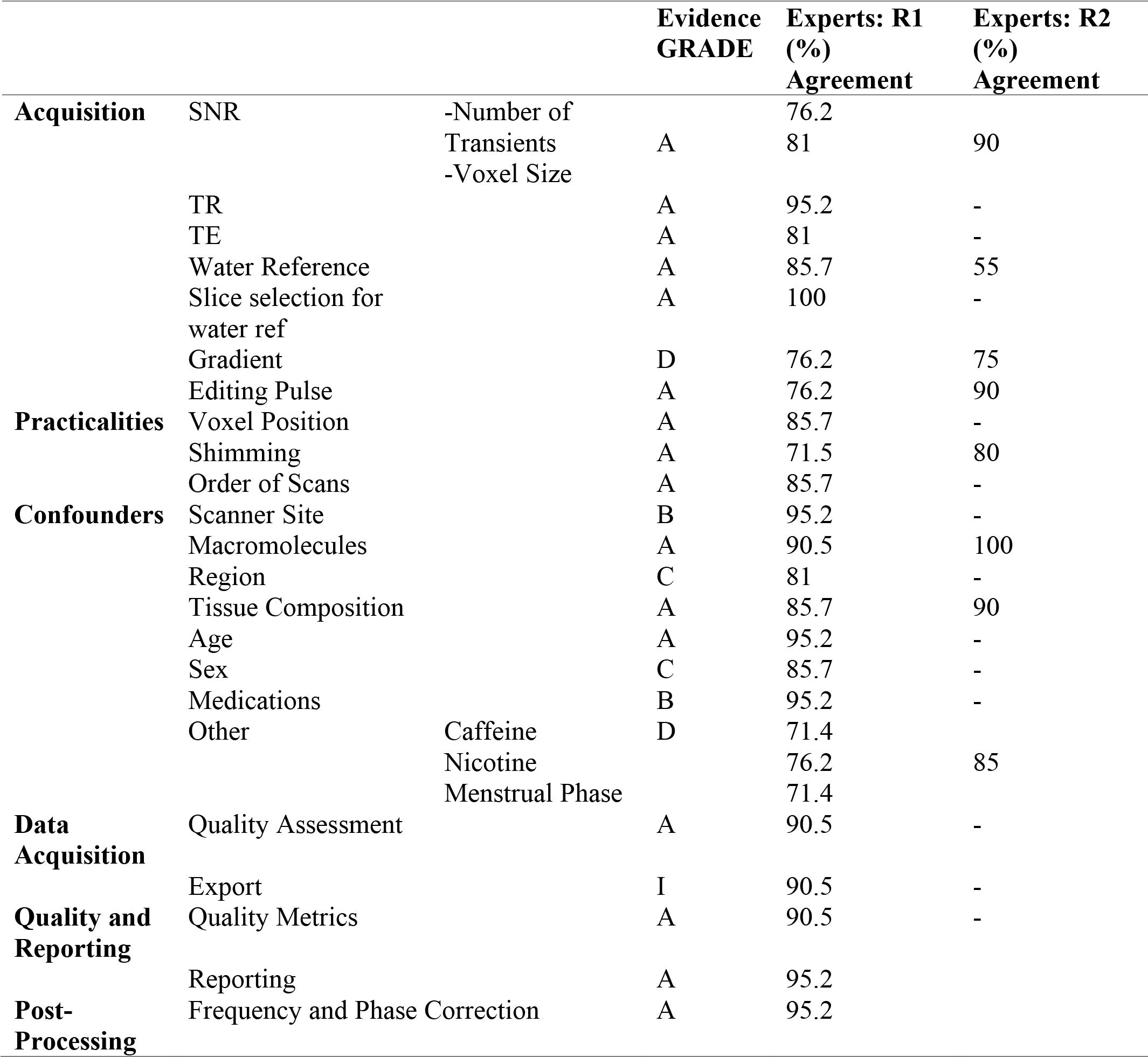
SUMMARY OF RECOMMENDATIONS.

### 3.1 PARAMETERS

#### 3.1.1 Signal-to-Noise Ratio Considerations (Number of transients and Voxel volume)

**ADAPT: Start with at least 192 transients (i.e. 96 Edit-ON + 96 Edit-OFF) and a voxel volume of 27 ml (e.g 3** ’ **3** ’ **3 cm^3^) to quantify GABA when scanning a favourable brain region.**

-Consider increasing the total number of transients when scanning smaller or more challenging brain regions (see 3.3.3 *Region*).

##### **Evidence GRADE A.** Round 1 Expert Panel Agreement number of transients 76.2%, voxel size 81%, Round 2 Expert Panel Agreement 90%

There were eight studies (Level 1 to Level 4) (Bhattacharyya *et al*., 2007; Harris *et al*., 2014; Mullins *et al*., 2014; Brix *et al*., 2017; Mikkelsen *et al*., 2017; Mikkelsen *et al*., 2018; Sanaei Nezhad *et al*., 2018; Mikkelsen *et al*., 2019) with recommendation about the number of transients, and seven studies (Level 1 to Level 4) (Mullins *et al*., 2014; Bai *et al*., 2015; Bergmann *et al*., 2016; Chen *et al*., 2017; Mikkelsen *et al*., 2017; de Graaf, 2019; Mikkelsen *et al*., 2019) with recommendation about voxel volume. The studies recommended using a range of transients from 126 (Sanaei Nezhad *et al*., 2018) to 320 (Mikkelsen *et al*., 2017; Mikkelsen *et al*., 2019) transients, with the majority recommending a minimum of 192 transients when using a voxel volume of e.g 3 ’ 3 ’ 3 cm^3^. A further two studies (Level 1)(Peek *et al*., 2020; Lin *et al*., 2021) highlight the importance of reporting the number of transients used and whether they refer to the total number of transients or separate (as Edit-ON and Edit-OFF). Failure to achieve adequate signal to noise has a significant effect on quality of the spectra as demonstrated in Figure 4. Round 1 agreement for this recommendation was 76.2% for number of transients and 81% for voxel size. Two key considerations were made: first, experts recommended combining the two separate recommendations to highlight the interdependence of the number of transients and voxel size. Second, the number of transients are best selected in multiples of 16 to allow for full phase cycles to be included. Round 2 agreement increased to 90%. Therefore, the revised recommendation was accepted.

**Figure 4:**
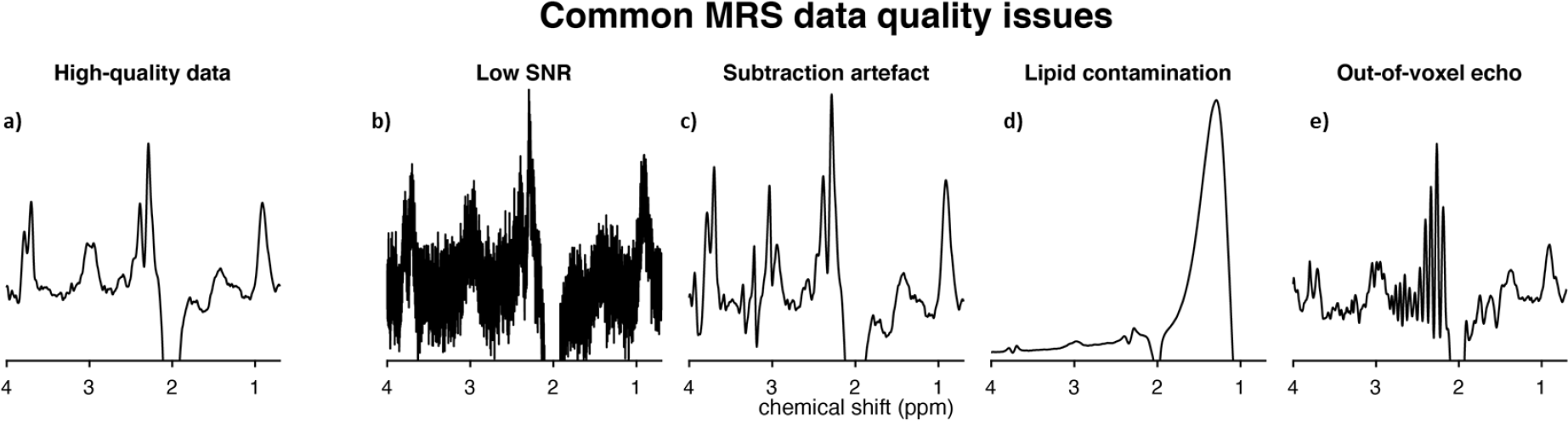
Common MEGA-PRESS data quality issues. a) High-quality data with sufficient SNR, narrow linewidths, a well-defined edited signal at 3 ppm, and no substantial artefacts; b) very high noise levels due to low number of transients or small voxel volume; c) severe subtraction artefacts due to scanner frequency drift; d) lipid contamination due to participant motion or voxel positioning too close to the skull; e) out-of-voxel echo (“ghost signal”).

#### 3.1.2 Repetition Time (TR)

**ADOPT: Use a TR of around 2000 ms at 3T.**

##### **Evidence GRADE A**; Round 1 Expert Panel Agreement 95.2%

There were five studies (Level 1 to Level 3) (Puts *et al*., 2013; Mikkelsen *et al*., 2017; Deelchand *et al*., 2019; Mikkelsen *et al*., 2019; Wilson *et al*., 2019) that provided recommendations on TR. The studies all concur that a TR of ∼2000ms is suitable for the measurement of GABA with edited MRS. Round 1 agreement was 95.2%. Therefore, this recommendation was adopted.

#### 3.1.3 Echo Time (TE)

**ADOPT: TE should be 68 ms (GABA+); 80 ms (macromolecule-suppressed GABA)**

##### **Evidence GRADE A**; Round 1 Expert Panel Agreement 81%

There were ten studies (Level 1 to Level 3) (Mescher *et al*., 1998; Edden *et al*., 2012; Mullins *et al*., 2014; Harris *et al*., 2015a; Edden *et al*., 2016; Mikkelsen *et al*., 2017; Deelchand *et al*., 2019; Mikkelsen *et al*., 2019; Wilson *et al*., 2019; Cudalbu, 2020) that provided recommendation on TE. The consensus across studies was to keep TE as close to 68 ms as possible when estimating GABA+, and 80 ms for macromolecule-suppressed measurements. Round 1 agreement was 81%. Therefore, this recommendation was accepted.

ADAPT: Water reference scans (required for eddy-current correction and water-scaled quantification): acquire two water reference scans for each volume of interest: one using the same parameters as MEGA-PRESS, but deactivated water suppression for eddy-current correction, and one short-TE (TE ∼ 30 ms) for quantification.

##### **Evidence GRADE A**; Round 1 Expert Panel Agreement 85.7%, Round 2 Expert Panel Agreement 55%

There were seven studies (Level 1 to Level 3) (Hall *et al*., 2014; Mullins *et al*., 2014; Oeltzschner *et al*., 2016; de Graaf, 2019; Wilson *et al*., 2019; Near *et al*., 2020; Öz *et al*., 2020) that provided recommendation on water reference scans. There was consensus across studies recommending that water reference scans are acquired from the same volume of interest using the same parameters and gradients in order to facilitate eddy-current correction. Round 1 agreement was 85.7%, but only 70% felt the recommendation was suitable for a beginner’s guide. Experts reasoned that those new to the field might not be aware that using long-TE water data for quantification purposes may introduce T2-weighting, which inadvertently has implications for quantification (Gasparovic *et al*., 2018). In line with the feedback and the publication of a new consensus document (Near *et al*., 2020), the recommendation was revised to recommend acquiring a separate short-TE water reference scan to be used for quantification. However, Round 2 agreement reduced to 55% due to several experts (n=8/20, 40%) not considering a short=TE scan necessary for quantification. The inclusion of this guideline was discussed by the working party. The decision was made to retain the revised recommendation due to it reflecting the most up-to-date recommendation in the literature. It was decided to further develop the preface and consideration section for educational purposes to help the translation of this new recommendation, given the feedback from the experts.

#### 3.1.4 Slice-selection centre frequency of water reference scan

**ADOPT: Set the water reference to be acquired from the same volume as the GABA signal.**

##### **Evidence GRADE B**; Round 1 Expert Panel Agreement 100%

There were three studies (Level 2 to Level 3) (Mikkelsen *et al*., 2017; Deelchand *et al*., 2019; Mikkelsen *et al*., 2019) that provided recommendations on slice-selection centre frequency of the water reference scan. The consensus across studies was that the frequency should be set to 0 ppm offset, i.e. localizing the 4.7 ppm water signal. Round 1 agreement was 100%.

Therefore, this recommendation was accepted.

#### 3.1.5 Order of slice-selective gradients

**ADAPT: When artefacts appear in pilot data, consider changing the order of the slice-selective gradients for each volume of interest.**

##### **Evidence GRADE D**; Round 1 Expert Panel Agreement 76.2%; Round 2 Expert Panel Agreement 75%

There was one paper (Level 3) (Ernst and Chang, 1996) that provided recommendations on the order of slice-selective gradients. The paper highlighted how changing the order of gradients can remove artefacts from data. Round 1 agreement was 76.2% due the recommendations suggesting that trial acquisitions with different orders should be conducted. In line with feedback from expert consensus, the recommendation was revised to suggest this as a troubleshooting option only when artefacts are consistently present in data. Round 2 agreement reduced to 75% due to concerns that those new to the field would not know which artefacts could be helped by changing gradient order (n=3/20, 15%) and that some systems do not allow for simple adjustment of gradient order. The decision to maintain the recommendation was made by the MRS sub-committee who felt this troubleshooting advice might be helpful to those new to the field with the addition of Figure 4, which demonstrates the artefacts that can be addressed through this method. This recommendation therefore was included, but not given expert approval.

#### 3.1.6 Editing pulse specifications

ADOPT: Editing pulses can be applied as follows (Table 4):

**Table 4.** editing pulse specifications.

|  | GABA+ | Macromolecule-suppressed |
| --- | --- | --- |
| Frequency (ppm) |  |  |
| Edit-ON | 1.9 ppm | 1.9 ppm |
| Edit-OFF | 7.46 ppm | 1.5 ppm |
| Bandwidth | 60 Hz | Usually 80 Hz (60 Hz on some implementations) |
| Spacing | 0.5 TE apart (this parameter is usually not accessible to the user) |  |

##### **Evidence GRADE A**; Round 1 Expert Panel Agreement 76.2%; Round 2 Expert Panel Agreement 90%

There were nine studies (Level 1 to Level 3) (Keltner *et al*., 1996; Mescher *et al*., 1998; Henry *et al*., 2001; Mullins *et al*., 2014; Edden *et al*., 2016; Mikkelsen *et al*., 2017; Deelchand *et al*., 2019; Mikkelsen *et al*., 2019; Saleh *et al*., 2019) that provided recommendation on editing pulse parameters. Recommendations were dependent on whether GABA+ or macromolecule-suppressed GABA was being acquired (see Supplement 4 full-length guideline for explanation). Round 1 agreement was 76.2%, the recommendation was therefore revised. Key points from the expert panel were that some sequence implementations do not allow for the adjustment of these parameters. The panel had many suggestions of variations that they use when applying editing pulses (n=8/21, 38.1%) which highlight the methodological heterogeneity even among experts in the MRS field. The revised recommendation removed recommendations for pulse duration and highlighted that editing pulses could be applied using these parameters as a starting point for those new to the field.

Round 2 expert panel agreement was 90%. Therefore, this revised recommendation was accepted.

### 3.2 PRACTICALITIES

#### 3.2.1 Voxel position

**ADAPT: Use automated voxel positioning tools where available. If manually positioning the voxel, use a screenshot and clear instructions regarding positioning relative to anatomical landmarks and degree of rotation.**

##### **Evidence GRADE A**; Round 1 Expert Panel Agreement 85.7%

There were five studies (Level 1 to Level 4) (Kreis, 2004; Bai *et al*., 2017; Chen *et al*., 2017; Park *et al*., 2018; Öz *et al*., 2020) that provided recommendations on positioning of the voxel. The studies recommended use of an automated voxel positioning tool. Although the expert panel agreed with this recommendation, 28.6% of experts highlighted that fully automated voxel positioning is not currently available as standard. Round 1 agreement was 85.7%.

Therefore, this recommendation was accepted.

#### 3.2.2 Shimming

**ADAPT: A beginner should use a readily available automated field-map-based shim and minimize the use of manual adjustments.**

##### **Evidence GRADE A**; Round 1 Expert Panel Agreement 71.5%; Round 2 Expert Panel Agreement 80%

There were eight studies (Level 1 to Level 3) (Gruetter and Tkáč, 2000; Saleh *et al*., 2016a; Juchem and de Graaf, 2017; Deelchand *et al*., 2018; Sanaei Nezhad *et al*., 2018; Wilson *et al*., 2019; Öz *et al*., 2020; Juchem *et al*.) that provided recommendation on shimming to maximize the homogeneity of the static magnetic field (B0). The studies demonstrated that projection-based shim optimisation or second-order pencil beam methods could provide narrower linewidths than the default 3D field map-based methods. These specific techniques may not be readily available on all systems, therefore the expert panel recommends that any readily-available automated field map-based methods are used with minimal manual adjustments where possible (9/21, 43%). Round 1 agreement was 71.5 %, subsequent adjustments were therefore made to highlight that linewidths are calculated differently by different vendors (see considerations in extended document). Despite evidence suggesting projection-based shim optimisation might achieve narrower linewidths, the recommendation states the beginner should use readily available field-map based shim methods. Round 2 expert agreement was 80%. Therefore, the revised recommendation was accepted.

#### 3.2.3 Order of scans and field drift

**ADOPT: Where possible, MRS should be conducted prior to gradient-heavy acquisitions or in small blocks of 2-5 minutes with frequency adjustments between adjustment blocks. Consider using real-time frequency correction if available.**

##### **Evidence GRADE A**; Round 1 Expert Panel Agreement 85.7%

There were seven studies (Level 1 to Level 3) (Harris *et al*., 2014; Edden *et al*., 2016; Mikkelsen *et al*., 2017; Andronesi *et al*., 2020; Cudalbu, 2020; Öz *et al*., 2020; Choi *et al*., 2021) that provided recommendation on the order of scans and the effect it has on field drift. The studies highlighted the negative impact gradient-heavy scanning (e.g. diffusion tensor imaging) has on frequency drift during subsequent MRS scans. Previous recommendations were to avoid scanning after gradient-heavy acquisitions, however owing to this not being practical due to scan scheduling problems, a recent consensus document made a new proposal. The recommendation was to acquire MRS data in small blocks with frequency adjustment after each block whilst monitoring the residual water signal on the inline display during the scan acquisition in order to detect drift. Round 1 agreement was 85.7%. Therefore, this recommendation was accepted.

### 3.3 CONFOUNDERS

#### 3.3.1 Scanner site and vendor

**ADOPT: In multi-site studies, standardised protocols should be used, and the degree of systematic differences between site/scanner should be reported.**

##### **Evidence GRADE B**; Round 1 Expert Panel Agreement 95.2%

There were three multi-site studies (Level 2) (Mikkelsen *et al*., 2017; Mikkelsen *et al*., 2019; Saleh *et al*., 2019) that provided recommendations on managing scanner site and different vendors as a confounder of GABA. The studies reported a coefficient of variation across all data sets of around 12% for GABA+/Cr and 17% for water-scaled GABA+. Macromolecule-suppressed MEGA-PRESS had larger CVs of 28%-29% for both GABA/Cr and water-scaled GABA (Mikkelsen *et al*., 2017; Mikkelsen *et al*., 2019). Round 1 expert panel agreement was 95.2%. Therefore, this recommendation was accepted.

#### 3.3.2 Macromolecules

**ADAPT: A beginner should use conventional MEGA-PRESS reporting GABA+. Macromolecule contamination should be acknowledged as a limitation, and consideration paid to whether macromolecules could be responsible for between-group differences.**

##### **Evidence GRADE A**; Round 1 Expert Panel Agreement 90.5%; Round 2 Expert Panel Agreement 100%

There were twelve studies (ranging from Level 1 to Level 3) (Mullins *et al*., 2014; Cudalbu, 2020; Choi *et al*., 2021) (Henry *et al*., 2001; Harris *et al*., 2015a; Edden *et al*., 2016; Mikkelsen *et al*., 2016b; Shungu *et al*., 2016; Gu *et al*., 2018; Oeltzschner *et al*., 2018a; Duncan *et al*., 2019) that provided recommendation on macromolecule contamination as a confounder of GABA. Contrary to the original consensus document for MEGA-PRESS (Level 1) (Mullins *et al*., 2014), the latest consensus documents recommend the use of macromolecule-suppressed editing where possible (Level 1)(Cudalbu, 2020). However, both consensus documents acknowledge this approach has a number of limitations including its susceptibility to frequency drift. The expert panel agreed that a macromolecule-suppressed study is more difficult to control and run as a beginner and therefore endorsed the recommendation that a beginner should acquire GABA+ data. Both consensus documents agree that in cases where GABA+ is acquired, results must be reported as GABA+ macromolecules, with macromolecule contaminations explicitly acknowledged as a limitation. Round 1 expert panel agreement was 90.5%. This recommendation was revised following publication of a new consensus document and therefore sent out for Round 2 grading despite achieving over 80% expert panel agreement on Round 1. Round 2 agreement was 100%. Therefore, the revised recommendation was accepted.

#### 3.3.3 Region

**ADAPT: Select brain regions relevant to the research question, however, acknowledge that brain regions have differing reliability with respect to data acquisition.**

##### **Evidence GRADE C**; Round 1 Expert Panel Agreement 81%

There were fourteen studies (Level 1^T^ to Level 4 ^T^) (Gruetter and Tkáč, 2000; Harada *et al*., 2011; Waddell *et al*., 2011; Puts and Edden, 2012; Gao *et al*., 2013; van der Veen and Shen, 2013; Harris *et al*., 2015c; Long *et al*., 2015; Greenhouse *et al*., 2016; Brix *et al*., 2017; Chen *et al*., 2017; Porges *et al*., 2017; Puts *et al*., 2018; Dhamala *et al*., 2019) that provided recommendation on brain region as a confounder of GABA levels. The studies demonstrated that GABA levels appear to be region-specific rather than reflective of a global GABAergic tone as once proposed (Puts and Edden, 2012). Therefore, it is important to consider the suitability of the brain region for ^1^H-MRS acquisition and recognize that different brain regions have different reliability with respect to signal-to-noise ratio and the likelihood of artefacts. Round 1 agreement was 81%. Therefore, this recommendation was accepted.

#### 3.3.4 Tissue composition

**ADAPT: Water-scaled quantification methods should consider the impact of partial volume effects on GABA estimation.**

-Segmented structural images should be used along with a tissue-correction method to account for grey matter, white matter and cerebrospinal fluid composition of the voxel. Grey-matter only correction should be avoided.

##### **Evidence GRADE A**; Round 1 Expert Panel Agreement 85.7%; Round 2 Expert Panel Agreement 90%

There were nine studies (Level 1 to Level 3) (Bhattacharyya *et al*., 2011; Geramita *et al*., 2011; Mullins *et al*., 2014; Harris *et al*., 2015b; Mikkelsen *et al*., 2016a; Porges *et al*., 2017; Gasparovic *et al*., 2018; Puts *et al*., 2018; Choi *et al*., 2021) that provided recommendation on tissue composition as a confounder of GABA estimation. The studies agreed that GABA levels were higher in grey matter than white matter, and therefore needed to be accounted for when quantifying GABA. The additional considerations from the expert panel were that tissue composition should be considered as a covariate in order to clarify whether between-group differences were being driven by GABA levels rather than tissue composition (n= 3/21, 14.3%). Round 1 agreement was 85.7%. However, the original recommendation included a significant number of caveats. Therefore, to improve clarity the recommendation was revised, where the caveats were removed from the recommendation and placed in the considerations section of the full document. Round 2 agreement was 90%. Therefore the revised recommendation was accepted.

#### 3.3.5 Age

**ADOPT: Age is likely to affect GABA levels, therefore age should be accounted for in study design or statistical analysis.**

##### **Evidence GRADE A**; Round 1 Expert Panel Agreement 95.2%

There were seven studies (Level 1^T^ to Level 3^T^) (Aufhaus *et al*., 2013; Gao *et al*., 2013; Porges *et al*., 2017; Maes *et al*., 2018; Marenco *et al*., 2018; Simmonite *et al*., 2019; Porges *et al*., 2020) that provided recommendations on age as a confounder of GABA levels. The studies all suggest that GABA+ decreases with age in adulthood. The recent meta-analysis (Porges *et al*., 2020) describes an early period of increase in frontal GABA levels, which stabilized throughout adulthood, and then decreased with aging. Round 1 agreement was 95.2%. Therefore, this recommendation was accepted.

#### 3.3.6 Sex

**ADOPT: Sex is likely to impact on GABA levels, therefore sex should be accounted for in study design or statistical analysis.**

##### **Evidence GRADE C**; Round 1 Expert Panel Agreement 85.7

There were four studies (Level 3^T^ to Level 4^T^) (O’Gorman *et al*., 2011; Aufhaus *et al*., 2013; Gao *et al*., 2013; Saleh *et al*., 2017) that provided recommendation on sex as a confounder for GABA levels. The variation in outcome across the studies suggest that differences in GABA levels between males and females may be region-specific. Round 1 agreement was 85.7%. Therefore, this recommendation was accepted.

#### 3.3.7 Medications

**ADAPT: Medications may impact GABA levels, as minimum best practice all medications should be recorded**.

-Consider excluding participants taking medications likely to affect the GABAergic system.

##### **Evidence GRADE B**; Round 1 Expert Panel Agreement 95.2%

There were eight studies (Level 1^T^ to Level 4^T^) (Rothman *et al*., 1993; Petroff *et al*., 1996a; Petroff *et al*., 1996b; Bhagwagar *et al*., 2004; Licata *et al*., 2009; Cai *et al*., 2012; Puts and Edden, 2012; Myers *et al*., 2014) that provided recommendations on medications that may confound GABA. The studies reported that medications that alter GABA concentration directly and those that affect GABA receptor agonists and antagonists may both influence brain GABA levels. Round 1 agreement was 95.2%. Therefore, this recommendation was accepted.

#### 3.3.8 Other potential confounders: Nicotine, Caffeine, Phase of menstrual cycle

**ADAPT: Potential confounders such as caffeine and nicotine intake and phase of menstrual cycle may affect GABA levels, as minimum best practice potential confounders should be recorded**.

##### **Evidence GRADE D**; Round 1 Expert Panel Agreement Caffeine 71.4%, Nicotine 76.2%, Phase of Menstrual Cycle 71.4%, Round 2 Expert Panel Agreement 85%

There were six studies (Level 3 ^T^ to 4^T^) (Epperson *et al*., 2002; Epperson *et al*., 2005; Harada *et al*., 2011; De Bondt *et al*., 2015; Schulte *et al*., 2017; Oeltzschner *et al*., 2018b) that provided recommendation on other potential confounders of GABA levels which included caffeine, nicotine, and phase of menstrual cycle. The studies were inconclusive to the degree of effect these potential confounders may have on GABA levels. Round 1 agreement was caffeine 71.4%, nicotine 76.2%, phase of menstrual cycle 71.4%. Expert panel feedback was that there was not sufficient high-quality evidence confirming these factors as confounders of GABA levels, and therefore the expert panel did not feel it was essential to control for all in study design. The recommendation was adjusted to reflect this. Round 2 agreement was 85%. Therefore, the revised recommendation was accepted.

### 3.4 DATA ACQUISITION

#### 3.4.1 Quality assessment during the scan

**ADOPT: It is recommended to monitor the quality of the acquisition using the inline data display at time of scanning.**

-Scans should be cancelled, and voxel position adjusted if evidence of weak water suppression, strong lipid contamination or other artefacts.

##### **Evidence GRADE A**; Round 1 Expert Panel Agreement 90.5%

There were two studies (Level 1) (Öz *et al*., 2020; Choi *et al*., 2021) that provided recommendations on quality assessment during the scan. Both recommended that the MR operator should evaluate and monitor water suppression efficiency, spectral linewidth and signal-to-noise ratio at the beginning and during the MRS acquisition. Round 1 expert panel agreement was 90.5%. Therefore, this recommendation was accepted.

#### 3.4.2 Data export

**DEVELOP: Export data in a format that saves individual transients to allow adequate post-processing.**

##### **Evidence GRADE I**; Round 1 Expert Panel Agreement 90.5%

There were no studies discussing file format export for MEGA-PRESS acquisitions. The recommendation was therefore developed based on a consensus document that made recommendations on the file format to export for ^1^H-MRS studies which also can be applied to MEGA-PRESS acquisitions (Near *et al*., 2020). Round 1 expert panel agreement was 90.5%. Therefore, the developed recommendation was accepted.

### 3.5 QUALITY AND REPORTING

#### 3.5.1 Quality Metrics

ADOPT: Report spectral quality in terms of the signal-to-noise ratio, linewidth, water suppression efficiency, fit quality, and the presence of unwanted spectral features

##### **Evidence GRADE A**; Round 1 Expert Panel agreement 90.5%

There were seven studies (Level 1 to Level 3) (Bolliger *et al*., 2013; Mullins *et al*., 2014; Kreis, 2016; Chen *et al*., 2017; Deelchand *et al*., 2018; Wilson *et al*., 2019; Öz *et al*., 2020) that provided recommendations on which variables should be used to assess data quality. The studies agree that spectral quality should be assessed using a number of aspects including signal-to-noise ratio, linewidth, water suppression efficiency, modelling quality, and presence of unwanted spectral features. Round 1 agreement was 90.5%. Therefore, this recommendation was accepted.

#### 3.5.2 Reporting

ADOPT: When reporting results use one of these two checklists (MRS in MRS, Lin *et al*. 2020 or MRS-Q, Peek *et al*. 2020) using the appropriate terminology (Kreis *et al*. 2020). Include detailed reporting of hardware, MEGA-PRESS-specific acquisition parameters, quantification details, quality metrics, and analysis methods.

##### **Evidence GRADE A**; Round 1 Expert Panel Agreement 95.2%

There were three studies (Level 1 to Level 3) (Deelchand *et al*., 2019; Peek *et al*., 2020; Lin *et al*., 2021) that provided recommendations on reporting in MEGA-PRESS GABA studies. Two studies provided checklists that could be utilized to improve reporting in these studies. Round 1 agreement was 95.2%. Therefore, this recommendation was accepted.

### 3.6 POST-PROCESSING

#### 3.6.1 Frequency-and-Phase Correction (Post-processing)

**ADOPT: Frequency-and-phase alignment of individual transients should be performed during post-processing.**

##### **Evidence GRADE A**; Round 1 Expert Panel Agreement 95.2%

There were ten studies (Level 1 to Level 3) (Edden and Barker, 2007; Edden *et al*., 2014; Harris *et al*., 2014; Near *et al*., 2015; Cleve *et al*., 2017; van der Veen *et al*., 2017; Wiegers *et al*., 2017; Tapper *et al*., 2019; Near *et al*., 2020; Choi *et al*., 2021) that provided recommendation on frequency-and-phase correction. The studies found that using frequency-and-phase correction was able to significantly improve editing efficiency. Round 1 agreement was 95.2%. Therefore, this recommendation was accepted.

## 4 DISCUSSION

The Comprehensive Guide to MEGA-PRESS for GABA Measurement presented in this manuscript, was developed following a translational framework to produce robust, user-friendly guidelines for those new to the field of MEGA-PRESS. The key strengths of this approach were conducting a systematically delivered scoping review to inform the evidence synthesis and the involvement of multiple stakeholders with diverse experience and expertise. Further, we performed blinded GRADEing of the quality of evidence for each recommendation, and then finally incorporated expert peer review through the modified-Delphi process. The result was a guideline with 23 recommendations; 73.9% of these recommendations had a high GRADE of evidence and high expert panel agreement, 4.4% had a high GRADE of evidence but low expert panel agreement, 17.4% had a low GRADE of evidence and high expert panel agreement, and 4.4% had a low GRADE of evidence and low expert panel agreement. Reasons for the differences between GRADE of evidence and the degree of expert panel agreement plus the decisions to retain the recommendations that did not gain expert panel approval are discussed below.

Both 3.1 ‘Parameters’ and 3.2 ‘Practicalities’ sections contain recommendations with a high GRADE of evidence and a high percentage of expert panel agreement, with the exception of just two recommendations (Table 3). This high level of evidence supported by the expert panel encourages confidence in our recommendations, as expert evaluation reflects practice in experienced MRS groups worldwide. The two recommendations in the guideline that were not sufficiently endorsed by the experts were both in the 3.2 ‘Practicalities’ section; *Order of slice-selective gradients* (Evidence GRADE I, Experts: 75% agreement, 25% neutral) and *Water reference scans for eddy current correction and water-scaled quantification* (Evidence GRADE A, Experts 55% agreement, 20% neutral, 25% disagreement). This lower level of expert panel agreement suggests that these recommendations are less reflective of current standard practice. The MRS sub-committee saw an opportunity to encourage the translation of evidence to practice and decided to keep the recommendations in the guideline without expert panel endorsement but with further discussion: Firstly, while experts were concerned that *Order of slice selective gradients* is not applicable to all systems, the MRS sub-committee found it a valuable troubleshooting option worth adopting as regular practice on systems where it is available. Secondly*, Water reference scans for eddy-current correction and quantification* failed to gain expert panel endorsement following the addition of a separate short-TE scan for quantification, although this practice is recommended in the latest consensus document (Near *et al*., 2020). The MRS sub-committee maintained this recommendation to further facilitate the implementation of this new recommendation into practice.

The 3.3 ‘Confounders’ section contained recommendations with generally lower levels of evidence but achieved high expert panel agreement. Firstly, five of the eight recommendations in this domain were assessed using a traditional hierarchy of evidence, where Level 1 evidence represents a systematic literature review of randomised controlled trials, a study design that has not been frequently adopted in the field of MRS to date.

Secondly, many of the recommendations in this section reflect principles and practices historically adopted from expert opinion and practical experience rather than from clear and systematic evidence collection. This explains the high level of expert agreement, but also shows further high quality research is required to establish the degree of confounding these factors present.

The 3.4 ‘Data Acquisition’, 3.5 ‘Quality and Reporting’ and 3.6 ‘Post-processing’ sections generally had high levels of evidence and high expert panel agreement. The high levels of evidence adapted from consensus documents and high expert panel agreement reflect that areas included in these domains are topical, relevant and are considered important in the acquisition of GABA using MEGA-PRESS. The one recommendation that had no evidence (Level I), and therefore required active development instead of adoption or adaption was *File export*. Previous consensus documents may not have included this explicit recommendation as it might be considered ‘assumed knowledge’, but the MRS-subcommittee and stakeholders valued its inclusion given the intended audience. This is especially relevant since failure to save the correct file type at time of scanning prevents appropriate post-processing and compromises data quality considerably; an easily-avoidable mistake that has been commonly observed by the MRS sub-committee.

The results of the evidence synthesis did not always provide recommendations suitable for a those new to the field. In two instances (*Shimming* and *Macromolecules*), the MRS sub-committee and expert panel agreed that a beginner would likely achieve a better result using a different approach to that recommended in the most recent consensus documents (Cudalbu, 2020; Juchem *et al*., 2021). An example was *shimming*: a recent consensus document (Juchem *et al*., 2021) recommends use of a tool that is not readily available on all systems, has limited technical support, requires approved distribution from its developers, and is more technically challenging to operate than system based shim methods (FASTMAP). Whilst proof-of-concept studies (Grewal *et al*., 2016; Saleh *et al*., 2016b; Deelchand *et al*., 2018) have demonstrated that narrower linewidths can be achieved using this approach compared to readily available automated field map-based methods, it requires specific expertise to be set up and used. Therefore, the MRS sub-committee and expert panel recommend that a beginner use a readily-available automated field-map-based shim method. Doing so is well-established in the field and should produce sufficient B0 homogeneity to generate high-quality spectra (Mullins *et al*., 2014). In summary, while this recommendation is not consistent with the recent consensus document, it is directly aligned with our aim of enabling a beginner to produce high-quality MEGA-PRESS spectra for the reliable quantification of GABA.

The second recommendation adapted for the beginner in our guideline was *Macromolecules* and how they should be handled. Feedback from 90.5% (20/21) experts was that beginners should choose sequence parameters to acquire GABA plus macromolecule (GABA+) data, despite the latest consensus document recommending the acquisition of macromolecule-suppressed data. This is supported by a previous consensus document (Mullins *et al*., 2014) and methodological publications (Mullins *et al*., 2014; Harris *et al*., 2015a; Mikkelsen *et al*., 2017) that all agree that symmetric macromolecule suppression is an order of magnitude more susceptible to frequency drift and that other methods of macromolecule signal removal all have substantial technical and practical limitations (Mullins *et al*., 2014; Harris *et al*., 2015a; Mikkelsen *et al*., 2017). The MRS sub-committee reviewed the evidence once more, and decided that despite our recommendation differing from the latest consensus document (Cudalbu, 2020), acquiring GABA+ currently offers the most robust, reliable and widely used method to measure GABA levels for a beginner user. Further the likelihood of failure acquiring GABA+ is substantially lower than if they were to use the delicate macromolecule suppression. Therefore, the recommendation to acquire GABA+ data and acknowledge the macromolecule contamination as a limitation (or discuss as a potential source of observed effects) was deemed most suitable for inclusion in the Comprehensive Guide.

The scope of the Comprehensive Guide was to largely focus on study design and data acquisition. We considered it to be beyond the scope to discuss further details of post-processing (beyond frequency-and-phase correction and the file format export it requires), modelling, or quantification of MEGA-PRESS data. We therefore direct the reader to comprehensive efforts on best practices in MEGA-PRESS (Mullins *et al*., 2014) and two recent consensus papers on pre-processing, modelling and quantification (Near *et al*., 2020) and spectral editing in general (Choi *et al*., 2021). Further, the beginner is advised to liaise with representatives from their vendor and sequence developers with regard to system-specific functions that may or may not be available, as highlighted throughout this Comprehensive Guide. Finally, the MRSHub (https://www.mrshub.org) provides an online resource hosting processing and analysis software, normative example data, and a discussion forum frequented by beginners and experts alike where questions about study design and protocol can be posed.

In conclusion, this Comprehensive Guide combines a robust evidence synthesis on the measurement of GABA levels with edited MRS and expert panel review. The result is an evidence-based, peer-reviewed guideline for those new to using MEGA-PRESS including higher degree research students, clinician-researchers, MRI technicians or anyone new to the field of MEGA-PRESS. The guideline helps to ensure sufficient quality of acquisition and reporting is achieved. The high level of agreement between evidence and expert assessment instils confidence in the validity, longevity, and applicability of these recommendations. The full accompanying documentation is freely available online here: https://osf.io/5v9cp/?view_only=b6c70761abdd4d6e9e38d2a7b9944f9f

## Supporting information

Supplement 4: Full Guideline

Supplement 5: Infographic

## Data Availability

All data produced in the present work are contained in the manuscript

https://osf.io/5v9cp/?view_only=b6c70761abdd4d6e9e38d2a7b9944f9f

## REFERENCES

1. Andronesi OC, Bhattacharyya PK, Bogner W, Choi I-Y, Hess AT, Lee P, et al. Motion correction methods for MRS: experts’ consensus recommendations. NMR in Biomedicine 2020; n/a(n/a): e4364.

2. Aufhaus E, Weber-Fahr W, Sack M, Tunc-Skarka N, Oberthuer G, Hoerst M, et al. Absence of changes in GABA concentrations with age and gender in the human anterior cingulate cortex: a MEGA-PRESS study with symmetric editing pulse frequencies for macromolecule suppression. Magnetic resonance in medicine 2013; 69(2): 317–20.

3. Bai X, Edden RA, Gao F, Wang G, Wu L, Zhao B, et al. Decreased gamma-aminobutyric acid levels in the parietal region of patients with Alzheimer’s disease. Journal of Magnetic Resonance Imaging 2015; 41(5): 1326–31.

4. Bai X, Harris AD, Gong T, Puts NAJ, Wang G, Schär M, et al. Voxel Placement Precision for GABA-Edited Magnetic Resonance Spectroscopy. Open J Radiol 2017; 7(1): 35–44.

5. Baulac S, Huberfeld G, Gourfinkel-An I, Mitropoulou G, Beranger A, Prud’homme J-F, et al. First genetic evidence of GABA A receptor dysfunction in epilepsy: a mutation in the γ2-subunit gene. Nature genetics 2001; 28(1): 46–8.

6. Bergmann J, Pilatus U, Genc E, Kohler A, Singer W, Pearson J. V1 surface size predicts GABA concentration in medial occipital cortex. Neuroimage 2016; 124(Pt A): 654–62.

7. Bhagwagar Z, Wylezinska M, Taylor M, Jezzard P, Matthews PM, Cowen PJ. Increased brain GABA concentrations following acute administration of a selective serotonin reuptake inhibitor. The American journal of psychiatry 2004; 161(2): 368–70.

8. Bhandage AK, Cunningham JL, Jin Z, Shen Q, Bongiovanni S, Korol SV, et al. Depression, GABA, and Age Correlate with Plasma Levels of Inflammatory Markers. Int J Mol Sci 2019; 20(24).

9. Bhattacharyya PK, Lowe MJ, Phillips MD. Spectral quality control in motion-corrupted single-voxel J-difference editing scans: an interleaved navigator approach. Magnetic resonance in medicine 2007; 58(4): 808–12.

10. Bhattacharyya PK, Phillips MD, Stone LA, Lowe MJ. In vivo magnetic resonance spectroscopy measurement of gray-matter and white-matter gamma-aminobutyric acid concentration in sensorimotor cortex using a motion-controlled MEGA point-resolved spectroscopy sequence. Magnetic Resonance Imaging 2011; 29(3): 374–9.

11. Bolliger CS, Boesch C, Kreis R. On the use of Cramer-Rao minimum variance bounds for the design of magnetic resonance spectroscopy experiments. NeuroImage 2013; 83: 1031–40.

12. Brix MK, Ersland L, Hugdahl K, Dwyer GE, Gruner R, Noeske R, et al. Within- and between-session reproducibility of GABA measurements with MR spectroscopy. Journal of magnetic resonance imaging : JMRI 2017; 46(2): 421–30.

13. Cai K, Nanga RP, Lamprou L, Schinstine C, Elliott M, Hariharan H, et al. The impact of gabapentin administration on brain GABA and glutamate concentrations: a 7T (1)H-MRS study. Neuropsychopharmacology : official publication of the American College of Neuropsychopharmacology 2012; 37(13): 2764–71.

14. Chen M, Liao C, Chen S, Ding Q, Zhu D, Liu H, et al. Uncertainty assessment of gamma-aminobutyric acid concentration of different brain regions in individual and group using residual bootstrap analysis. Medical & biological engineering & computing 2017; 55(6): 1051–9.

15. Choi IY, Andronesi OC, Barker P, Bogner W, Edden RAE, Kaiser LG, et al. Spectral editing in 1 H magnetic resonance spectroscopy: Experts’ consensus recommendations. NMR in Biomedicine 2021; 34(5).

16. Cleve M, Gussew A, Wagner G, Bar KJ, Reichenbach JR. Assessment of intra-and inter-regional interrelations between GABA+, Glx and BOLD during pain perception in the human brain - A combined (1)H fMRS and fMRI study. Neuroscience 2017; 365:125–36.

17. Coghlan S, Horder J, Inkster B, Mendez MA, Murphy DG, Nutt DJ. GABA system dysfunction in autism and related disorders: from synapse to symptoms. Neuroscience & Biobehavioral Reviews 2012; 36(9): 2044–55.

18. Cudalbu Cea. Contribution of macromolecules to magnetic resonane spectra: Experts’ consensus recommendations. NMR Biomed 2020.

19. De Bondt T, De Belder F, Vanhevel F, Jacquemyn Y, Parizel PM. Prefrontal GABA concentration changes in women-Influence of menstrual cycle phase, hormonal contraceptive use, and correlation with premenstrual symptoms. Brain research 2015; 1597: 129–38.

20. de Graaf RA. In Vivo NMR Spectroscopy: Principles and Techniques: Wiley; 2019. de Jonge JC, Vinkers CH, Hulshoff Pol HE, Marsman A. GABAergic Mechanisms in Schizophrenia: Linking Postmortem and In Vivo Studies. Frontiers in Psychiatry 2017; 8(118).

21. Deelchand DK, Kantarci K, Oz G. Improved localization, spectral quality, and repeatability with advanced MRS methodology in the clinical setting. Magnetic resonance in medicine 2018; 79(3): 1241–50.

22. Deelchand DK, Marjańska M, Henry P-G, Terpstra M. MEGA-PRESS of GABA+: Influences of acquisition parameters. NMR in Biomedicine 2019; n/a(n/a): e4199.

23. Dhamala E, Abdelkefi I, Nguyen M, Hennessy TJ, Nadeau H, Near J. Validation of in vivo MRS measures of metabolite concentrations in the human brain. NMR in biomedicine 2019; 32(3): e4058.

24. Duncan NW, Zhang J, Northoff G, Weng X. Investigating GABA concentrations measured with macromolecule suppressed and unsuppressed MEGA-PRESS MR spectroscopy and their relationship with BOLD responses in the occipital cortex. Journal of magnetic resonance imaging : JMRI 2019; 50(4): 1285–94.

25. Edden RA, Barker PB. Spatial effects in the detection of gamma-aminobutyric acid: improved sensitivity at high fields using inner volume saturation. Magnetic resonance in medicine 2007; 58(6): 1276–82.

26. Edden RA, Oeltzschner G, Harris AD, Puts NA, Chan KL, Boer VO, et al. Prospective frequency correction for macromolecule-suppressed GABA editing at 3T. Journal of Magnetic Resonance Imaging 2016; 44(6): 1474–82.

27. Edden RA, Puts NA, Harris AD, Barker PB, Evans CJ. Gannet: A batch-processing tool for the quantitative analysis of gamma-aminobutyric acid-edited MR spectroscopy spectra. Journal of magnetic resonance imaging : JMRI 2014; 40(6): 1445–52.

28. Edden RAE, Puts NAJ, Barker PB. Macromolecule-suppressed GABA-edited magnetic resonance spectroscopy at 3T. Magnetic resonance in medicine 2012; 68(3): 657–61.

29. Enna S, McCarson KE. The role of GABA in the mediation and perception of pain. Advances in pharmacology 2006; 54: 1–27.

30. Epperson CN, Haga K, Mason GF, Sellers E, Gueorguieva R, Zhang W, et al. Cortical gamma-aminobutyric acid levels across the menstrual cycle in healthy women and those with premenstrual dysphoric disorder: a proton magnetic resonance spectroscopy study. Archives of general psychiatry 2002; 59(9): 851–8.

31. Epperson CN, O’Malley S, Czarkowski KA, Gueorguieva R, Jatlow P, Sanacora G, et al. Sex, GABA, and nicotine: the impact of smoking on cortical GABA levels across the menstrual cycle as measured with proton magnetic resonance spectroscopy. Biological psychiatry 2005; 57(1): 44–8.

32. Ernst T, Chang L. Elimination of artifacts in short echo time H MR spectroscopy of the frontal lobe. Magnetic resonance in medicine 1996; 36(3): 462–8.

33. Gao F, Edden RA, Li M, Puts NA, Wang G, Liu C, et al. Edited magnetic resonance spectroscopy detects an age-related decline in brain GABA levels. Neuroimage 2013; 78: 75–82.

34. Gasbarri A, Pompili A. 3 - The Role of GABA in Memory Processes. In: Meneses A, editor. Identification of Neural Markers Accompanying Memory. San Diego: Elsevier; 2014. p. 47–62.

35. Gasparovic C, Chen H, Mullins PG. Errors in 1 H-MRS estimates of brain metabolite concentrations caused by failing to take into account tissue-specific signal relaxation. NMR in Biomedicine 2018; 31(6): e3914.

36. Geramita M, van der Veen JW, Barnett AS, Savostyanova AA, Shen J, Weinberger DR, et al. Reproducibility of prefrontal γ-aminobutyric acid measurements with J-edited spectroscopy. NMR in Biomedicine 2011; 24(9): 1089–98.

37. Godfrey KE, Gardner AC, Kwon S, Chea W, Muthukumaraswamy SD. Differences in excitatory and inhibitory neurotransmitter levels between depressed patients and healthy controls: a systematic review and meta-analysis. Journal of psychiatric research 2018; 105: 33–44.

38. Greenhouse I, Noah S, Maddock RJ, Ivry RB. Individual differences in GABA content are reliable but are not uniform across the human cortex. Neuroimage 2016; 139: 1–7.

39. Grewal M, Dabas A, Saharan S, Barker PB, Edden RA, Mandal PK. GABA quantitation using MEGA-PRESS: Regional and hemispheric differences. Journal of magnetic resonance imaging : JMRI 2016; 44(6): 1619–23.

40. Gruetter R, Tkáč I. Field mapping without reference scan using asymmetric echo-planar techniques. Magnetic Resonance in Medicine: An Official Journal of the International Society for Magnetic Resonance in Medicine 2000; 43(2): 319–23.

41. Gu M, Hurd R, Noeske R, Baltusis L, Hancock R, Sacchet MD, et al. GABA editing with macromolecule suppression using an improved MEGA-SPECIAL sequence. Magnetic resonance in medicine 2018; 79(1): 41–7.

42. Hall EL, Stephenson MC, Price D, Morris PG. Methodology for improved detection of low concentration metabolites in MRS: Optimised combination of signals from multi-element coil arrays. NeuroImage 2014; 86: 35–42.

43. Harada M, Kubo H, Nose A, Nishitani H, Matsuda T. Measurement of variation in the human cerebral GABA level by in vivo MEGA-editing proton MR spectroscopy using a clinical 3 T instrument and its dependence on brain region and the female menstrual cycle. Human Brain Mapping 2011; 32(5): 828–33.

44. Harris AD, Glaubitz B, Near J, John Evans C, Puts NA, Schmidt-Wilcke T, et al. Impact of frequency drift on gamma-aminobutyric acid-edited MR spectroscopy. Magnetic resonance in medicine 2014; 72(4): 941–8.

45. Harris AD, Puts NA, Barker PB, Edden RA. Spectral-editing measurements of GABA in the human brain with and without macromolecule suppression. Magnetic resonance in medicine 2015a; 74(6): 1523–9.

46. Harris AD, Puts NA, Edden RA. Tissue correction for GABA-edited MRS: Considerations of voxel composition, tissue segmentation, and tissue relaxations. Journal of Magnetic Resonance Imaging 2015b; 42(5): 1431–40.

47. Harris AD, Puts NAJ, Anderson BA, Yantis S, Pekar JJ, Barker PB, et al. Multi-Regional Investigation of the Relationship between Functional MRI Blood Oxygenation Level Dependent (BOLD) Activation and GABA Concentration. PLOS ONE 2015c; 10(2): e0117531.

48. Harris AD, Saleh MG, Edden RA. Edited (1) H magnetic resonance spectroscopy in vivo: Methods and metabolites. Magnetic resonance in medicine 2017; 77(4): 1377–89.

49. Hasson F, Keeney S, McKenna H. Research guidelines for the Delphi survey technique. Journal of Advanced Nursing 2000; 32(4): 1008–15.

50. Henry P-G, Dautry C, Hantraye P, Bloch G. Brain GABA editing without macromolecule contamination. Magnetic resonance in medicine 2001; 45(3): 517–20.

51. Juchem C, Cudalbu C, de Graaf RA, Gruetter R, Henning A, Hetherington HP, et al. B0 shimming for in vivo magnetic resonance spectroscopy: Experts’ consensus recommendations. NMR Biomed 2021; 34(5): e4350.

52. Juchem C, de Graaf RA. B0 magnetic field homogeneity and shimming for in vivo magnetic resonance spectroscopy. Anal Biochem 2017; 529: 17–29.

53. Keltner JR, Wald LL, Christensen JD, Maas LC, Moore CM, Cohen BM, et al. A technique for detecting GABA in the human brain with PRESS localization and optimized refocusing spectral editing radiofrequency pulses. Magnetic resonance in medicine 1996; 36(3): 458–61.

54. Kolasinski J, Hinson EL, Divanbeighi Zand AP, Rizov A, Emir UE, Stagg CJ. The dynamics of cortical GABA in human motor learning. J Physiol 2019; 597(1): 271–82.

55. Kreis R. Issues of spectral quality in clinical 1H-magnetic resonance spectroscopy and a gallery of artifacts. NMR Biomed 2004; 17(6): 361–81.

56. Kreis R. The trouble with quality filtering based on relative Cramér-Rao lower bounds. Magnetic resonance in medicine 2016; 75(1): 15–8.

57. Kreis R, Boer V, Choi I-Y, Cudalbu C, de Graaf RA, Gasparovic C, et al. Terminology and concepts for the characterization of in vivo MR spectroscopy methods and MR spectra: Background and experts’ consensus recommendations. NMR in biomedicine 2021; 34(5): e4347.

58. Licata SC, Jensen JE, Penetar DM, Prescot AP, Lukas SE, Renshaw PF. A therapeutic dose of zolpidem reduces thalamic GABA in healthy volunteers: a proton MRS study at 4 T. Psychopharmacology 2009; 203(4): 819–29.

59. Lin A, Andronesi O, Bogner W, Choi I-Y, Coello E, Cudalbu C, et al. Minimum Reporting Standards for in vivo Magnetic Resonance Spectroscopy (MRSinMRS): Experts’ consensus recommendations. NMR in Biomedicine 2021; 34(5): e4484.

60. Long Z, Dyke JP, Ma R, Huang CC, Louis ED, Dydak U. Reproducibility and effect of tissue composition on cerebellar gamma-aminobutyric acid (GABA) MRS in an elderly population. NMR in biomedicine 2015; 28(10): 1315–23.

61. Maes C, Hermans L, Pauwels L, Chalavi S, Leunissen I, Levin O, et al. Age-related differences in GABA levels are driven by bulk tissue changes. Human Brain Mapping 2018; 39(9): 3652–62.

62. Marenco S, Meyer C, van der Veen JW, Zhang Y, Kelly R, Shen J, et al. Role of gamma-amino-butyric acid in the dorsal anterior cingulate in age-associated changes in cognition. Neuropsychopharmacology : official publication of the American College of Neuropsychopharmacology 2018; 43(11): 2285–91.

63. Marotta R, Risoleo MC, Messina G, Parisi L, Carotenuto M, Vetri L, et al. The neurochemistry of autism. Brain sciences 2020; 10(3): 163.

64. Mescher M, Merkle H, Kirsch J, Garwood M, Gruetter R. Simultaneous in vivo spectral editing and water suppression. NMR in biomedicine 1998; 11(6): 266–72.

65. Mikkelsen M, Barker PB, Bhattacharyya PK, Brix MK, Buur PF, Cecil KM, et al. Big GABA: Edited MR spectroscopy at 24 research sites. Neuroimage 2017; 159: 32–45.

66. Mikkelsen M, Loo RS, Puts NAJ, Edden RAE, Harris AD. Designing GABA-edited magnetic resonance spectroscopy studies: Considerations of scan duration, signal-to-noise ratio and sample size. Journal of neuroscience methods 2018; 303: 86–94.

67. Mikkelsen M, Rimbault DL, Barker PB, Bhattacharyya PK, Brix MK, Buur PF, et al. Big GABA II: Water-referenced edited MR spectroscopy at 25 research sites. Neuroimage 2019; 191: 537–48.

68. Mikkelsen M, Singh KD, Brealy JA, Linden DE, Evans CJ. Quantification of gamma-aminobutyric acid (GABA) in 1-H MRS volumes composed heterogeneously of grey and white matter. NMR in Biomedicine 2016a; 29(11): 1644–55.

69. Mikkelsen M, Singh KD, Sumner P, Evans CJ. Comparison of the repeatability of GABA-edited magnetic resonance spectroscopy with and without macromolecule suppression. Magnetic resonance in medicine 2016b; 75: 946–53.

70. Miller KA, Collada B, Tolliver D, Audi Z, Cohen A, Michelson C, et al. Using the Modified Delphi Method to Develop a Tool to Assess Pediatric Residents Supervising on Inpatient Rounds. Academic Pediatrics 2020; 20(1): 89–96.

71. Mullins PG, McGonigle DJ, O’Gorman RL, Puts NA, Vidyasagar R, Evans CJ, et al. Current practice in the use of MEGA-PRESS spectroscopy for the detection of GABA. Neuroimage 2014; 86: 43–52.

72. Myers JF, Evans CJ, Kalk NJ, Edden RA, Lingford-Hughes AR. Measurement of GABA using J-difference edited 1H-MRS following modulation of synaptic GABA concentration with tiagabine. Synapse (New York, NY) 2014; 68(8): 355–62.

73. Near J, Edden R, Evans CJ, Paquin R, Harris A, Jezzard P. Frequency and phase drift correction of magnetic resonance spectroscopy data by spectral registration in the time domain. Magnetic resonance in medicine 2015; 73(1): 44–50.

74. Near J, Harris AD, Juchem C, Kreis R, Marjańska M, Öz G, et al. Preprocessing, analysis and quantification in single-voxel magnetic resonance spectroscopy: experts’ consensus recommendations. NMR in biomedicine 2020.

75. NHMRC. A guide to the development, implementation adn evaluation fo clincial practice guidelines. Canberra: National Health and Medical Research Council; 1999.

76. NHMRC. NHMRC levels of evidence and grades for recommendations for developers of guidelines. Canberra: NHMRC; 2009.

77. NHMRC. Guidelines for Guidelines Handbook. 2021 [cited 2020 Jan]; Available from: www.nhmrc.gov.au/guidelinesforguidelines

78. O’Gorman RL, Michels L, Edden RA, Murdoch JB, Martin E. In vivo detection of GABA and glutamate with MEGA-PRESS: reproducibility and gender effects. Journal of magnetic resonance imaging : JMRI 2011; 33(5): 1262–7.

79. Oeltzschner G, Butz M, Baumgarten TJ, Hoogenboom N, Wittsack HJJr, Schnitzler A, et al. Use of quantitative brain water imaging as concentration reference for J-edited MR spectroscopy of GABA. Metabolic brain disease 2016; 30: 1429–38.

80. Oeltzschner G, Chan KL, Saleh MG, Mikkelsen M, Puts NA, Edden RAE. Hadamard editing of glutathione and macromolecule-suppressed GABA. NMR in biomedicine 2018a; 31(1).

81. Oeltzschner G, Zöllner HJ, Jonuscheit M, Lanzman RS, Schnitzler A, Wittsack H-J. J-difference-edited MRS measures of γ-aminobutyric acid before and after acute caffeine administration. Magnetic resonance in medicine 2018b.

82. Öz G, Deelchand DK, Wijnen JP, Mlynárik V, Xin L, Mekle R, et al. Advanced single voxel 1H magnetic resonance spectroscopy techniques in humans: Experts’ consensus recommendations. NMR in biomedicine 2020; n/a(n/a): e4236.

83. Park YW, Deelchand DK, Joers JM, Hanna B, Berrington A, Gillen JS, et al. AutoVOI: real-time automatic prescription of volume-of-interest for single voxel spectroscopy. Magnetic resonance in medicine 2018; 80(5): 1787–98.

84. Peek AL, Rebbeck T, Puts NA, Watson J, Aguila ME, Leaver AM. Brain GABA and glutamate levels across pain conditions: A systematic literature review and meta-analysis of 1H-MRS studies using the MRS-Q quality assessment tool. Neuroimage 2020: 116532.

85. Petroff OA, Rothman DL, Behar KL, Collins TL, Mattson RH. Human brain GABA levels rise rapidly after initiation of vigabatrin therapy. Neurology 1996a; 47(6): 1567–71.

86. Petroff OA, Rothman DL, Behar KL, Lamoureux D, Mattson RH. The effect of gabapentin on brain gamma-aminobutyric acid in patients with epilepsy. Annals of Neurology 1996b; 39(1): 95–9.

87. Petty F. Plasma concentrations of gamma-aminobutyric acid (GABA) and mood disorders: a blood test for manic depressive disease? Clin Chem 1994; 40(2): 296–302.

88. Porges EC, Jensen G, Foster B, Edden RAE, Puts NAJ. The trajectory of cortical GABA levels across the lifespan: An individual participant data meta-analysis of edited MRS studies. bioRxiv 2020: 2020.07.23.218792.

89. Porges EC, Woods AJ, Edden RA, Puts NA, Harris AD, Chen H, et al. Frontal Gamma-Aminobutyric Acid Concentrations Are Associated With Cognitive Performance in Older Adults. Biological psychiatry Cognitive neuroscience and neuroimaging 2017; 2(1): 38–44.

90. Puts NA, Barker PB, Edden RA. Measuring the longitudinal relaxation time of GABA in vivo at 3 Tesla. Journal of Magnetic Resonance Imaging 2013; 37(4): 999–1003.

91. Puts NA, Edden RA. In vivo magnetic resonance spectroscopy of GABA: a methodological review. Prog Nucl Magn Reson Spectrosc 2012; 60: 29–41.

92. Puts NAJ, Heba S, Harris AD, Evans CJ, McGonigle DJ, Tegenthoff M, et al. GABA Levels in Left and Right Sensorimotor Cortex Correlate across Individuals. Biomedicines 2018; 6(3): 24.

93. Puts NAJ, Wodka EL, Harris AD, Crocetti D, Tommerdahl M, Mostofsky SH, et al. Reduced GABA and altered somatosensory function in children with autism spectrum disorder. Autism Research 2017; 10(4): 608–19.

94. Rothman DL, Petroff OA, Behar KL, Mattson RH. Localized 1H NMR measurements of gamma-aminobutyric acid in human brain in vivo. Proceedings of the National Academy of Sciences of the United States of America 1993; 90(12): 5662–6.

95. Saleh MG, Near J, Alhamud A, Robertson F, van der Kouwe AJ, Meintjes EM. Reproducibility of macromolecule suppressed GABA measurement using motion and shim navigated MEGA-SPECIAL with LCModel, jMRUI and GANNET. Magma 2016a; 29(6): 863–74.

96. Saleh MG, Near J, Alhamud A, Van Der Kouwe AJW, Meintjes EM. Effects of tissue and gender on macromolecule suppressed gamma-aminobutyric acid. International Journal of Imaging Systems and Technology 2017; 27(2): 144–52.

97. Saleh MG, Oeltzschner G, Chan KL, Puts NAJ, Mikkelsen M, Schär M, et al. Simultaneous edited MRS of GABA and glutathione. Neuroimage 2016b; 142: 576–82.

98. Saleh MG, Rimbault D, Mikkelsen M, Oeltzschner G, Wang AM, Jiang D, et al. Multi-vendor standardized sequence for edited magnetic resonance spectroscopy. Neuroimage 2019; 189: 425–31.

99. Sanaei Nezhad F, Anton A, Michou E, Jung J, Parkes LM, Williams SR. Quantification of GABA, glutamate and glutamine in a single measurement at 3 T using GABA-edited MEGA-PRESS. NMR in biomedicine 2018; 31(1).

100. Schulte MHJ, Kaag AM, Wiers RW, Schmaal L, van den Brink W, Reneman L, et al. Prefrontal Glx and GABA concentrations and impulsivity in cigarette smokers and smoking polysubstance users. Drug and alcohol dependence 2017; 179: 117–23.

101. Schur RR, Draisma LW, Wijnen JP, Boks MP, Koevoets MG, Joels M, et al. Brain GABA levels across psychiatric disorders: A systematic literature review and meta-analysis of (1) H-MRS studies. Human Brain Mapping 2016; 37(9): 3337–52.

102. Schür RR, Draisma LW, Wijnen JP, Boks MP, Koevoets MG, Joëls M, et al. Brain GABA levels across psychiatric disorders: A systematic literature review and meta-analysis of 1H-MRS studies. Human brain mapping 2016; 37(9): 3337–52.

103. Shungu DC, Mao X, Gonzales R, Soones TN, Dyke JP, van der Veen JW, et al. Brain gamma-aminobutyric acid (GABA) detection in vivo with the J-editing (1) H MRS technique: a comprehensive methodological evaluation of sensitivity enhancement, macromolecule contamination and test-retest reliability. NMR in Biomedicine 2016; 29(7): 932–42.

104. Simmonite M, Carp J, Foerster BR, Ossher L, Petrou M, Weissman DH, et al. Age-Related Declines in Occipital GABA are Associated with Reduced Fluid Processing Ability. Academic radiology 2019; 26(8): 1053–61.

105. Sun W, Zhou Q, Ba X, Feng X, Hu X, Cheng X, et al. Oxytocin Relieves Neuropathic Pain Through GABA Release and Presynaptic TRPV1 Inhibition in Spinal Cord. Frontiers in Molecular Neuroscience 2018; 11(248).

106. Tapper S, Tisell A, Helms G, Lundberg P. Retrospective artifact elimination in MEGA-PRESS using a correlation approach. Magnetic resonance in medicine 2019; 81(4): 2223–37.

107. The ADAPTE Collaboration. The ADAPTE Process: Resource Toolkit for Guidline Adaptation. 2009 [cited 2019 01/07]; Version 2.0:[Available from: http://www.g-i-n.net

108. van der Veen JW, Marenco S, Berman KF, Shen J. Retrospective correction of frequency drift in spectral editing: The GABA editing example. NMR in Biomedicine 2017: e3725.

109. van der Veen JW, Shen J. Regional difference in GABA levels between medial prefrontal and occipital cortices. Journal of Magnetic Resonance Imaging 2013; 38(3): 745–50.

110. Vengeliene V, Bilbao A, Molander A, Spanagel R. Neuropharmacology of alcohol addiction. British journal of pharmacology 2008; 154(2): 299–315.

111. Waddell KW, Zanjanipour P, Pradhan S, Xu L, Welch EB, Joers JM, et al. Anterior cingulate and cerebellar GABA and Glu correlations measured by 1H J-difference spectroscopy. Magnetic Resonance Imaging 2011; 29(1): 19–24.

112. Wiegers EC, Philips BWJ, Heerschap A, van der Graaf M. Automatic frequency and phase alignment of in vivo J-difference-edited MR spectra by frequency domain correlation. Magma 2017; 30(6): 537–44.

113. Wilson M, Andronesi O, Barker PB, Bartha R, Bizzi A, Bolan PJ, et al. Methodological consensus on clinical proton MRS of the brain: Review and recommendations. Magnetic resonance in medicine 2019; 82: 527–50.

114. Wood ET, Cummings KK, Jung J, Patterson G, Okada N, Guo J, et al. Sensory over-responsivity is related to GABAergic inhibition in thalamocortical circuits. Transl Psychiatry 2021; 11(1).

115. Zacharopoulos G, Sella F, Cohen Kadosh K, Hartwright C, Emir U, Cohen Kadosh R. Predicting learning and achievement using GABA and glutamate concentrations in human development. PLOS Biology 2021; 19(7): e3001325.

