## Supplement 1 for "A Comprehensive Guide to MEGA-PRESS for GABA Measurement"

**Review Methods:**

**Database:**

Search Ovid MEDLINE, Embase and PubMed

**Search Strategy**

((mega press OR gaba OR gamma-amino but* OR spectral editing)) AND (MRS OR magnetic resonance spectro* OR "magnetic resonance methods") AND (white paper OR consensus OR acquisition parameters OR gradients OR water suppression OR editing pulses OR voxel OR methods OR cramer lower bounds OR CRLB OR full width half maximum OR FWHM OR shim OR frequency drift OR phase drift OR quality OR reporting)

**Inclusion/ Exclusion**

P: Include: humans, phantom or computer simulations

Exclude: animals

I: Magnetic resonance spectroscopy, single voxel, spectral-edited, brain

Exclude: 2D techniques, MRSI

O: Contribute to the six identified domains

Exclude: studies reporting outcomes of clinical cohorts, except those related to medications

S: White papers, consensus, methods papers, systematic or quasi-systematic reviews, peer reviewed journal

Exclude conference proceedings, clinical commentaries, editorials, narrative reviews

**Study selection**

Studies were screened using a two-stage approach. Firstly, two reviewers (AP, GO) independently screened the titles and abstracts to identify papers potentially suitable for inclusion. Secondly, the same two reviewers independently reviewed the full text to determine the final eligible papers for each of the six domains. Disagreements were discussed and resolved through consultation of a third reviewer (NP). Rationale for exclusion was documented and duplicates were removed. The MRS sub-committee reviewed the results of the search and identified any missing papers.

**Data Extraction**

Data was independently extracted by two reviewers with a special interest and experience in GABA MRS into a predesigned data extraction sheet. Inconsistencies or disagreements were discussed for resolution. Data was then synthesised using qualitative analysis.

**Parameters not addressed in publications specifically concerned with spectral editing (grey literature search)**

In cases where parameters were not addressed in publications specifically concerned with spectral editing, literature concerning conventional (un-edited) single voxel spectroscopy, e.g. guidelines, white papers, consensus documents and other publications were drawn upon.
