## Supplementary figures and images for "A Comprehensive Guide to MEGA-PRESS for GABA Measurement"

### Supplement 2

#### SUPPLEMENT 2

##### PRISMA Flow Diagram

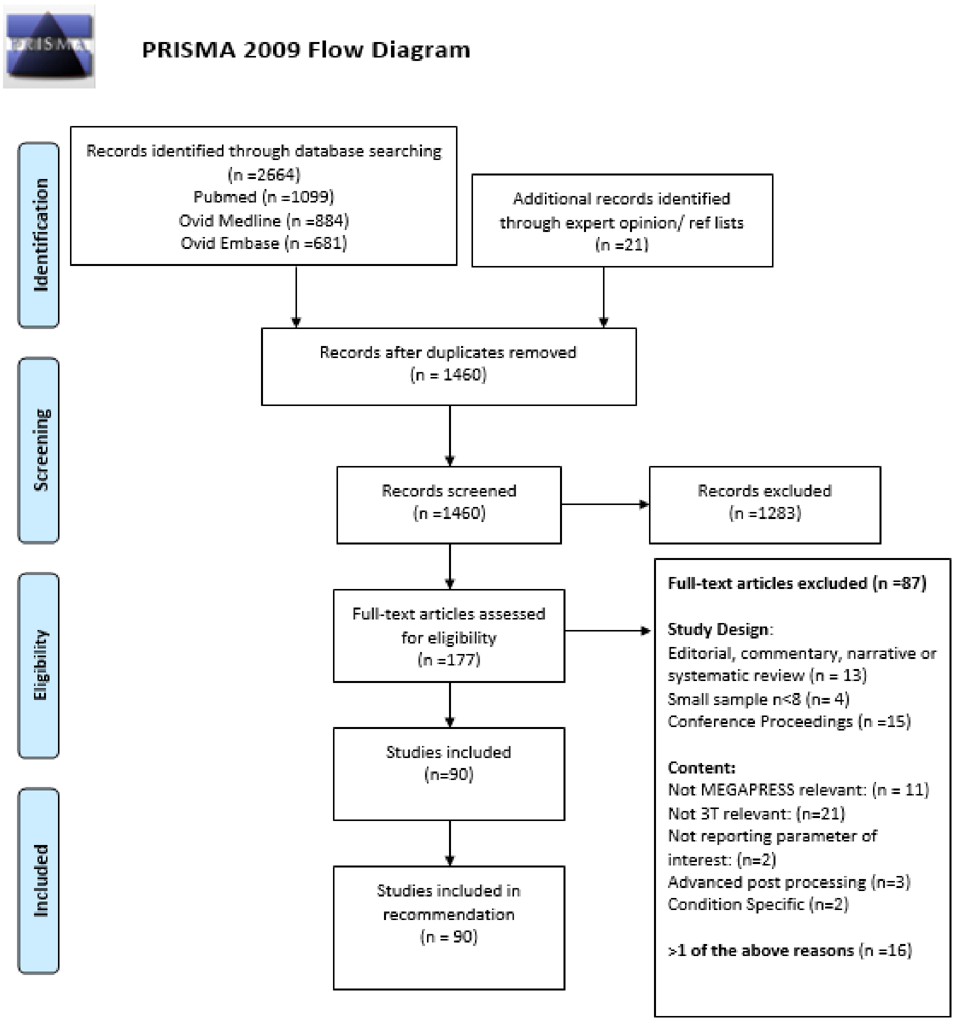

### Supplement 5: Infographic

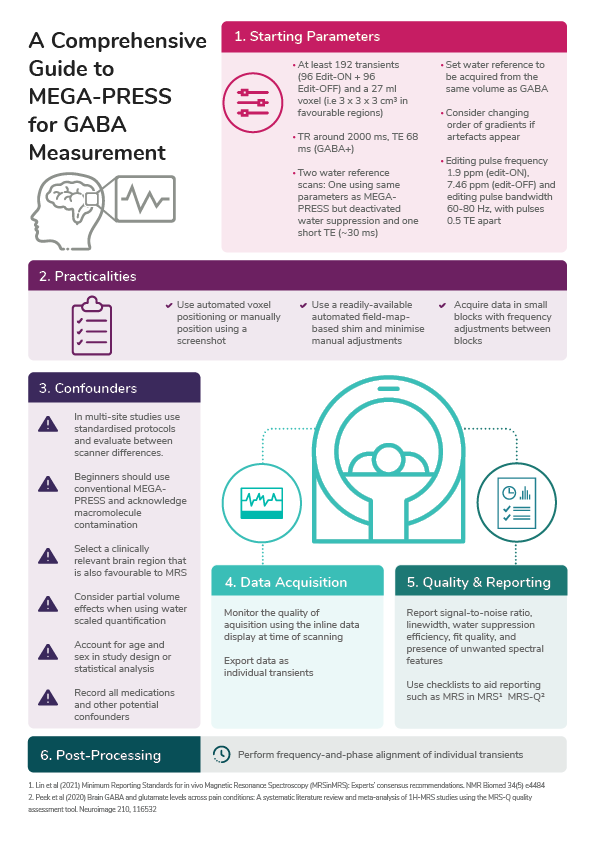
