## Supplement 3 for "A Comprehensive Guide to MEGA-PRESS for GABA Measurement"

**Table 1: Summary of papers used to inform the guideline**

| | | **Papers with Recommendations** | **Study Design** | **Level of Evidence** |
|---|---|---|---|---|
| **Parameters** | **Signal to noise** | | | |
| | - Transients | Mullins *et al.*, 2014 | Cons | 1 |
| | | Mikkelsen *et al.*, 2017 | MSS | 2 |
| | | Mikkelsen *et al.*, 2019 | MSS | 2 |
| | | Bhattacharyya *et al.*, 2007 | M | 3 |
| | | Harris *et al.*, 2014 | M | 3 |
| | | Brix *et al.*, 2017 | M | 3 |
| | | Mikkelsen *et al.*, 2018 | M | 3 |
| | | Sanaei Nezhad *et al.*, 2018 | M | 3 |
| | *Reporting of* | Lin *et al.,* 2020 | Cons | 1 |
| | | Peek *et al.,* 2020 | SR | 2 |
| | - Voxel Volume | Mullins *et al.*, 2014 | Cons | 1 |
| | | de Graaf, 2019 | Sem | 1 |
| | | Mikkelsen *et al.*, 2017 | MSS | 2 |
| | | Mikkelsen *et al.*, 2019 | MSS | 2 |
| | | Bai *et al.*, 2015 | M | 3 |
| | | Bergmann *et al.*, 2016 | M | 3 |
| | | Chen *et al.*, 2017 | M | 3 |
| | TR | Wilson *et al.*, 2019 | Cons | 1 |
| | | Mikkelsen *et al.*, 2017 | MSS | 2 |
| | | Mikkelsen *et al.*, 2019 | MSS | 2 |
| | | Puts *et al.*, 2013; | M | 3 |
| | | Deelchand *et al.*, 2019 | M | 3 |
| | TE | Cudalbu *et al.,* 2020 | Cons | 1 |
| | | Mullins *et al.*, 2014; | Cons | 1 |
| | | Wilson *et al.*, 2019 | Cons | 1 |
| | | Mescher *et al.*, 1998 | SemP | 1 |
| | | Puts *et al.*, 2012 | SR | 2 |
| | | Mikkelsen *et al.*, 2017 | MSS | 2 |
| | | Mikkelsen *et al.*, 2019 | MSS | 3 |
| | | Harris *et al.*, 2015a; | M | 3 |
| | | Edden *et al.*, 2016; | M | 3 |
| | | Deelchand *et al.*, 2019 | M | 1 |
| | Water Reference | Mullins *et al.*, 2014; | Cons | 1 |
| | | Wilson *et al.*, 2019; | Cons | 1 |
| | | Öz *et al.*, 2020 | Cons | 1 |
| | | Near et al.*, 2021* | Cons | 1 |
| | | de Graaf, 2019 | Sem T | 1 |
| | | Hall *et al.*, 2014; | M | 3 |

| | | | | |
|---|---|---|---|---|
| | | Oeltzschner *et al.*, 2016 | M | 3 |
| | Slice-selection frequency of water reference | Mikkelsen *et al.*, 2017 | MSS | 2 |
| | | Mikkelsen *et al.*, 2019 | MSS | 2 |
| | | Deelchand *et al.*, 2019 | M | 3 |
| | Slice-selective gradients | Ernst and Chang, 1996 | M | 3 |
| | Editing pulse specifications | Mullins *et al.*, 2014 | Cons | 1 |
| | | Mikkelsen *et al.*, 2017 | MSS | 2 |
| | | Mikkelsen *et al.*, 2019 | MSS | 2 |
| | | Saleh *et al., 2019* | MSS | 2 |
| | | Keltner *et al.*, 1996 | M | 3 |
| | | Mescher *et al.*, 1998 | M | 3 |
| | | Henry *et al.*, 2001 | M | 3 |
| | | Edden *et al.*, 2016 | M | 3 |
| | | Deelchand *et al., 2019* | M | 3 |
| **Practicalities** | Voxel Position | Öz *et al.*, 2020 | Cons | 1 |
| | | Bai *et al.*, 2017 | M | 3 |
| | | Chen *et al.*, 2017 | M | 3 |
| | | Park *et al.,* 2018 | M | 3 |
| | | Kreis, 2004 | Nar | 4 |
| | Shimming | Juchem *et al. 2020* | Cons | 1 |
| | | Wilson *et al.*, 2019 | Cons | 1 |
| | | Öz *et al.*, 2020 | Cons | 1 |
| | | Juchem et al 2017 | SR | 2 |
| | | Grewal et al 2016 | M | 3 |
| | | Saleh *et al.*, 2016 | M | 3 |
| | | Deelchand *et al.*, 2018; | M | 3 |
| | | Sanaei Nezhad *et al.*, 2018 | M | 3 |
| | Order of scans and field drift | Öz *et al.*, 2020 | Cons | 1 |
| | | Andronesi *et al.*, 2020 | Cons | 1 |
| | | Choi *et al.,* 2021 | Cons | 1 |
| | | Cudalbu *et al.,* 2020 | Cons | 1 |
| | | Mikkelsen *et al.*, 2017; | MSS | 2 |
| | | Harris *et al.*, 2014a | M | 3 |
| | | Edden *et al.*, 2016 | M | 3 |
| **Confounders** | Scanner site and vendor | Mikkelsen *et al.*, 2017; | MSS | 2 |
| | | Mikkelsen *et al.*, 2019; | MSS | 2 |
| | | Saleh *et al.*, 2019 | MSS | 2 |
| | Macromolecules | Mullins *et al.,* 2014 | Cons | 1 |
| | | Cudalbu *et al.,* 2020 | Cons | 1 |
| | | Choi *et al.,* 2021 | Cons | 1 |
| | | Henry *et al.*, 2001; | M | 3 |
| | | Harris *et al.*, 2015a; | M | 3 |
| | | Edden *et al.*, 2016; | M | 3 |

| | | | |
|---|---|---|---|
| | Mikkelsen *et al.*, 2016b; | M | 3 |
| | Shungu *et al.*, 2016; | M | 3 |
| | Gu *et al.*, 2018; | M | 3 |
| | Oeltzschner *et al.*, 2018a; | M | 3 |
| | Duncan *et al.*, 2019 | M | 3 |
| Region | Puts and Edden, 2012 | SR | 1[T] |
| | Harada *et al.*, 2011; | M | 4[T] |
| | Waddell *et al.*, 2011; | M | 4[T] |
| | Gao *et al.*, 2013; | M | 4[T] |
| | van der Veen 2013; | M | 4[T] |
| | Harris *et al.*, 2015c; | M | 4[T] |
| | Long *et al.*, 2015; | M | 4[T] |
| | Greenhouse *et al.*, 2016; | M | 4[T] |
| | Grewal *et al.*, 2016a; | M | 4[T] |
| | Brix *et al.*, 2017; | M | 4[T] |
| | Chen *et al.*, 2017b; | M | 4[T] |
| | Porges *et al.*, 2017a; | M | 4[T] |
| | Puts *et al.*, 2018; | M | 4[T] |
| | Dhamala *et al.*, 2019 | M | 4[T] |
| Tissue | Mullins *et al.*, 2014 | Cons | 1 |
| Composition | Choi *et al.*, 2006; | M | 3 |
| | Bhattacharyya *et al.*, | M | 3 |
| | 2011; | M | 3 |
| | Geramita *et al., 2011;* | M | 3 |
| | Harris *et al.*, 2015b; | M | 3 |
| | Mikkelsen *et al.*, | M | 3 |
| | 2016a; | M | 3 |
| | Porges *et al.*, 2017b; | M | 3 |
| | Gasparovic *et al.,* 2018 | | |
| Age | Porges *et al.*, 2020; | SR | 1[T] |
| | Aufhaus *et al.*, 2013a; | M | 3[T] |
| | Gao *et al.*, 2013; | M | 3[T] |
| | Porges *et al.*, 2017a; | M | 3[T] |
| | Maes *et al.*, 2018; | M | 3[T] |
| | Marenco *et al.*, 2018; | M | 3[T] |
| | Simmonite *et al.*, 2019 | M | 3[T] |
| Sex | Aufhaus *et al.*, 2013a; | M | 3[T] |
| | Gao *et al.*, 2013; | M | 3[T] |
| | Saleh *et al.*, 2017; | M | 3[T] |
| | O'Gorman *et al.*, 2011a | M | 4[T] |
| Medication | Puts and Edden, 2012; | SR | 1[T] |
| | Bhagwagar *et al.*, 2004; | RCT | 2[T] |
| | Rothman *et al.*, 1993; | M | 4[T] |
| | Petroff *et al.*, 1996a; | M | 3[T] |

| | | | | |
|---|---|---|---|---|
| | | Petroff *et al.*, 1996b; | M | $3^{T}$ |
| | | Licata *et al.*, 2009; | M | $4^{T}$ |
| | | Cai *et al.*, 2012; | M | $4^{T}$ |
| | | Myers *et al.*, 2014 | M | $4^{T}$ |
| | **Potential Confounders** | | | |
| | - Caffeine | Oeltzschner *et al.*, 2018b; | M | $4^{T}$ |
| | - Nicotine | Epperson *et al.*, 2005; | M | $3^{T}$ |
| | | Schulte *et al.*, 2017; | M | $3^{T}$ |
| | - Menstrual Cycle | Epperson *et al.*, 2005; | M | $3^{T}$ |
| | | De Bondt *et al.*, 2015a | M | $3^{T}$ |
| | | Epperson *et al.*, 2002; | M | $4^{T}$ |
| | | Harada *et al.*, 2011; | M | $4^{T}$ |
| **Data Acquisition** | Quality assessment during the scan | Öz *et al.*, 2020; | Cons | 1 |
| | | Choi *et al.,* 2021 | Cons | 1 |
| | Data Export | - | | |
| **Quality and Reporting** | Quality Metrics | Mullins *et al.*, 2014; | Cons | 1 |
| | | Wilson *et al.*, 2019; | Cons | 1 |
| | | Öz *et al.*, 2020; | Cons | 1 |
| | | Bolliger *et al.*, 2013; | M | 3 |
| | | Kreis, 2016; | M | 3 |
| | | Chen *et al.*, 2017b; | M | 3 |
| | | Deelchand *et al.*, 2018 | M | 3 |
| | Reporting | Lin *et al.,* 2020; | Cons | 1 |
| | | Peek *et al.*, 2020; | SR | 2 |
| | | Deelchand *et al.*, 2019 | M | 3 |
| **Post-processing** | Frequency and Phase Correction | Near *et al.*, 2020; | Cons | 1 |
| | | Choi *et al.,* 2021; | Cons | 1 |
| | | Edden and Barker, 2007; | M | 3 |
| | | | M | 3 |
| | | Edden *et al.*, 2014b; | M | 3 |
| | | Harris *et al.*, 2014c; | M | 3 |
| | | Cleve *et al.*, 2015; | M | 3 |
| | | Near *et al.*, 2015; | M | 3 |
| | | van der Veen *et al.*, 2017; | M | 3 |
| | | Wiegers *et al.*, 2017; | M | 3 |
| | | Tapper *et al.*, 2019a | | |

**Cons= consensus document; MSS= multi-site study SemT/P= seminal textbook/paper; SR= systematic review; M: methodological publication, Nar: Narrative.** Note levels of evidence documented with 'Level $X^{T}$' are assessed using a traditional hierarchy of evidence (NHMRC, 2009) rather than the modified-hierarchy of evidence (Table 1).
