## Supplement 4: Full Guideline for "A Comprehensive Guide to MEGA-PRESS for GABA Measurement"

V1.0: Last updated 12th November 2021

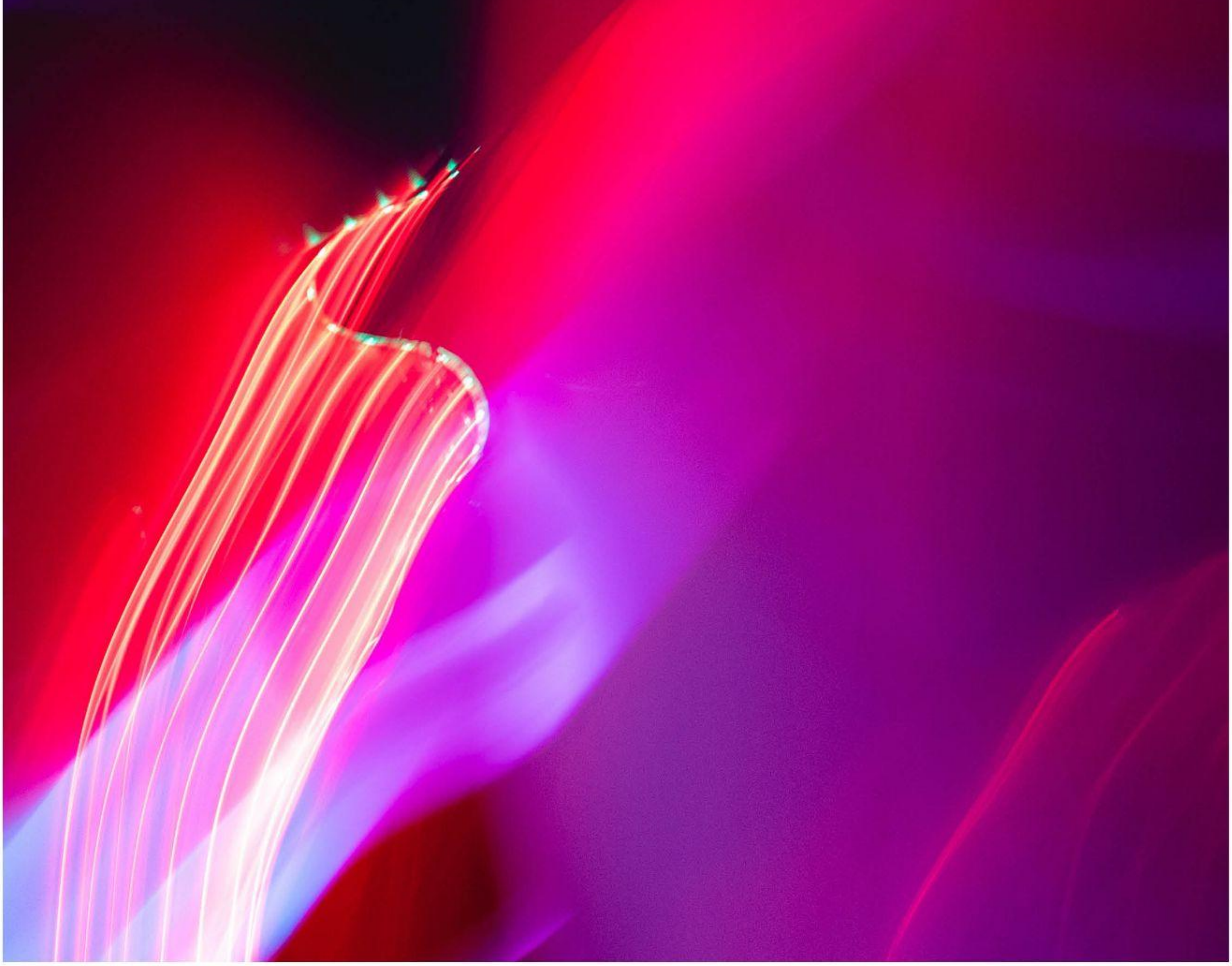

### Table of Contents


Working Group Members 6

Preface 7

1. Parameters 13

2. Practicalities 25

3. Confounders 32

4. Data Acquisition 42

5. Quality & Reporting 46

6. Post-Processing 49

References 51

Supplementary Material 54

### Guideline Working Party

**Aimie L Peek[1,2*],**

**Trudy J Rebbeck[1,2],**

**Andrew M Leaver[1],**

**Nicolaas A Puts[3,4],**

**Sheryl L Foster[1,5],**

**Kathryn M Refshauge[1],**

**Georg Oeltzschner[6,7]**

1. Faculty of Medicine and Health, University of Sydney, New South Wales Australia
2. NHMRC Centre of Research Excellence in Road Traffic Injury Recovery, Queensland, AUSTRALIA;
3. Department of Forensic and Neurodevelopmental Sciences, Sackler Institute for Translational Neurodevelopment, Institute of Psychiatry, Psychology, and Neuroscience, King's College London, UK,
4. MRC Centre for Neurodevelopmental Disorders, King's College London, UK
5. Department of Radiology, Westmead Hospital, New South Wales, Australia
6. Department of Radiology and Radiological Science, Johns Hopkins University School of Medicine, Baltimore, Maryland, USA
7. F.M. Kirby Research Center for Functional Brain Imaging, Kennedy Krieger Institute, Baltimore, Maryland, USA

### MRS Expert Panel:

**Alexander P Lin, Ashley D Harris, David J Lythgoe, Dinesh K Deelchand, Gülin Öz, In-Young Choi, Jamie Near, Kim M Cecil, Caroline D Rae, Mark Mikkelsen, Martin Wilson, Melissa Terpstra, Ovidiu Andronesi, Paul G Mullins, Peter B Barker, Richard AE Edden, Robin de Graaf, Stephen Williams, Ulrike Dydak, Uzay E Emir and Wolfgang Bogner.**

### Preface

**These guidelines aim to enable those new to the field of MEGA-PRESS to acquire high-quality data for the reliable quantification of GABA**

Gamma-aminobutyric acid (GABA) is the main inhibitory neurotransmitter of the central nervous system (CNS) and plays an important role in regulating healthy brain function[1]. Altered GABAergic function has been identified in a number of pathological conditions that affect the central nervous system such as pain[2], psychological[3,4] and neurodevelopmental disorders[5].

Magnetic Resonance Spectroscopy (MRS) is currently the only non-invasive brain imaging technique which enables the *in-vivo* measurement of GABA. The measurement of GABA using conventional MRS is challenging given its relatively low concentration in the human brain, the spectral overlap by more abundant neurometabolites and its complicated peak pattern[6]. Therefore, an edited sequence such as MEGA-PRESS (MEscher–GArwood Point RESolved Spectroscopy[7]) is required.

MEGA-PRESS is the most widely used technique for measuring GABA levels at 3T. The sequence uses J-difference editing which consists of two sub-experiments, usually acquired in an interleaved fashion. One sub-experiment applies editing pulses at a frequency of 1.9 ppm to selectively refocus the coupling evolution of the GABA signal at 3 ppm ('Edit-ON'), while the other allows the free evolution of the spin system throughout the echo time ('Edit-OFF'). Subtracting the Edit-OFF from the Edit-ON spectrum reveals a difference-edited GABA signal while removing the stronger overlapping signals from creatine-containing compounds[7]. The composite edited signal at 3ppm contains up to 50% co-edited macromolecules and is therefore commonly referred to as GABA+ (GABA+ macromolecules). Macromolecule signals can be suppressed by adding a second editing pulse at 1.5 ppm, however this experiment is significantly less stable[8].

MEGA-PRESS significantly improves the accurate detection of GABA, however, it is a technically challenging process that relies on the observation of a number of caveats and avoiding numerous pitfalls. At present the large heterogeneity of sequences and parameters used to study GABA  demonstrates the lack of standardisation within the field, resulting in variable reliability of the data[2]. The study of GABA has been met with growing interest from those with clinical backgrounds but without a background in magnetic resonance physics. This has motivated an easily translatable guideline to assist the avoidance of pitfalls and ensure the accurate detection of GABA in clinical and research populations.

### Purpose

These guidelines are intended to assist those new to the field of MEGA-PRESS to plan and implement a study to reliably measure brain GABA levels in clinical and research populations.

#### Specifically these guidelines will assist those new to the field to:

- → Select appropriate **'acquisition parameters'** depending on the brain region of interest and specific study population
- → Be aware of the impact the choice of **'acquisition parameters'** is likely to have on the spectral output
- → Be aware of specific '**practicalities'** to be considered when running a MEGA-PRESS experiment.
- → Be aware of identified and potential '**confounders'** of GABA and methods to handle these confounders.
- → Be aware of the implications associated with the methods for handling **'confounders**'.
- → Have knowledge of key aspects during **'data acquisition'**
- → Conduct appropriate **'quality assessment and reporting'** of the experiment
- → Understand the importance of frequency-and-phase correction for **'post-processing'**

#### Scope

These guidelines largely focus on study design and data acquisition to ensure steps are followed to collect high-quality data. It was not within the scope of the guideline to discuss further details of post-processing (beyond frequency-and-phase correction and the requirements for file export at the time of acquisition), modelling, or quantification of MEGA-PRESS data. Resources to assist the next stages of post-processing and quantification have been outlined in the accompanying manuscript[9]. Further it is advised that the beginner liaise with MRS experts and representatives from their scanner vendor to provide further information on vendor-specific variances highlighted in the Comprehensive Guide.

#### Development

The guideline was developed using a translation framework widely used for the development of clinical guidelines, the NHMRC framework Guidelines for Guidelines[10] and the ADAPTE toolkit[11]. Full details of the development process including search strategy of the scoping review are outlined in the accompanying manuscript[9]. In brief, this framework divides the evidence synthesis and recommendation formation into three stages: set up, adaptation and finalisation[11]. The stages are summarized in Figure 1. The key strengths of this approach include the involvement of multiple stakeholders with diverse experience and expertise, conducting a systematically delivered scoping review, the blinded quality assessment of each recommendation, and the modified-Delphi approach[12,13] used to integrate external expert peer review.

**Set up**

→ Establish working group, committee and stakeholders
→ Develop work plan

**Adaptation**

→ Plan scope and purpose of guideline
→ Devise comprehensive search and screening strategy
→ Conduct scoping review
→ Synthesise evidence from scoping review
→ Identify where existing evidence can be Adapted, Adopted or requires development
→ Quality assessment-Level of evidence and GRADE
→ Decision and Selection

**Finalisation**

→ External review
→ Recommendation development
→ Final guideline output
→ Plan for Dissemination, Implementation & Review

Figure 1: A summary of the process followed to develop the guideline based on the ADAPTE framework[11]

#### SET UP

A working party consisting of MRS experts, translation/implementation experts and key end-users including higher-degree research students, research radiographers and MRS mentors was established.

#### ADAPTATION

##### Evidence Synthesis

Evidence to inform the guideline was identified through a systematically conducted scoping review (see manuscript for full search strategy[9]). Evidence was summarized, and the ADAPTE framework for guideline adaptation[11] was used in an iterative process to establish where evidence currently exists for each recommendation. The process considers if recommendations are suitable for *Adoption*- when it can be lifted directly from an existing guideline or for *Adaptation*- when the recommendation needs to be adjusted to suit the audience or context. Where no evidence exists, the recommendations require development *DeNovo* ('from scratch')[10].

##### Quality Assessment

The quality of evidence was established in a two-stage process. First, the NHMRC Level of Evidence was established using the traditional and a modified NHMRC hierarchy of evidence framework (Supplement 1). The Level of Evidence describes the suitability of a study design to address a research question (ranging from Level 1 indicating the most robust design to Level 4 indicating the least robust design)[14].In this guideline, studies best answered through a systematic review of randomised controlled trials (e.g. 'Medications') were assessed using a traditional hierarchy of evidence, whilst those examining MRS principles and acquisition parameters, best answered through expert consensus, were assessed using a modified hierarchy (Supplement 1). Second, the modified Grading of recommendations, Assessment, Development and Evaluation (GRADE)[14] determined the degree of certainty in the body of evidence used to inform each of the recommendations (Table 1).

##### Table 1: GRADE matrix

| GRADE | Criteria | Description |
|---|---|---|
| A | → Good evidence (One or more Level 1 study or studies with consistent findings) | Body of evidence can be trusted to guide recommendation |
| B | → Fair evidence (One or more Level 2 or 3 study or studies with consistent findings) | Body of evidence can be trusted to guide recommendation in most situations |
| C | → Conflicting evidence (One or more Level 1 to 3 study or studies with inconsistent findings) OR<br>→ Low level evidence (More than one Level 4 study) | Body of evidence provides some support for recommendation, but care should be taken in its application |
| I | → Insufficient evidence (no studies) OR<br>→ Poor evidence (Level 4-5 studies with inconsistent findings) | Body of evidence is weak, and recommendation must be applied with caution |

#### FINALISATION

##### Expert Panel Agreement

The expert panel consisted of 21 expert MRS researchers from 15 universities in eight countries. A modified-Delphi process[12,13] was used to determine expert agreement on the content and suitability of the recommendation for the Comprehensive Guide (See manuscript[9] for further detail). In brief, recommendations were classified as having 'expert panel endorsement' and accepted into the final guideline if at least 80% of the expert panel had agreed to the recommendation. In cases where recommendations did not reach the 80% threshold in Round 1 or new evidence had become available, recommendations were revised and sent out for re-assessment in Round 2. Recommendations that did not reach the 80%-threshold in Round 2 were not given an 'expert panel endorsement' label.

#### Final Guideline Outputs

The three outputs from this work include this full guideline, a peer-reviewed publication and a one-page infographic summary.

**A Comprehensive Guide to MEGA-PRESS for GABA measurement (This extended guideline)**

This guideline briefly provides an overview of the background and the development process of the guideline, and then provides a detailed document which gives context for each recommendation, an evidence synthesis, and considerations from the expert panel. The full-length guideline is recommended for consultation when upskilling in the field of MEGA-PRESS, particularly during the study protocol design phase.

**The peer-reviewed publication**

The peer-reviewed publication[9] is an accompanying manuscript that first outlines the rigor of the methodological process of recommendation development and then provides a summary of the recommendations. The manuscript provides the GRADE of evidence, percentage of expert panel agreement and a shortened summary of the evidence synthesis and expert panel feedback that informed the recommendation. The manuscript can be used instead of the full-length guideline when a brief overview of parameters that determine data quality is sufficient.

**One-page infographic summary**

The infographic provides a quick visual reference guide, summarises the key messages of the Comprehensive Guide and provides a memory aid to users who have previously read the full guideline. Its purpose is to improve the translation of the guideline into standard practice.

#### Table 2: Summary of Recommendations

| | | | Evidence GRADE | Experts: R1 (%) Agreement | Experts: R2 (%) Agreement |
|---|---|---|---|---|---|
| Acquisition | SNR | -Number of Transients<br>-Voxel Size | A | 76.2<br>81 | 90 |
| | TR | | A | 95.2 | - |
| | TE | | A | 81 | - |
| | Water reference | | A | 85.7 | 55 |
| | Slice selection for water reference | | A | 100 | - |
| | Gradient | | I | 76.2 | 75 |
| | Editing pulse | | A | 76.2 | 90 |
| Practicalities | Voxel position | | A | 85.7 | - |
| | Shimming | | A | 71.5 | 80 |
| | Order of scans | | A | 85.7 | - |
| Confounders | Scanner site | | B | 95.2 | - |
| | Macromolecules | | A | 90.5 | 100 |
| | Region | | C | 81 | - |
| | Tissue composition | | A | 85.7 | 90 |
| | Age | | A | 95.2 | - |
| | Sex | | C | 85.7 | - |
| | Medications | | B | 95.2 | - |
| | Other | -Caffeine<br>-Nicotine<br>-Menstrual phase | I | 71.4<br>76.2<br>71.4 | 85 |
| Data Acquisition | Quality assessment | | A | 90.5 | - |
| | Export | | I | 90.5 | - |
| Quality and Reporting | Quality metrics | | A | 90.5 | - |
| | Reporting | | A | 95.2 | |
| Post-Processing | Frequency and phase correction | | A | 95.2 | |

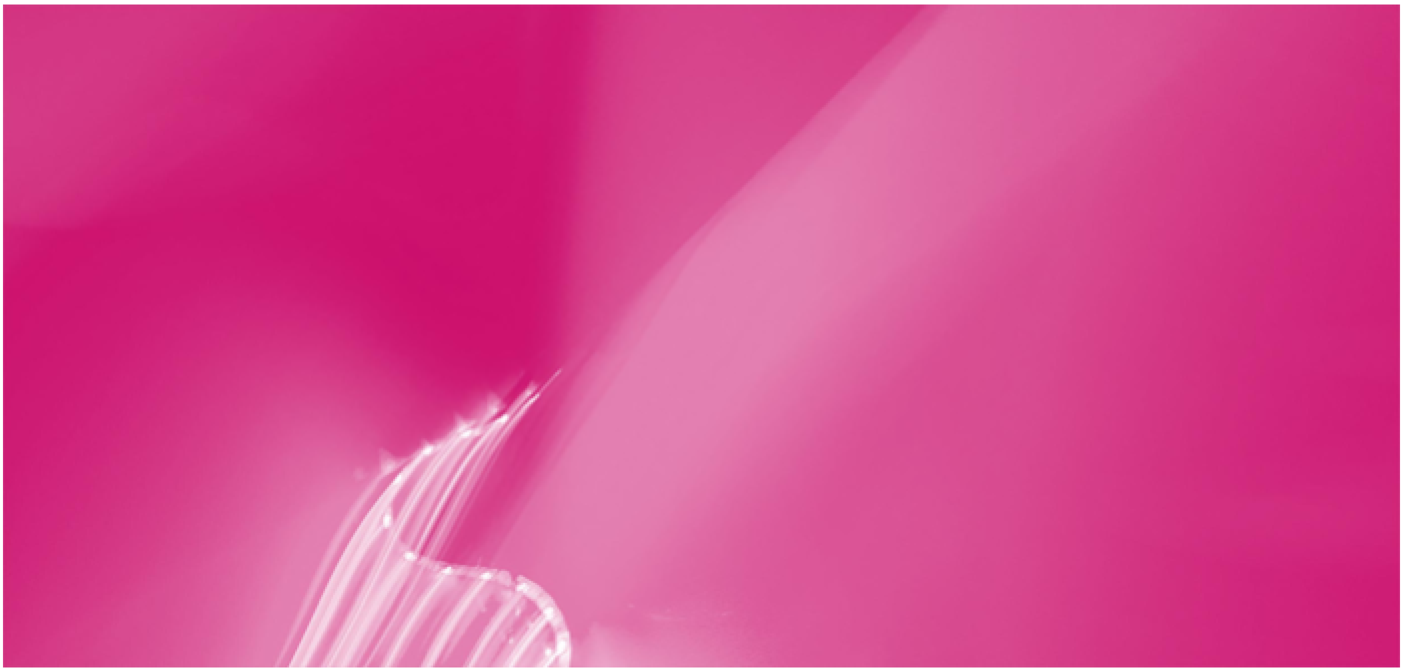

### 1. Parameters


1.1 Signal-to-noise ratio

 1.1.1 Number of transients

 1.1.2 Voxel Volume

1.2 Repetition Time (TR)

1.3 Echo Time (TE)

1.4 Water Reference

1.5 Slice-selection frequency for water reference

1.6 Slice-selective gradients

1.7 Editing pulse specifications

### 1.1. Signal-to-Noise Ratio Considerations (Number of transients and Voxel volume)

| Recommendation | | |
|---|---|---|
| Recommendation | **ADAPT: Start with at least 192 transients (i.e. 96 Edit-ON + 96 Edit-OFF) and a voxel volume of 27ml (e.g. 3 x 3 x $3cm^3$) to quantify GABA when scanning a favourable brain region.** Consider increasing the total number of transients when scanning smaller or more challenging brain regions (See Region) | GRADE A<br>Agreement |

#### 1.1.1. Number of transients

##### Background

MRS measurements suffer from low signal-to-noise ratio (SNR) and require the acquisition of multiple repetitions of the experiment, which are averaged at the end of the scan. The terms transients, averages, excitations or acquisitions have all been used in the literature. Notably, the SNR of the averaged spectrum increases with the square root of the number of transients[15], but linearly with the voxel volume. In order to achieve sufficient SNR, the number of transients and the voxel volume must therefore be considered as a whole. Our recommendation serves to provide a starting point from which further optimisation can be performed, but does not take into account specific adjustments for particular brain regions, conditions or populations.

##### Evidence summary

Eight studies provided recommendations for the number of transients required to acquire data for a MEGA-PRESS experiment (one consensus document; Level 1[6], two large multi-site trials; Level 2[16,17] and five methodological publications; Level 3[8,18-21]). The number of transients recommended for MEGA-PRESS acquisitions ranged from the lowest recommending 126 [20] to the highest recommending 320 [16,17], equating to 4-13 minutes of scan time (at typical repetition time (TR) = 2s). The consensus document suggested 10 minutes scan-time will typically suffice (Level 1). Three of the five methodological publications (Level 3) directly investigated the number of averages required for a MEGA-PRESS dataset and found little improvement in variation when increasing the number of transients from 128 to 296[20], or 200 to 300[18], respectively, with only modest gains demonstrated beyond 218 transients[19]. One methodological publication (Level 3) noted a decrease in stability in the ACC beyond 262 transients[3]. Reasons for reduced stability over longer scans could either be due to increased frequency drift[8,21] or motion artefact[18]. Although a single Level 3 study has demonstrated 126 transients are sufficient, the overall recommendation is to use at least 196 transients (96 Edit-ON and 96 Edit-OFF) to ensure adequate SNR.

A consensus document (Level 1)[22] and a quality assessment tool within a systematic review (Level 2)[2] highlight the importance of *reporting* the number of transients used in the study. Reporting should specify whether the number of acquisitions are separate (as Edit-ON and Edit-OFF) or total number of transients.

#### Considerations

The expert panel commented that the number of transients and voxel volume must be considered together (n=11/21, 52.4%). Choice of brain region and study population will also impact the number of transients required (6/21, 28.6%). Firstly, less favourable brain regions such as the thalamus or dorsolateral prefrontal cortex (DLPFC) may require a greater number of transients to maintain adequate SNR, or alternatively, a larger sample size[18,23], whereas regions such as the occipital and parietal lobe are more favourable to MRS. Secondly, certain study populations, such as paediatric or clinical cohorts, may be more likely to move in longer acquisitions compared to healthy control participants. Therefore, a balance between gaining sufficient SNR and length of scan time needs to be achieved. In addition, it is recommended to choose multiples of 16 to allow for full phase cycles to be included.

Furthermore, it should be noted that the reference to number of transients (total vs. number of ON/OFF) has not been standardized across implementations of MEGA-PRESS and therefore must be checked at time of setup.

#### 1.1.2. Voxel volume

| | |
|---|---|
| **Background** | Voxel volume refers to the volume of the area the spectroscopic signal originates from, i.e. the product of the voxel dimensions. There is variation in how voxel volume is reported in the literature, and may be reported as $mm^3$, $cm^3$ or ml, although consensus documents recommend all three dimensions are reported in the format 30 × 30 × 30 $mm^3$ [22]. |
| **Evidence Summary** | Seven studies provided recommendations for voxel volume (one consensus document; Level 1[6], one seminal text; Level 1[24], two large multi-site trials; Level 2[16,17] and three methodological publications; Level 3[25-27]). All seven studies recommended the use of a ~27ml voxel (e.g. 3 × 3 × 3 $cm^3$, although the voxel does not have to have equal dimensions) for MEGA-PRESS acquisitions (Level 1 to 4). The rationale for this large voxel volume is to compensate for the low SNR of MRS methods, particularly for low-concentration compounds like GABA. One consensus document[6] supports a reduction in voxel volume if the number of averages is increased to adequately compensate for the loss in SNR. It should also be noted that studies using smaller voxel volumes may result in lower GABA estimates (Level 3)[27]. |
| **Considerations** | The relationship between the number of transients and SNR provides 'diminishing returns'. SNR increases only with the square root of the number of transients, while the relationship between voxel volume and the number of transients is linear (Level 1)[24]. For example, an 8-ml volume (e.g. a 2 × 2 × 2 $cm^3$ voxel) only has approximately 30% of the SNR of a 27-ml volume when the number of transients is the same[15]. In this scenario (using an 8-ml vs. 27-ml voxel), the scan time would need to be increased nine-fold to obtain comparable SNR, which is not feasible in most studies. A larger voxel volume will increase SNR; however, it also reduces regional specificity, and further increases partial volume effects (See section 3.4 Tissue composition). To improve regional specificity, the dimensions can be adjusted to make the voxel more rectangular-cuboid-shaped. Quantification is not discussed in detail here, however, using a larger voxel volume increases the importance of including partial volume correction during data analysis. The expert panel commented that high quality data could be obtained with a slightly smaller voxel volume, e.g. 25 ml, (n=6, 28.6%) but it was agreed that 27 ml is an appropriate volume for the beginner to start with when using 192 transients (96 Edit-ON + 96 Edit-OFF). |

#### 1.2. Repetition Time

| Recommendation | | |
|---|---|---|
| Recommendation | **Use a TR of around 2000 ms at 3T.** | GRADE A<br>Agreement |

##### Background

Repetition time (TR) refers to the amount of time from the application of an excitation pulse to the application of the next pulse. TR determines the degree of recovery of the longitudinal magnetization between each repetition of the experiment. This section reports the most commonly used TR across 3T MEGA-PRESS in milliseconds (ms)

##### Evidence Summary

Five studies provided recommendations on TR (one consensus document; Level 1[28], two large multi-site trials; Level 2[16,17], and two methodological publications; Level 3[29,30]). All five papers recommended a TR of 2000 ms for edited MRS of GABA. One methodological publication (Level 3)[29] investigated the $T_1$ of GABA and determined it as 1310 ms. This $T_1$ is the same order of magnitude as other commonly measured metabolites, and therefore does not require the TR to be adjusted beyond 2000 ms for GABA acquisitions.

##### Considerations

This recommendation reached 95.2 % consensus in the first round. However, 3/21 14.3 % of the expert panel commented that TR was unlikely to have a large effect on SNR. Therefore, an appropriate TR could be considered as anywhere between 1500 and 3000 ms. The effect of changing the TR has not been specifically investigated in the literature, however, 2000 ms is most commonly used for single-voxel MRS.

#### 1.3. Echo Time (TE)

| Recommendation | | |
|---|---|---|
| Recommendation | **ADOPT: TE should be 68 ms (GABA+); 80 ms (macromolecule-suppressed GABA).** | GRADE A<br>Agreement |

##### Background

Echo time (TE) is defined as the time between the application of the excitation pulse and the time where optimal refocusing of the signal occurs[13]. For in-vivo proton MRS, TE is usually between 30 and 200 ms. Longer TE results in decreased SNR due to $T_2$ relaxation losses. However, multiplet signals from coupled resonances change with increasing TE, and facilitate the detection of certain compounds with higher sensitivity at longer echo times using experiments such as MEGA-PRESS.

##### Evidence Summary

Ten studies provided recommendations on TE (three consensus documents; Level 1[6,28,31], a systematic review; Level 2[32] two large multi-site trials; Level 2[16,17], three methodological publications; Level 3[30,33,34] and a seminal paper; Level 1[7]). A TE of 68 ms for measuring GABA was first adopted in 1998 when the first seminal MEGA-PRESS study was published. The rationale for using 68 ms is to allow for complete evolution of the GABA multiplet in the edit-OFF acquisition, thus allowing for maximum editing efficiency [7]. The ten studies (Level 1 to 4) [6,7,16,17,28,30-34] have discussed the length of TE. As a result, the consensus remains to keep the TE as close to 68 ms as possible when estimating GABA+. When using a method to suppress macromolecule (MM) contamination, four studies (Level 1 to 4) [17,31-33] agree that the TE should be lengthened to 80 ms to allow for longer, more selective editing pulses, without substantial signal loss due to $T_2$ relaxation (Level 1-4)[7,16,30,31].

##### Considerations

The longer TE used to measure MM-suppressed GABA may not be required for some Siemens scanners as they can apply MM-suppression at 68 ms due to their higher maximum $B_1$ (Level 3)[17].

#### 1.4. Water reference scan required for eddy-current correction and water-scaled quantification:

| Recommendation | ADAPT: Acquire two water reference scans for each volume of interest: One using the same parameters as MEGA-PRESS but deactivated water suppression for eddy-current correction and one short-TE (~30 ms) for quantification. | GRADE A<br>Agreement |
| --- | --- | --- |

##### Background

A water reference scan is an additional acquisition from the same volume, but without water suppression. A water reference scan using the same timing as the water-suppressed MEGA-PRESS experiment is required to perform eddy-current correction. To clarify, the water reference scan should have identical localization, TR, TE and water suppression gradients, but water suppression radiofrequency pulses deactivated. While this acquisition can also be used to perform metabolite quantification relative to tissue water, it will be heavily $T_2$-weighted due to the longer TE. For quantification purposes, an additional water reference scan with the shortest possible TE is therefore suggested. Due to the high concentration of water in the brain, water reference scans require only a few transients, so the time penalty of acquiring two separate water reference scans is negligible[24].

##### Evidence Summary

Seven studies provided recommendations for water reference scans without water suppression for eddy-current correction and water-scaled quantification (five consensus documents; Level 1[6,28,35-37], one seminal text; Level 1[24] and two methodological publications; Level 3[38,39]). One study recommends acquiring a separate short-TE scan to account for the difference in $T_2$ weighting (one consensus document[37]; Level 1). There is consensus across the studies recommending that the water reference scan is acquired from the same volume of interest, using the same gradients in order to facilitate eddy-current correction, water-scaled quantification, and receiver-coil combination.

##### Considerations

Some sequences automatically acquire a water reference during the MEGA-PRESS acquisition, whereas others require a separate scan. While not explicitly stated in the literature, it is necessary to ensure the water suppression gradients are active, while the water suppression pulses are deactivated. The strong signal requires only a few transients for sufficient SNR; the expert panel recommends that typically between 4-8 transients will suffice (4/21, 19%). Round 2 saw a separate short-TE scan for quantification added to the recommendation following expert panel feedback, and a recent consensus document[37]. This significantly reduced expert panel agreement from 85.7 to 55%, suggesting this has yet to be widely accepted in the field.

#### 1.5. Slice-selection center frequency of water reference scan

| Recommendation | ADOPT: Set the water reference to be acquired from the same volume as the GABA signal | GRADE B<br>Agreement |
| --- | --- | --- |

##### Background

The slice-selection frequency is the carrier frequency of the slice-selective RF pulses for the water reference scan. Due to the chemical shift displacement effect, the slice-selection frequency needs to be adjusted appropriately to ensure that the metabolite signals and the water signals originate from the same volume.

##### Evidence Summary

Three studies provided recommendations on the slice-selection centre frequency for the localization of the water reference (two large multi-site trials; Level 2[16,17] and a methodological publication; Level 3[30]). All three studies agree that it should be set to 0 ppm offset, i.e. localizing the 4.7-ppm water signal[16,17,30]. The water-suppressed MEGA-PRESS data is commonly collected with –1.7 ppm offset relative to water, i.e. localizing the 3-ppm GABA signal. This ensures that the water reference scan is co-localized with the same volume as the GABA signal from the MEGA-PRESS scan.

##### Considerations

Implementing the slice-selection centre frequency (also referred to as the delta frequency) may vary between different vendors. Some need to be set explicitly, some can be selected via a drop-down menu and others are fully automated.

#### 1.6. Order of slice-selective gradients

| Recommendation | ADAPT: When artefacts appear in pilot data, consider changing the order of the slice-selective gradients for each volume of interest. | GRADE I<br>Agreement ✕ |
| --- | --- | --- |

##### Background

For PRESS localization, the MRS signal is generated from a cuboid volume by applying three consecutive slice-selective radiofrequency pulses and slice-selective gradients in three orthogonal planes, anterior to posterior, head to foot, and left to right[24]. MRS experiments are sensitive to the appearance of spectral artefacts. These artefacts are often caused by incomplete dephasing of unwanted signal from outside the volume of interest, resulting in out-of-voxel echos (Figure 2). Artefacts are common in regions subjected to abrupt changes of magnetic susceptibility, for example the sinuses or the mouth[24,40]. The order in which the three slice-selective gradients are applied can usually be modified to minimize the appearance of these artefacts.

##### Evidence Summary

One study provided recommendations on the order of slice-selective gradient application (methodological publication; Level 3[40]). The study recommended that the axial gradient should be applied last for frontal voxels, in order to minimize out-of-voxel water ("ghost") artefacts in the data[40]. No other studies have discussed the order of slice-selective gradients, however this was discussed at an international expert workshop in 2018[41]. The consensus from this workshop was to conduct 2-minute pilot scans with varied gradient order prior to commencing data collection. Beginners should then observe the impact of gradient order on spectra with respect to reducing artefacts (e.g. lipid contamination or out-of-voxel echoes) and improving spectral quality. This recommendation is supported by experiments conducted yet unpublished (Level 5)[15], where substantially less lipid in a motor region was demonstrated when gradient order was optimized.

#### Considerations

Not all vendors allow for the easy adjustment of gradient order. In instances where MEGA-PRESS has not been run on a scanner before, a series of pilot acquisitions (as detailed above) should be completed to select the optimal gradient order. The expert panel suggest if artefacts exist and gradient order cannot be adjusted, the VOI can be rotated slightly (2/21, 9.5%). Note that rotations beyond 45 degrees may automatically flip the direction of the gradients, which can make the directions of the chemical shift displacement difficult to predict, particularly when rotating the voxel in more than one plane. Experts also suggested using slice-selective pulses with large bandwidth (e.g. adiabatic localization) to reduce effects of static magnetic field ($B_0$) inhomogeneity, although these may not be available in every implementation (2/21, 9.5%). Some possible artefacts are demonstrated in Figure 2, but for further information it is recommended to refer to Kreis *et al.* (2004)[42] for visual examples of other common spectral artefacts (2/21, 9.5%).

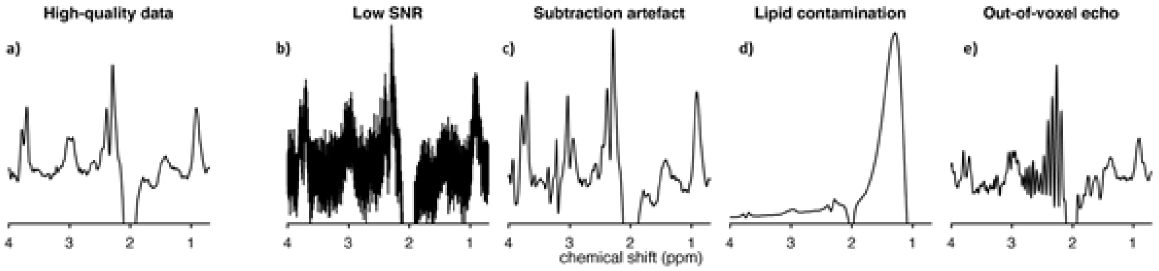


**Figure 2: Common MEGA-PRESS data quality issues.** a) High-quality data with sufficient SNR, narrow linewidths, a well-defined edited signal at 3 ppm, and no substantial artefacts; b) very high noise levels due to low number of transients or small voxel volume; c) severe subtraction artefacts due to scanner frequency drift; d) lipid contamination due to participant motion or voxel positioning too close to the skull; e) out-of-voxel echo ("ghost signal").

#### 1.7. Editing pulse specifications

##### Recommendation

**ADOPT: Editing pulses can be applied as follows:**

| | GABA+ | Macromolecule -suppressed |
|---|---|---|
| **Frequency (ppm)**<br>Edit-ON<br>Edit-OFF | <br>1.9 ppm<br>7.46 ppm | 1.9 ppm<br>1.5 ppm |
| Bandwidth | 60 Hz | Usually 80Hz (60 Hz on some implementations) |
| Spacing | 0.5 TE apart (this parameter is usually not accessible to the user) | |

GRADE

A

Agreement

##### Background

Editing pulse frequency is defined as the frequency at which the frequency-selective editing pulses are applied. The frequencies of the editing pulses are different dependent on whether GABA+ or MM-suppressed GABA is being acquired. In GABA+ data, the edited signal at 3 ppm is contaminated by co-edited MM signal (estimated to account for about 50% of the edited signal area[43,44]). To reduce the MM contamination, a second editing pulse can be applied at 1.5 ppm. However, the increase in specificity comes at the expense of a much greater sensitivity to experimental instability, particularly thermal drift of the magnetic field. Editing pulse bandwidth refers to the full-width half-maximum (FWHM) bandwidth of these pulses, which is a measure for their selectivity, and inversely related to their duration. Editing pulse spacing refers to the time between the two editing pulses. Not all these settings can be adjusted by the user, depending on the sequence.

##### Evidence Summary

Nine studies provided recommendations on the frequency, bandwidth, and spacing of the editing pulses. Five discussed the frequency of the editing pulse for GABA+ (one consensus document; Level 1[6], two large multi-site trials; Level 2[16,17] and four methodological publications; Level 3[7,30,45,46]) and four discussed the position of the editing pulse for MM-suppressed GABA (Level 3) [17,34,46,47]. The consensus for GABA+ was that editing pulses should be placed at 1.9 ppm and 7.46 ppm (Level 3 to 4) [7,16,30,45,46]. For MM-suppressed GABA, the editing pulses need to be positioned symmetrically around the MM resonance at 1.7 ppm (i.e. 1.9 ppm / 1.5 ppm)[17,34,47]. Five of the nine studies specifically recommend that the editing pulses are spaced TE/2 apart (Level 1 to 3)[6,16,17,30,46] . However, certain implementations on Siemens and Canon platforms do not comply with the TE/2 requirement, therefore reducing editing efficiency if deviating from TE = 68 ms (Level 3)[30,46]. The editing pulse bandwidth should be kept as narrow as possible (FWHM = 60–80 Hz). The minimum achievable bandwidth may depend on vendor, sequence implementation, and available hardware (Level 1 and 3). Given the duration of the editing pulse is inversely proportional to the bandwidth, the duration should therefore be as long as the TE permits- usually around 15 ms for GABA+ and 20 ms for MM-suppressed GABA (Level 2 to 3)[16,17,30].

#### Considerations

The definition and specification of the editing pulse bandwidth differs between sequence implementations. Some implementations require the editing pulse duration as the input, while others require the FWHM of the bandwidth. Notably, the FWHM entered on the exam card may differ from the actual FWHM of the editing pulse. For example, Siemen's implementations that apply a smoothing filter to the pulse, result in an actual FWHM considerably larger than the nominal one. Therefore, specifications for bandwidth duration were not added to this recommendation. The expert panel had numerous suggestions for variations on these parameters (n=8, 38.1%), which highlight that there is variation on what can be applied at an expert level.

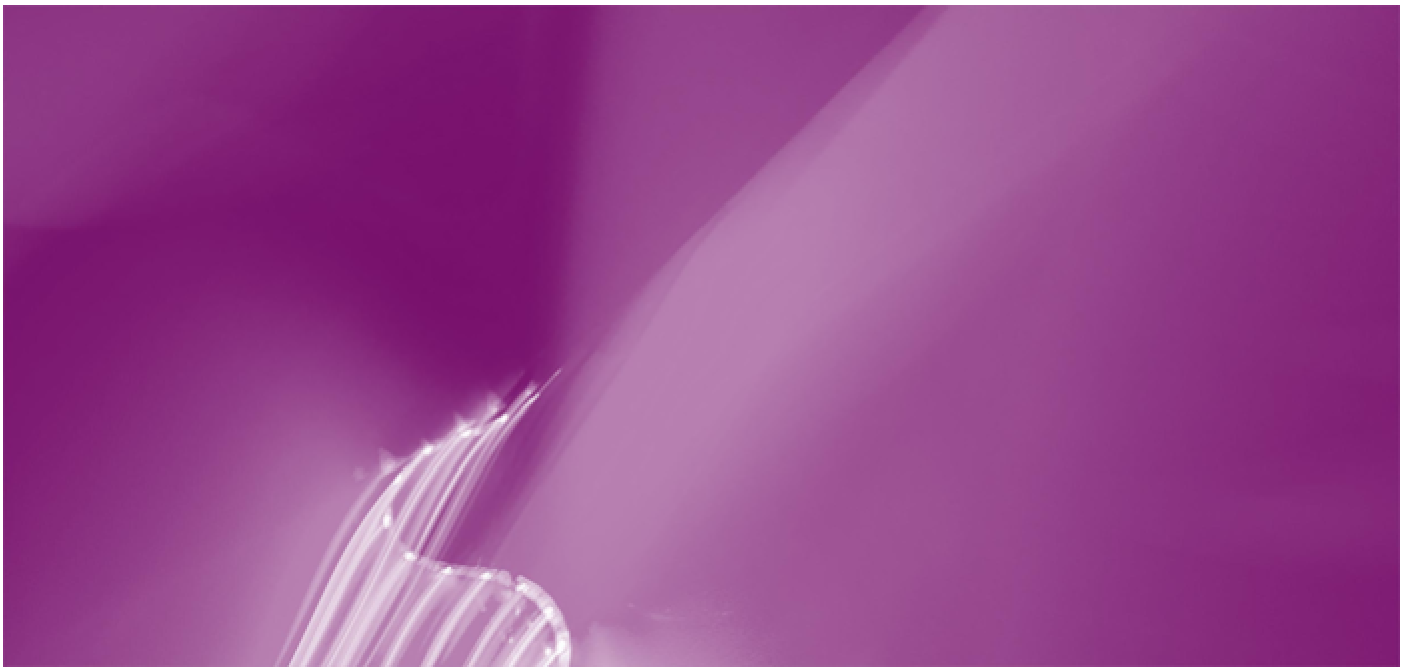

### 2. Practicalities


2.1 Voxel position

2.2 Shimming

2.3 Order of scans and field drift

#### 2.1. Voxel position

| Recommendation | ADAPT: Use automated voxel positioning tools where available. If manually positioning the voxel use a screenshot and clear instructions regarding positioning relative to anatomical landmarks and degree of rotation. | GRADE A<br>Agreement |
|---|---|---|

##### Background

Voxels are generally positioned according to the research interest of the investigator. However, several factors limit the freedom to position voxels. In this section, practical approaches to voxel placement are discussed.

##### Evidence Summary

Five studies provided recommendations on the practicalities of voxel positioning (one consensus document; Level 1[35], three methodological publications; Level 3[27,48,49] and one narrative review; Level 4[42]). One of the 5 studies (Level 4)[42] demonstrates the implications of positioning the voxel. The positioning of voxels has the potential to cause significant variation in data: Two of the five studies (Level 3) examined reproducibility of manual voxel placement and found that the overlap in repositioning the voxel within a scan ranged between 75% and 85%, 75%, corresponding to a 2-3 mm displacement along three axes[48,38]. Therefore, to aid manual voxel placement it is recommended that a screen-shot is used with detailed written instructions including reference to anatomical landmarks. A consensus document (Level 1) and a methodological publication (Level 3) recommend the use of an automated voxel positioning tool . This has been found to improve reliability of voxel repositioning both within and between scans[35,49].

##### Considerations

When manually positioning a voxel, care needs to be taken to not position the voxel too closely to the skull, since dural fat signals may contribute strong lipid signals that significantly distort the spectrum and hamper subsequent quantification (Figure 2). Significant lipid signals may still occur even if the voxel shown on the inline display did not contact the dura, this is due to the chemical shift displacement effect. Some vendors allow the display of a second voxel box to visualize the origin of a shifted metabolite signal (e.g. the lipid resonance), which can help guide voxel placement.

Brain regions such as the occipital lobe or parietal lobe are considered favourable for scanning. It is therefore recommended that when first utilizing MEGA-PRESS sequences, the beginner pilots their methodology in these regions prior to scanning more challenging regions. More challenging regions involve positioning voxels deeper in the brain (i.e. further away from receiver coils), close to ventricles or iron deposits (e.g. subcortical regions). These may suffer from decreased signal-to-noise ratio (SNR) or spectral quality compared to cortical voxels (Figure 2).

Fully automated voxel positioning software is not currently integrated into standard scanner operating software (6/21, 28.6%). The expert panel note that freely available AutoAlign (Siemens), ReadyBrain (GE), SmartExam (Philips) or NeuroLine (Canon) software all can improve alignment reproducibility by referencing the anatomical images on which a volume of interest (VOI) is planned. The accuracy of these tools relies on the quality of the anatomical images and consistency of voxel placement should be carefully reviewed during acquisition (2/21, 9.5%).

#### 2.2. Shimming

| Recommendation | A beginner should use a readily available automated field-map-based shim and minimize the use of manual adjustments. | GRADE A<br>Agreement |
|---|---|---|

##### Background

Shimming is the process of maximizing the homogeneity of the static magnetic field ($B_0$) over the measurement volume of interest. Since high homogeneity results in narrow linewidths and increased SNR, the quality of the shim is considered one of the most important parameters for determining spectral quality[24]. Shim is typically reported in terms of the full-width at half-maximum (FWHM) of the water linewidth and is reported in Hz, although this may vary between vendors. Linewidth values largely depend on brain region and the surrounding interfaces between air/tissue and tissue/bone. These will influence the magnetic field causing field distortions[28]. Most vendors offer automated field map-based shim routines and/or projection-based shim routines and may also offer manual adjustment of the shim currents. Dynamic shim updates are the subject of ongoing research, and not readily available across all systems (refer to Juchem *et al.* 2020[50] for visual representation of the impact of shim quality on the spectra.)

##### Evidence Summary

Eight studies provided recommendations on the practicalities of shimming (three consensus documents; Level 1[28,35,50], one systematic review; Level 2[51] and four methodological publications; Level 3[20,52-54]). The consensus across the eight studies was that FASTMAP, FASTESTMAP (projection-based shim optimization) or second-order pencil beam methods could provide narrower linewidths than the default 3D-field-map-based methods. However, these systems are not openly available and are technically more challenging to operate. It is further recommended that manual shimming should only be implemented to optimize suboptimal automated shims to minimise user intervention (Level 1) [35]. Three consensus documents (Level 1)[28,35,50] and one methodological publication (Level 3) [52] recommend that the attainable values of $B_0$ shim quality are expressed as linewidths of the water peak. The three consensus papers (Level 1)[28,35,50] report that; at 3T a FWHM of 5-7 Hz is considered excellent, 8-10 Hz is considered good and 11-13 Hz acceptable for the brain. However, these values are significantly lower than those reported in the methodological publication (Level 3)[52]. These values are likely to increase in brain regions that are more difficult to shim, such as frontal, temporal or subcortical regions. An example is the frontal region where a shim of <16 Hz is considered acceptable[52].

#### Considerations

Evidence suggests that projection-based shim optimization (e.g. FASTMAP, FASTESTMAP) methods or pencil-beam methods can potentially achieve a better shim than default 3D-field-map-based methods. However, experts highlight these are not readily available and can be more challenging for a beginner to use (3/21, 14.3%). Therefore, the expert panel recommend that readily available automated field-map-based methods are used by the beginner and that manual adjustments are avoided wherever possible (9/21, 43%).

The expert panel commented that different vendors calculate spectral linewidths differently (8/21, 38.1%). Therefore, the inline display values of linewidth may not correspond to the above mentioned quality criteria. It is therefore useful to measure the actual FWHM from processed spectra to determine how it relates to the inline-displayed value for that system.

#### 2.3. Order of scans and field drift

| Recommendation | ADOPT: Where possible MRS should be conducted prior to gradient-heavy acquisitions or in small blocks of 2-5 minutes with frequency adjustments between adjustment blocks. Consider using real-time frequency correction if available. | GRADE A<br>Agreement |
|---|---|---|

##### Background

The order of scans refers to the position of the MRS acquisitions within the study protocol. Several critical effects relating to the order of scans need to be considered during protocol design. This section focuses on issues associated with field drift resulting from imaging sequences widely used in neuroscientific and clinical studies.

##### Evidence Summary

Eight studies provided recommendations with regard to field drift. Five specifically with regard to the order of scanning (three consensus documents; Level 1[31,35,36] and two methodological publications; Level 3[8,34]). Three discussed the use of real-time frequency correction (three consensus documents; Level 1[31,36,55]) and two investigated the impact of field drift on reported GABA level (large multi-site trials; Level 2)[16,17]. Five of the eight studies (Level 1-4)[8,31,34,36,55] highlighted the negative impact gradient-heavy scanning such as diffusion-tensor imaging (DTI) has on frequency drift during subsequent MRS scans. Frequency drift was observed for as long as 30 minutes following a fMRI scan[8,36]. The degree of frequency drift can vary between scanners. The range of frequency drift reported in the two methodological publications following fMRI/DTI (Level 3) were; –2 Hz/min[8] and 4.6 Hz/min respectively on a MM-suppressed acquisition[34]. While all MRS studies are susceptible to drift, MM-suppressed MEGA-PRESS is an order of magnitude more susceptible[34]. The impact of frequency drift is that it reduces editing efficiency, changes signal and increases subtraction artefacts (Figure 2) [8,34]. For conventional MEGA-PRESS, drifts of 10 Hz will result in a moderate signal change of 4-6%, while the MM-suppressed GABA signal may change by approximately 30%[8,34]. Given the potential impact of frequency drift on editing efficiency, it is recommended that MRS be conducted before any fMRI or DTI with application of real-time (prospective) frequency correction if available[8,34,35]. A recent consensus document (Level 1) proposes that data be acquired in small blocks of 2-5 minutes to monitor frequency drift or subject motion with interleaved scanner frequency adjustments between acquisition blocks [36].

An additional recommendation regarding the order of scanning is that the water reference scan is acquired first (Level 1)[26]. This will ensure that the water reference is acquired from the same VOI in case the metabolite acquisition needs to be stopped and/or repeated due to participant motion.

#### Considerations

It is acknowledged that conducting MRS prior to high-gradient imaging is not always possible. The expert panel recommended establishing the drift behaviour of an individual scanner by piloting MRS before and after a functional MRI (fMRI) or diffusion tensor imaging (DTI) sequence in order to gauge the potential effects on subsequent MRS (2/21, 9.5%). Alternatively, experts endorse the new recommendation to acquire MRS data in small blocks with frequency adjustment after each block, whilst monitoring the water residual during the scan acquisition in order to detect drift (3/21, 14.3%).

Although one consensus document (Level 1) [35] recommends acquiring the water reference scan prior to the metabolite spectrum, this might not be possible for all vendors e.g. GE. An alternative might be to consider interleaving the water reference.

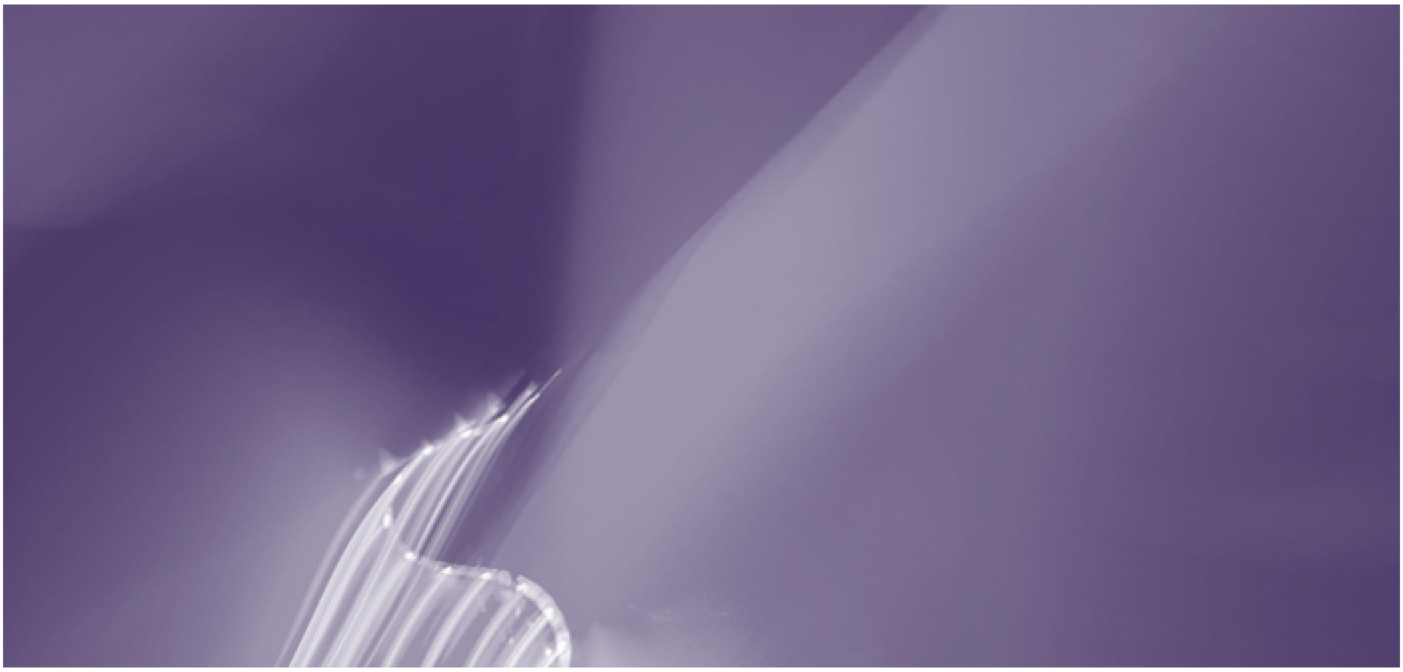

### 3. Confounders

This section covers known and potential confounders to MRS studies of GABA. This list is not exhaustive due to the specific nature of our search strategy and the wide range of known and unknown potential confounders. This section focuses on major confounders including:

3.1 Scanner site and vendor

3.2 Macromolecules

3.3 Region

3.4 Tissue composition

3.5 Age

3.6 Sex

3.7 Medication

3.8 Other potential confounders which include caffeine, nicotine and phase of menstrual cycle.

(Note levels of evidence documented with 'Level $X^{T}$' are assessed using a traditional hierarchy of evidence[14] rather than the modified-hierarchy of evidence (Supplement 1).

#### 3.1. Scanner site and vendor

| Recommendation | In multi-site studies standardized protocols should be used, and the degree of systematic differences between site/scanner should be reported. | GRADE B<br>Agreement |
|---|---|---|

##### Evidence Synthesis

Three studies discussed site and vendor as confounders for GABA estimates (three multi-site trials; Level 2 [16,17,46]). Two of these studies (Level 2)[16,17] analysed a dataset from 272 participants across 24 sites, using vendor-specific MEGA-PRESS implementations.  The studies reported a coefficient of variation across all data sets of around 12% for GABA+/Cr and 17% for water-scaled GABA+. MM-suppressed MEGA-PRESS had larger CVs of 28%-29% for both GABA/Cr and water-scaled GABA [16,17].

Linear-mixed effects analysis of variance showed that only 20% of the overall variance of GABA+/Cr measures was accounted for by site-level differences, while 8% was accounted for by differences between scanner vendors. In contrast, water-scaled GABA+ data variance was mainly accounted for by between-vendor difference (53% of total variance) with just 11% being accounted for by site-level differences. The third study (Level 2)[46] showed that using a 'universal' MEGA-PRESS sequence that was implemented for all major vendors (with identical timing, RF pulses, and gradients) improved the within-subject agreement of GABA+/Cr estimates acquired on different systems compared to the vendor-specific implementations. However, this study only included eight participants.

##### Considerations

There is considerable difference between individual scanners, especially with different vendors. To establish the difference between sites scanning the same phantoms or control participants on all the scanners might help to quantify the between scanner differences. When designing a multi-site study always use a balanced design where the same number of controls and participants are scanned on each of the scanners (2/21, 9.5%).

#### 3.2. Macromolecules

| Recommendation | | GRADE / Agreement |
|---|---|---|
| Recommendation | **ADAPT: A beginner should use conventional MEGA-PRESS reporting GABA+. Macromolecule contamination should be acknowledged as a limitation, and consideration paid to whether macromolecules could be responsible for between-group differences.** | GRADE A; Agreement |

##### Evidence Synthesis

Eleven studies discussed MM-contamination as a confounder of GABA (three consensus documents; Level 1 [6,31,36] and eight methodological publications; Level 3 [33,34,43,44,47,56-58]). A limitation of conventional MEGA-PRESS is the co-editing of MM that underlie the 3-ppm GABA signal. The degree of this contamination has been reported to be within 41%[43] and 60% of the total GABA+ signal area (Level 3)[44]. Three main approaches to account for MM contamination have been proposed, however all three approaches have significant limitations as agreed in two consensus documents[6,31]. The most widely used approach is symmetric MM-suppressed editing. This technique, however, is highly susceptible to frequency drift, thus reducing the reliability of MM-suppressed GABA measurements compared to conventional editing for GABA+2. Only one of seven methodological publications (Level 3)[44] found comparable repeatability of symmetric MM-suppressed editing compared to conventional GABA+ MEGA-PRESS. Recent consensus documents (Level 1) on MM in MRS[31] and edited MRS[36] recommend using MM-suppressed editing where possible. However, they also acknowledge the limitations of this approach and that it might not be practical in a clinical environment.

When interpreting data it should be noted that, while there was a moderate correlation between GABA and GABA+ levels pooled across brain regions in two methodological publications (Level 3)[33,56], there was only a weak correlation in a region-specific analysis[33]. Therefore, care needs to be taken when comparing or pooling results from conventional GABA+ and MM-suppressed GABA studies.

##### Considerations

The expert panel recommended that beginners should use conventional MEGA-PRESS at present despite consensus documents recommending the use of MM- suppressed sequences (19/21, 90.5%). A MM-suppressed sequence is more challenging for beginners to acquire because it is more susceptible to experimental instabilities such as frequency drift. Therefore, the expert panel recommends that in line with a previous consensus document (Level 1)[6] beginners adopt the most widely utilized approach of conventional GABA+ acquisition. They should report this as GABA+, acknowledging MM-contamination of the edited signal as a limitation. The MM baseline could be measured in a group of control participants, if differences in MM might explain between group-differences in the study population or between the time points[31].

#### 3.3. Region

| Recommendation | | |
|---|---|---|
| Recommendation | **ADAPT: Select brain regions relevant to research question, however, acknowledge that brain regions have differing reliability with respect to data acquisition.** | GRADE A<br>Agreement |

##### Evidence Synthesis

Fourteen studies discussed brain region as a confounder of GABA (1 review; Level 1[T 59], 13 methodological publications; Level 4[T 19,27,52,60-69]). Historically, it was hypothesised that brain GABA Levels may be universal across all brain regions, reflecting a “global GABAergic tone”[59]. However, there is growing evidence that this is not the case (Level 4[T]) [27,52,60-62,65,68,69]. Several methodological publications have demonstrated that GABA levels are different between anterior and posterior brain regions (Level 4[T])[52,61,68-70], but less so between hemispheres (Level 4[T])[52,66].

##### Considerations

It is important to consider that certain brain regions may be less suitable for stable and reliable data acquisition than others depending on size, depth, tissue composition of the voxel and whether signals are obtained from cortical or subcortical regions. The occipital lobe, and posterior cingulate gyrus, for example, are associated with high quality spectra whereas regions such as the amygdala are more challenging. (See sections 1.1 SNR and 2.1 voxel position).

#### 3.4. Tissue composition

| Recommendation | | |
|---|---|---|
| | **ADAPT: Water-scaled quantification methods should consider the impact of partial volume effects on GABA estimation.** Segmented structural images should be used along with a tissue correction method to account for grey matter, white matter and cerebrospinal fluid. Grey-matter only correction should be avoided. | GRADE A<br>Agreement |

##### Evidence Synthesis

Nine studies discussed the relative volumes of grey and white matter within the MRS volume, as a confounder for GABA estimates (one consensus document; Level 1 [6], eight methodological publications; Level 3 [66,71-77]). There was agreement across all nine studies that GABA levels are higher in grey matter than white. This is substantiated by data using brain tissue extracted during surgery[78] and chemical shift imaging studies[79] (not included in this review). The studies that used MEGA-PRESS and optimised parameters found that GABA levels are approximately twice as high in grey compared to white matter[74-77]. The three other studies reported a range of ratios from 2:1[72] to 8:1[71,73]. All of these studies investigated a healthy population or simulation, the ratio may be altered in the presence of pathology.

The recommended approach for handling grey and white matter differences within the MRS volume is debated. One methodological publication (Level 3)[76] demonstrated that choice of tissue correction method significantly impacts the water-scaled quantification of GABA+ and therefore needs to be considered with care. All studies agree that correction for grey matter alone is insufficient and leads to over estimation of GABA, especially in voxels containing less than 50% grey matter. All methodological publications recommend a degree of tissue correction which allow for the different voxel composition of grey matter, white matter and CSF when using water-scaled quantification. However, this approach alone does not take into account tissue specific relaxation times. One methodological publication (Level 3)[74] investigated tissue relaxation times and recommends the use of pulse sequence parameters that minimize the effect of signal relaxation, owing to not knowing the composition of the voxel *a priori*. The consensus document (Level 1)[6] recommends that the most appropriate approach is to use grey matter to white matter ratios as a covariate in any statistical analysis rather than to attempt to correct measures based on the reported differences in concentration between tissue types.

##### Considerations

There are a number of tissue correction algorithms available, however there are limitations to each approach. Beginners should be aware of the limitations of their chosen approach (e.g. unaccounted difference in relaxation times). Using tissue composition as a covariate helps to clarify that between-group differences in GABA are driven by differences in GABA levels rather than by differences in tissue composition (3/21, 14.3%). However, including tissue composition as an additional covariate may reduce power in study with a small sample size (1/21, 4.8%).

#### 3.5. Age

| Recommendation | ADOPT: Age is likely to affect GABA levels, therefore age should be accounted for in study design or statistical analysis. | GRADE A<br>Agreement |
|---|---|---|

##### Evidence Synthesis

Seven studies investigated age as a confounder for GABA (one systematic literature review and meta-analysis[80]; Level 1[T] and six methodological publications; Level 3[T] [68,69,81-84]). All seven papers report that GABA+ decreases with age, however, one found no relationship between MM-suppressed GABA and age. One methodological publication (Level 3[T])[68] proposed that the observed decrease in GABA levels is a result of grey matter atrophy, and further supports the recommendation to correct for tissue composition. One of the six methodological publications (Level 3[T])[69] reported a 5% decrease in GABA/Cr and 4% decrease for GABA/NAA per decade, however, this was not calculated for GABA to water ratios or investigated in any other study. The meta-analysis[80] (Level 1) that extracted single-subject data found an increase in GABA in early development, plateauing in adolescence and early adulthood, followed by a steady decline with age.

##### Considerations

Age should be considered as a covariate due to a substantial age trajectory, however, including age as an additional covariate will reduce the power of a study with a small sample size. Use of an age-matched design, i.e. matching the age of participants across all groups may avoid the need to include age as an additional covariate.

#### 3.6. Sex

| Recommendation | ADOPT: Sex is likely to affect GABA levels, therefore sex should be accounted for in study design or statistical analysis. | GRADE C<br>Agreement |
|---|---|---|

##### Evidence Synthesis

Four studies investigated sex as a confounder for GABA (four methodological publications; three Level 3[T][69,81,85] and one Level 4[T][86]. The sample size ranged from 14[86] to 100[69] participants. The study with the largest number of participants found no difference in GABA+ levels between males and females in the anterior cingulate cortex (ACC). These results were reproduced by two other studies investigating the ACC[81,85]. Conversely, two studies of the parietal and dorsolateral prefrontal cortex (DLPFC) found statistically significantly higher levels of GABA in males compared to females[85,86]. Taken together, these results suggest that sex differences in GABA may be region-specific.

##### Considerations

When designing a study, consider recruiting equal numbers of female and male participants unless the study has an important sex component, or the condition being studied is more prevalent in a particular sex. A study design with sex-matching between groups can also be used to account for sex differences.

#### 3.7. Medication

| Recommendation | ADAPT: Medications may affect GABA levels, as minimum best practice all medications should be recorded. | GRADE B<br>Agreement |
|---|---|---|

##### Evidence Synthesis

Eight studies discussed medications that may confound GABA (one systematic review; Level 1[T] [59]one RCT; Level 2[T] [87], six methodological publications; two Level 3[T] [88,89] and four Level 4 [T] [90-93]). In the seven clinical studies returned by our search, five drugs were investigated: vigabatrin, citalopram, zolpidem, gabapentin, tiagabine. Level 4 evidence suggests vigabatrin, citalopram, zolpidem, gabapentin may confound GABA measurements, while the data were inconclusive regarding tiagabine. The systematic review (Level 1[T]) [59] further concluded GABA levels might increase following administration of levetiracetam or topiramate but not valproate, carbamazepine and phenytoin, and lamotrigine. Taken together, brain GABA levels may be influenced by a variety of medications regardless of whether their primary mechanism of action is on the concentration of GABA itself for GABA receptor agonists or antagonists and therefore medication should be recorded and considered as a potential confounder of GABA (Level 1 [T])[59].

##### Considerations

The aim of this section was to highlight that medications may confound measures of GABA. Given the broad aim of our scoping review, our evidence synthesis is not a full systematic review of this question. As a result, studies investigating the confounding effects of specific medications on GABA levels may have been missed. It is recommended that a medical specialist is consulted to discuss the mechanism of action of any pertinent drugs in the planned studies population. It is important that a considered decision is made with regard to handling patients who are medicated. It is likely that exclusion of these participants will considerably bias the study population.

#### 3.8. Other potential confounders: Nicotine, Caffeine, Phase of menstrual cycle

| Recommendation | ADAPT: Potential confounders such as caffeine and nicotine intake and phase of menstrual cycle may affect GABA levels, as minimum best practice potential confounders should be recorded. | GRADE I<br>Agreement |
|---|---|---|

##### 3.8.1. Nicotine

**Evidence Synthesis**

Two studies investigated nicotine as a confounder for GABA levels (two methodological publications; Level 3[T 94,95]). One study found no difference in GABA Levels between 48 heavy smokers (n=48) and healthy controls[94]. Another study found no difference in GABA Levels in 36 smokers between baseline measures and following 48 hours abstinence[95].

##### 3.8.2. Caffeine

**Evidence Synthesis**

One study discussed caffeine as a confounder of GABA Levels (methodological publication; Level 4[T 96]). A study of 15 healthy participants found no significant difference in GABA levels before and after acute administration of 200 mg of caffeine.

##### 3.8.3. Menstrual Cycle

**Evidence Synthesis**

Four studies discussed the menstrual cycle as a confounder of GABA (four methodological publications; two Level 3[T 97,98] two Level 4[T 60,95]). One[95] of the four studies investigated phase of menstrual cycle as a secondary aim looking at a subgroup of six participants and therefore did not provide sufficient data to determine the effect of phase of menstrual cycle. The three remaining studies had sample sizes ranging from seven[60] to 75[98] participants. The study with the largest sample size found higher GABA levels during ovulation compared to the rest of the cycle in women with a natural cycle[98]. There was no difference between the follicular and luteal phases. In contrast, the two remaining studies found higher levels of GABA in the follicular phase compared to the luteal phase, but did not investigate GABA during ovulation[60,97]. Furthermore, one paper investigated women taking the hormonal contraceptive pill and found no difference between the active or inactive pill[98]. Current evidence suggests menstrual cycle may affect GABA levels, although there are some methodological limitations to the included studies.

##### Considerations

The effect of caffeine, nicotine and phase of menstrual cycle on GABA cannot be fully established from current evidence. Therefore, it is suggested that the impact of these potential confounders are considered in the design of the study, especially when conducting longitudinal or repeat measure studies.

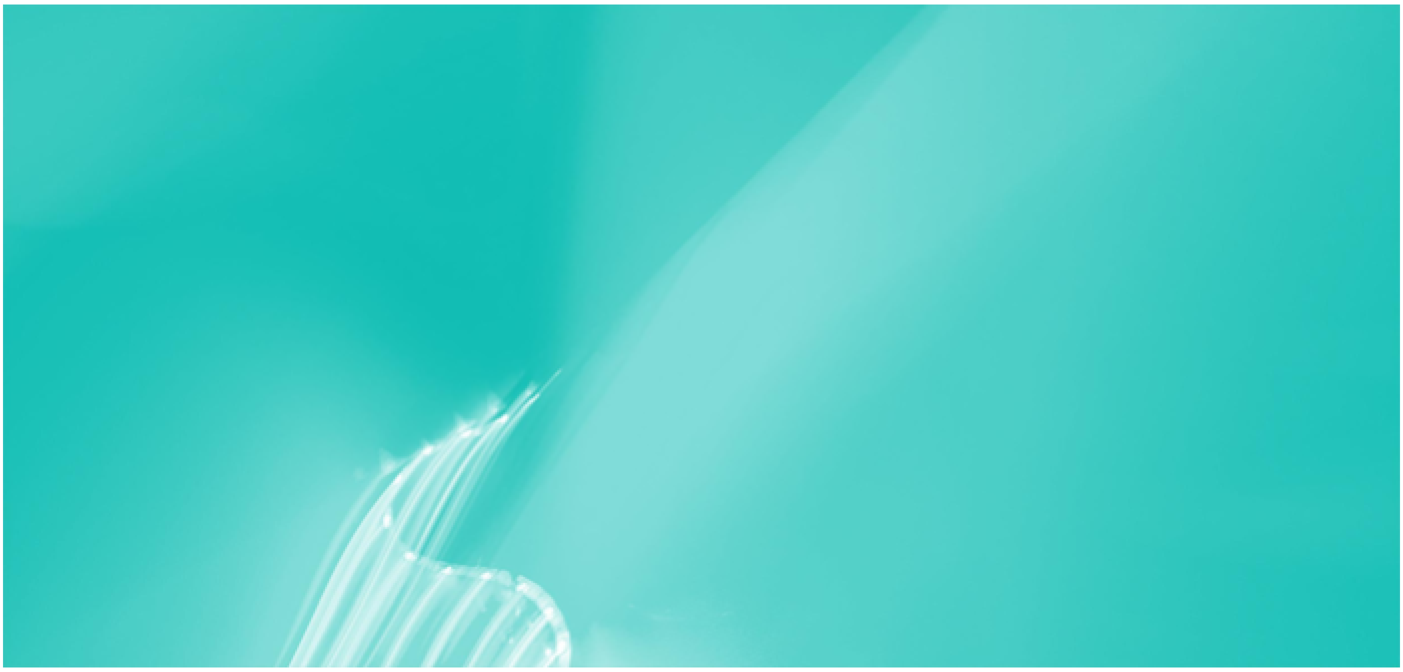

### 4. Data Acquisition

4.1 Quality assessment during the scan

4.2 Data export

#### 4.1. Quality assessment during the scan

| Recommendation | | |
|---|---|---|
| Recommendation | **ADOPT: It is recommended to monitor the quality of the acquisition using the inline data display at time of scanning.** Scans should be cancelled, and voxel position adjusted if evidence of weak water suppression, strong lipid contamination or other artefacts. | GRADE A<br>Agreement |

##### Preface

Most modern MRI scanners offer inline displays showing the last acquired spectral transient. This display can be used to determine spectral quality during the acquisition (water suppression, potential lipid contaminations and other artefacts), and make time-saving decisions whether a scan should be cancelled (and potentially repeated), or the voxel should be repositioned.

##### Evidence summary

Two studies provided recommendations to monitor quality of data acquisition during the scan (two consensus documents; Level 1[35,36]). Both consensus recommended that the MR operator should evaluate and monitor water suppression efficiency, spectral linewidth and signal-to-noise ratio at the beginning and during the MRS acquisition. A change in linewidth, frequency or spectral pattern, or worsening water suppression, suggests the participant has moved. It is recommended that the participant is visually checked, and the acquisition repeated if necessary (potentially including the localizer/scout image to account for the new participant position).

##### Considerations

Experts highlight that not all vendors provide the option to monitor the scan using an inline display at time of scanning e.g. GE. One expert noted that running an inline display can affect the TR on certain systems (prolonging TR up to 200 ms), this has important implications for relaxation correction or functional MRS experiments.

#### 4.2. Data export

| Recommendation | DEVELOP: Export data in a format that saves individual transients to allow adequate post-processing. | GRADE I<br>Agreement |
|---|---|---|

##### Preface

Spectroscopic data is often saved in vendor-specific file formats with varying degrees of processing. To ensure that all necessary post-processing steps can be performed, export MEGA-PRESS data in a file format that stores all individual transients separately.

##### Evidence summary

There is currently no discussion concerning which files to export specifically for MEGA-PRESS acquisitions, however, one study generally discusses the file format to export for MRS studies which could also be applied to MEGA-PRESS studies (consensus document; Level 1[37]). This consensus document (Level 1)[37] recommends that data be saved as single transients to allow for post-acquisition frequency-and-phase aligment[37]. Based on MEGA-PRESS-applicable recommendations extracted from the consensus document, we have developed the following recommendation:

| Scanner | Format | Description | Comment |
|---|---|---|---|
| Philips | SDAT/SPAR | Two files for each acquisition, SDAT contains acquired signal data, SPAR contains header info. | Use SDAT/SPAR only when individual transients are exported. If this is not the case, also export DATA/LIST (which does not contain voxel location information, so both formats are then required). |
| | DATA/LIST | As above, DATA, LIST respectively. | |
| GE | GE-P (.7) | Default combines RF coil channels and groups in a phase encoded step. Fully customizable to preserve or combine any/all dimensions. | |

| Scanner | Format | Description | Comment |
|---|---|---|---|
| Siemens | TWIX (.dat), | All dimensions (RF channels, transients) preserved without modification. | Older sequence implementations may not allow the export of single-average RDA files. |
| | single-average RDA | All dimensions (except time/spectral dimensions) are pre-combined.<br>Can be customized to preserve or combine any/all dimensions. | |
| All Vendors | Single-average DICOM | Default setting: dimensions are collapsed. Depending on settings, individual transients can be exported in separate DICOM files. | |

#### Considerations

Experts noted that specific customized options have to be set on the exam cards to enable the export of individual transients (2/21, 9.5%), however these options may not be available on all scanners or implementations.

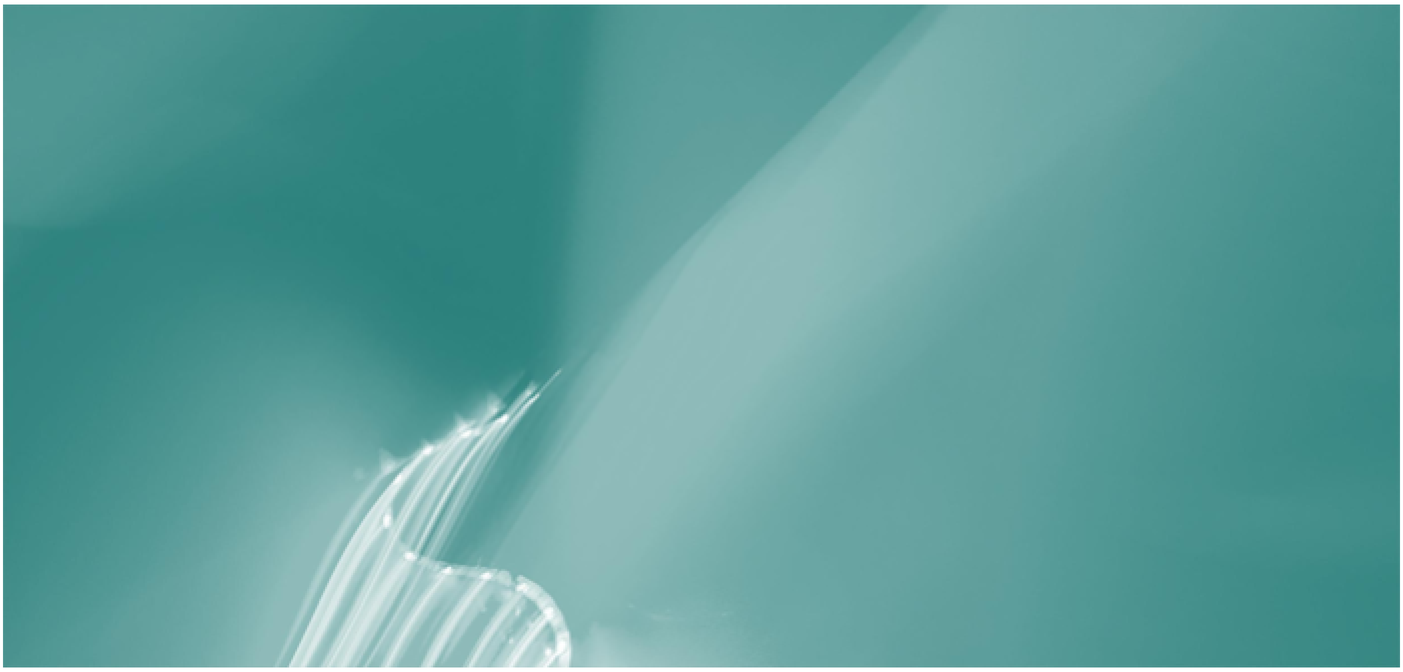

### 5. Quality & Reporting


5.1 Quality Metrics

5.2 Reporting

#### 5.1. Quality Metrics

| Recommendation | ADOPT: Report spectral quality in terms of the signal-to-noise ratio, linewidth, water suppression efficiency, fit quality and the presence of unwanted spectral features | GRADE A<br>Agreement |
|---|---|---|

##### Preface

Due to the inherently low SNR, MRS acquisitions require a high degree of stability from the participant and the equipment. Spectra of low quality will result in less reliable (or wrong) quantification of the metabolite of interest. Judging the quality of an MRS spectrum by visual inspection requires experience. Several quantitative metrics of data quality allow more objective judgement whether the acquisition has been successful in terms of shim quality, water suppression, presence of artefacts, and quality of the data modelling. Another commonly used expression of uncertainty is the Cramér-Rao lower bounds (CRLB). CRLB can be considered as the "maximum trust that can be associated with an area (and thus concentration) estimated in model fitting"[99]. While they can be a useful indicator of quality for quantitative MRS, if used as a percentage of the estimated value (relative CRLB) results can be significantly biased due to the exclusion of potentially clinically meaningful data.[28,99]. An alternative is to use absolute CRLBs [99]. However, no single quality measure alone is sufficient to demonstrate the overall quality of data.

##### Evidence summary

Seven studies provide recommendations on quality metrics (three consensus documents; Level 1[6,28,35] and four methodological publications; Level 3[27,54,99,100]). One consensus document (Level 1)[35] made 7 recommendations on the variables to assess in order to determine spectroscopy quality: (1) SNR, (2) metabolite and unsuppressed water resonance linewidths, (3), residual water signal, (4) line shape, (5) CRLBs of the data fit, (6) fit quality (relative size of residuals versus the standard deviation of noise), and (7) presence of artefacts (spurious signals, baseline distortions, contamination from subcutaneous lipids). Of the four methodological publications (Level 3), 2 discussed CRLBs[99,100], 1 discussed bootstrapping[27], and 1 reported expected values for water linewidth cutoffs[54]. Two studies recommend the use of absolute CRLBs but not relative CRLBs based on the risk of introducing selection bias[35,99] (Level 1 and Level 5). Relative-CRLB cut-off recommendations of between 20-50% have been shown to bias exclusion and potentially obscure clinically meaningful differences in clinical populations[28,99].

##### Considerations

In cases where MRS analysis software packages do not report CRLBs, an alternative metric of fit error is the standard deviation of the fit residual[6].

#### 5.2. Reporting

| Recommendation | ADOPT: When reporting results use one of these two checklists (MRS in MRS- Lin et al. 2020[22]) or the MRS-Q (Peek et al. 2020[2]) using the appropriate terminology (Kreis et al. 2020). Include detailed reporting of hardware, MEGA-PRESS specific acquisition parameters including quantification details, quality and analysis methods. | GRADE A<br>Agreement |
|---|---|---|

##### Preface

Reporting of methods needs to contain sufficient information for readers to replicate the study and the data analysis. The quantitative results of MRS measurements depend strongly on the acquisition parameters, data quality, and the choice of analysis methodology. It is therefore required to report every step of data acquisition, processing, and analysis in as much detail as possible.

##### Evidence summary

Three studies provide recommendations on what should be reported in a MEGA-PRESS GABA study (one consensus document; Level 1[22], one systematic review; Level 2[2] and one methodological publication; Level 3[30]). The consensus document and systematic review were in agreement. The consensus document (Level 1) [22] reported five areas that require reporting; 1) hardware, 2) acquisition, 3) data analysis, 4) methods and 5) outputs and data quality. The systematic review (Level 2)[2] produced an 11-point checklist (MRS-Q) under the broader domains; 1) scanner, sequence parameters, 2) quality measures, 3) sample size calculation, 4) partial volume correction, and 5) analysis. The methodological publication (Level 3)[54] highlighted five aspects of optimization often not reported in edited MRS studies: 1) procedure to calculate and set the frequency of editing pulse; 2) time when editing pulse frequency is set and whether it is updated during acquisition; 3) length and bandwidth of localization pulses; 4) GABA relaxation times used for quantification; 5) homocarnosine co-editing often not mentioned (while MM is).

##### Considerations

When reporting MRS studies, refer to the consensus document on terminology and concepts for characterization[101]

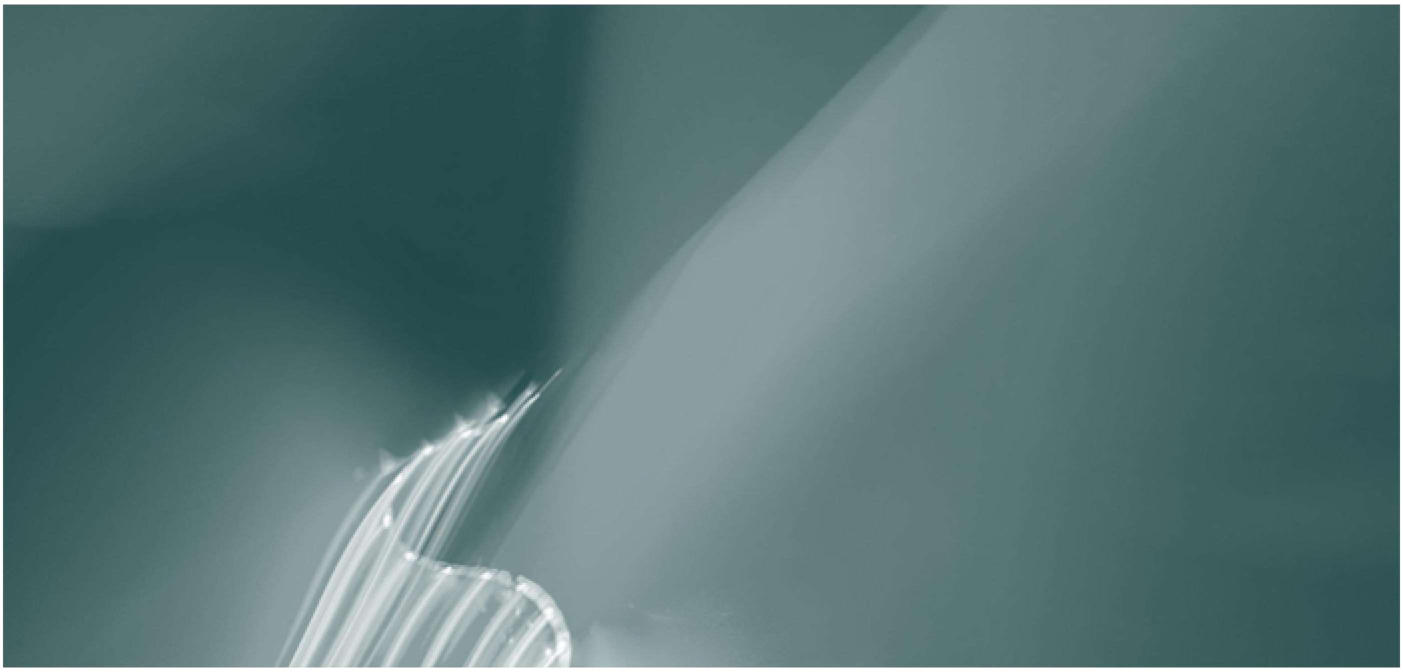

### 6. Post-Processing

6.1 Frequency-and-Phase Correction (Post-processing)

#### 6.1. Frequency-and-Phase Correction (Post-processing)

| Recommendation | ADOPT: Frequency-and-phase alignment of individual transients should be performed during post-processing. | GRADE A<br>Agreement |
|---|---|---|

##### Preface

Frequency-and-phase correction (FPC) is the post-processing step of aligning individual transients of a MEGA-PRESS acquisition and aligning the averaged edit-ON and edit-OFF spectra to each other. FPC techniques have been developed to address the strong susceptibility of MEGA-PRESS to subtraction artefacts, i.e. unwanted artefacts arising from spectral misalignment during the calculation of the GABA-edited difference spectrum. These artefacts can commonly occur in clinical populations due to head position, and significantly reduce the precision of data modelling and quantification.

##### Evidence summary

Ten studies provide recommendations on frequency and phase correction (two consensus documents; Level 1[36,37] and eight methodological publications; Level 3 [8,102-108]). The two consensus documents [36,37] recommended that spectral alignment routines be used during post-processing to improve the quality of the final spectrum for both unedited and edited MRS data. Two of the methodological publications (Level 3)[104,105] found that using the spectral registration algorithm for FPC of individual averages improves the linewidth and SNR of MRS data, and reduces subtraction artefacts in MEGA-PRESS data. One methodological publication (Level 3)[8] concurs that subtraction artefact can be improved in scans showing significant drift, however, editing efficiency and the GABA-to-MM signal ratio cannot be improved with this step alone. Three papers[106-108] demonstrated that appropriate alignment of edit-ON and edit-OFF spectra reduces subtraction artefacts in MEGA-PRESS data, and improved quantification. One methodological publication (Level 3)[103] found that determining the individual frequency history of an acquisition and calculating individual basis sets for linear-combination modelling based on this history, improves modelling accuracy. However, this method is not implemented in any currently available analysis software package.

##### Considerations

Experts highlighted that post-processing cannot compensate retrospectively for the impact of acquiring data with incorrect editing pulse frequency, e.g. as a result of frequency drift (2/21, 9.52%). Therefore, frequency drift needs to be monitored at time of acquisition (see Quality assessment during the scan).

### References

1. Enna SJ, McCarson KE. The Role of GABA in the Mediation and Perception of Pain. In: *Gaba.*2006:1-27.
2. Peek AL, Rebbeck T, Puts NAJ, Watson J, Aguila ME, Leaver AM. Brain GABA and glutamate levels across pain conditions: A systematic literature review and meta-analysis of 1H-MRS studies using the MRS-Q quality assessment tool. *NeuroImage.* 2020:116532.
3. Schür RR, Draisma LW, Wijnen JP, et al. Brain GABA levels across psychiatric disorders: A systematic literature review and meta-analysis of 1H-MRS studies. *Human brain mapping.* 2016;37(9):3337-3352.
4. Godfrey KE, Gardner AC, Kwon S, Chea W, Muthukumaraswamy SD. Differences in excitatory and inhibitory neurotransmitter levels between depressed patients and healthy controls: a systematic review and meta-analysis. *Journal of psychiatric research.* 2018;105:33-44.
5. Marotta R, Risoleo MC, Messina G, et al. The neurochemistry of autism. *Brain sciences.* 2020;10(3):163.
6. Mullins PG, McGonigle DJ, O'Gorman RL, et al. Current practice in the use of MEGA-PRESS spectroscopy for the detection of GABA. *Neuroimage.* 2014;86:43-52.
7. Mescher M, Merkle H, Kirsch J, Garwood M, Gruetter R. Simultaneous in vivo spectral editing and water suppression. *NMR Biomed.* 1998;11(6):266-272.
8. Harris AD, Glaubitz B, Near J, et al. Impact of frequency drift on gamma-aminobutyric acid-edited MR spectroscopy. *Magnetic resonance in medicine.* 2014;72(4):941-948.
9. Peek AL, Rebbeck T, Leaver AM, et al. The Comprehensive Guide to MEGA-PRESS for GABA Measurement using MEGA-PRESS. *TBC.* 2021.
10. NHMRC. Guidelines for Guidelines Handbook. National Health and Medical Research Council www.nhmrc.gov.au/guidelinesforguidelines. Published 2021. Accessed Jan, 2020.
11. The ADAPTE Collaboration. The ADAPTE Process: Resource Toolkit for Guideline Adaptation. http://www.g-i-n.net. Published 2009. Accessed 01/07, 2019.
12. Hasson F, Keeney S, McKenna H. Research guidelines for the Delphi survey technique. *Journal of Advanced Nursing.* 2000;32(4):1008-1015.
13. Miller KA, Collada B, Tolliver D, et al. Using the Modified Delphi Method to Develop a Tool to Assess Pediatric Residents Supervising on Inpatient Rounds. *Academic Pediatrics.* 2020;20(1):89-96.
14. NHMRC. *NHMRC levels of evidence and grades for recommendations for developers of guidelines.* Canberra: NHMRC;2009.
15. Puts NA. *The in vivo functional neuroanatomy and neurochemistry of vibrotactile processing*, Cardiff University; 2011.
16. Mikkelsen M, Barker PB, Bhattacharyya PK, et al. Big GABA: Edited MR spectroscopy at 24 research sites. *Neuroimage.* 2017;159:32-45.
17. Mikkelsen M, Rimbault DL, Barker PB, et al. Big GABA II: Water-referenced edited MR spectroscopy at 25 research sites. *Neuroimage.* 2019;191:537-548.
18. Mikkelsen M, Loo RS, Puts NAJ, Edden RAE, Harris AD. Designing GABA-edited magnetic resonance spectroscopy studies: Considerations of scan duration, signal-to-noise ratio and sample size. *Journal of neuroscience methods.* 2018;303:86-94.
19. Brix MK, Ersland L, Hugdahl K, et al. Within- and between-session reproducibility of GABA measurements with MR spectroscopy. *Journal of magnetic resonance imaging : JMRI.* 2017;46(2):421-430.
20. Sanaei Nezhad F, Anton A, Michou E, Jung J, Parkes LM, Williams SR. Quantification of GABA, glutamate and glutamine in a single measurement at 3 T using GABA-edited MEGA-PRESS. *NMR in biomedicine.* 2018;31(1).
21. Bhattacharyya PK, Lowe MJ, Phillips MD. Spectral quality control in motion-corrupted single-voxel J-difference editing scans: an interleaved navigator approach. *Magnetic resonance in medicine.* 2007;58(4):808-812.
22. Lin Aea. Reporting guidelines for magnetic resonance spectroscopy publications: Experts' consensus recommendations. *NMR Biomed.* 2020.
23. Sanaei Nezhad F, Lea-Carnall CA, Anton A, et al. Number of subjects required in common study designs for functional GABA magnetic resonance spectroscopy in the human brain at 3 Tesla. *European Journal of Neuroscience.* 2019;09:09.
24. de Graaf RA. *In Vivo NMR Spectroscopy: Principles and Techniques.* Wiley; 2019.
25. Bergmann J, Pilatus U, Genc E, Kohler A, Singer W, Pearson J. V1 surface size predicts GABA concentration in medial occipital cortex. *Neuroimage.* 2016;124(Pt A):654-662.

26. Bai X, Edden RA, Gao F, et al. Decreased gamma-aminobutyric acid levels in the parietal region of patients with Alzheimer's disease. *Journal of Magnetic Resonance Imaging.* 2015;41(5):1326-1331.
27. Chen M, Liao C, Chen S, et al. Uncertainty assessment of gamma-aminobutyric acid concentration of different brain regions in individual and group using residual bootstrap analysis. *Medical & biological engineering & computing.* 2017;55(6):1051-1059.
28. Wilson M, Andronesi O, Barker PB, et al. Methodological consensus on clinical proton MRS of the brain: Review and recommendations. *Magnetic resonance in medicine.* 2019;82:527-550.
29. Puts NA, Barker PB, Edden RA. Measuring the longitudinal relaxation time of GABA in vivo at 3 Tesla. *Journal of Magnetic Resonance Imaging.* 2013;37(4):999-1003.
30. Deelchand DK, Marjańska M, Henry P-G, Terpstra M. MEGA-PRESS of GABA+: Influences of acquisition parameters. *NMR in Biomedicine.* 2019;n/a(n/a):e4199.
31. Cudalbu Cea. Contribution of macromolecules to magnetic resonance spectra: Experts' consensus recommendations. *NMR Biomed.* 2020.
32. Edden RAE, Puts NAJ, Barker PB. Macromolecule-suppressed GABA-edited magnetic resonance spectroscopy at 3T. *Magnetic Resonance in Medicine.* 2012;68(3):657-661.
33. Harris AD, Puts NA, Barker PB, Edden RA. Spectral-editing measurements of GABA in the human brain with and without macromolecule suppression. *Magnetic Resonance in Medicine.* 2015;74(6):1523-1529.
34. Edden RA, Oeltzschner G, Harris AD, et al. Prospective frequency correction for macromolecule-suppressed GABA editing at 3T. *Journal of Magnetic Resonance Imaging.* 2016;44(6):1474-1482.
35. Öz G, Deelchand DK, Wijnen JP, et al. Advanced single voxel 1H magnetic resonance spectroscopy techniques in humans: Experts' consensus recommendations. *NMR in biomedicine.* 2020;n/a(n/a):e4236.
36. Choi IY, Andronesi OC, Barker P, et al. Spectral editing in 1 H magnetic resonance spectroscopy: Experts' consensus recommendations. *NMR in Biomedicine.* 2021;34(5).
37. Near J, Harris AD, Juchem C, et al. Preprocessing, analysis and quantification in single-voxel magnetic resonance spectroscopy: experts' consensus recommendations. *NMR in biomedicine.* 2020.
38. Hall EL, Stephenson MC, Price D, Morris PG. Methodology for improved detection of low concentration metabolites in MRS: optimised combination of signals from multi-element coil arrays. *Neuroimage.* 2014;86:35-42.
39. Oeltzschner G, Schnitzler A, Wickrath F, Zollner HJ, Wittsack HJ. Use of quantitative brain water imaging as concentration reference for J-edited MR spectroscopy of GABA. *Magnetic Resonance Imaging.* 2016;34(8):1057-1063.
40. Ernst T, Chang L. Elimination of artifacts in short echo time H MR spectroscopy of the frontal lobe. *Magnetic resonance in medicine.* 1996;36(3):462-468.
41. Dydak U. Practicalities of Acquisition. GABA MRS Editing School; 2018; Mexico.
42. Kreis R. Issues of spectral quality in clinical 1H-magnetic resonance spectroscopy and a gallery of artifacts. *NMR Biomed.* 2004;17(6):361-381.
43. Shungu DC, Mao X, Gonzales R, et al. Brain gamma-aminobutyric acid (GABA) detection in vivo with the J-editing (1) H MRS technique: a comprehensive methodological evaluation of sensitivity enhancement, macromolecule contamination and test-retest reliability. *NMR in Biomedicine.* 2016;29(7):932-942.
44. Mikkelsen M, Singh KD, Sumner P, Evans CJ. Comparison of the repeatability of GABA-edited magnetic resonance spectroscopy with and without macromolecule suppression. *Magnetic Resonance in Medicine.* 2016;75(3):946-953.
45. Keltner JR, Wald LL, Christensen JD, et al. A technique for detecting GABA in the human brain with PRESS localization and optimized refocusing spectral editing radiofrequency pulses. *Magnetic resonance in medicine.* 1996;36(3):458-461.
46. Saleh MG, Rimbault D, Mikkelsen M, et al. Multi-vendor standardized sequence for edited magnetic resonance spectroscopy. *Neuroimage.* 2019;189:425-431.
47. Henry PG, Dautry C, Hantraye P, Bloch G. Brain GABA editing without macromolecule contamination. *Magnetic Resonance in Medicine.* 2001;45(3):517-520.
48. Bai X, Harris AD, Gong T, et al. Voxel Placement Precision for GABA-Edited Magnetic Resonance Spectroscopy. *Open J Radiol.* 2017;7(1):35-44.

49. Park YW, Deelchand DK, Joers JM, et al. AutoVOI: real-time automatic prescription of volume-of-interest for single voxel spectroscopy. *Magnetic resonance in medicine.* 2018;80(5):1787-1798.
50. Juchem C, Cudalbu C, de Graaf RA, et al. B0 shimming for in vivo magnetic resonance spectroscopy: Experts' consensus recommendations. *NMR Biomed.* 2021;34(5):e4350.
51. Juchem C, de Graaf RA. B0 magnetic field homogeneity and shimming for in vivo magnetic resonance spectroscopy. *Anal Biochem.* 2017;529:17-29.
52. Grewal M, Dabas A, Saharan S, Barker PB, Edden RA, Mandal PK. GABA quantitation using MEGA-PRESS: Regional and hemispheric differences. *Journal of magnetic resonance imaging : JMRI.* 2016;44(6):1619-1623.
53. Saleh MG, Near J, Alhamud A, Robertson F, van der Kouwe AJ, Meintjes EM. Reproducibility of macromolecule suppressed GABA measurement using motion and shim navigated MEGA-SPECIAL with LCModel, jMRUI and GANNET. *Magma.* 2016;29(6):863-874.
54. Deelchand DK, Kantarci K, Oz G. Improved localization, spectral quality, and repeatability with advanced MRS methodology in the clinical setting. *Magnetic resonance in medicine.* 2018;79(3):1241-1250.
55. Andronesi OC, Bhattacharyya PK, Bogner W, et al. Motion correction methods for MRS: experts' consensus recommendations. *NMR in Biomedicine.* 2020;n/a(n/a):e4364.
56. Duncan NW, Zhang J, Northoff G, Weng X. Investigating GABA concentrations measured with macromolecule suppressed and unsuppressed MEGA-PRESS MR spectroscopy and their relationship with BOLD responses in the occipital cortex. *Journal of Magnetic Resonance Imaging.* 2019;50(4):1285-1294.
57. Gu M, Hurd R, Noeske R, et al. GABA editing with macromolecule suppression using an improved MEGA-SPECIAL sequence. *Magnetic Resonance in Medicine.* 2018;79(1):41-47.
58. Oeltzschner G, Chan KL, Saleh MG, Mikkelsen M, Puts NA, Edden RAE. Hadamard editing of glutathione and macromolecule-suppressed GABA. *NMR in Biomedicine.* 2018;31(1).
59. Puts NA, Edden RA. In vivo magnetic resonance spectroscopy of GABA: a methodological review. *Prog Nucl Magn Reson Spectrosc.* 2012;60:29-41.
60. Harada M, Kubo H, Nose A, Nishitani H, Matsuda T. Measurement of variation in the human cerebral GABA level by in vivo MEGA-editing proton MR spectroscopy using a clinical 3 T instrument and its dependence on brain region and the female menstrual cycle. *Human Brain Mapping.* 2011;32(5):828-833.
61. Waddell KW, Zanjanipour P, Pradhan S, et al. Anterior cingulate and cerebellar GABA and Glu correlations measured by 1H J-difference spectroscopy. *Magnetic Resonance Imaging.* 2011;29(1):19-24.
62. van der Veen JW, Shen J. Regional difference in GABA levels between medial prefrontal and occipital cortices. *Journal of Magnetic Resonance Imaging.* 2013;38(3):745-750.
63. Harris AD, Puts NAJ, Anderson BA, et al. Multi-Regional Investigation of the Relationship between Functional MRI Blood Oxygenation Level Dependent (BOLD) Activation and GABA Concentration. *PLOS ONE.* 2015;10(2):e0117531.
64. Long Z, Dyke JP, Ma R, Huang CC, Louis ED, Dydak U. Reproducibility and effect of tissue composition on cerebellar gamma-aminobutyric acid (GABA) MRS in an elderly population. *NMR in Biomedicine.* 2015;28(10):1315-1323.
65. Greenhouse I, Noah S, Maddock RJ, Ivry RB. Individual differences in GABA content are reliable but are not uniform across the human cortex. *Neuroimage.* 2016;139:1-7.
66. Puts NAJ, Heba S, Harris AD, et al. GABA Levels in Left and Right Sensorimotor Cortex Correlate across Individuals. *Biomedicines.* 2018;6(3):24.
67. Dhamala E, Abdelkefi I, Nguyen M, Hennessy TJ, Nadeau H, Near J. Validation of in vivo MRS measures of metabolite concentrations in the human brain. *NMR in Biomedicine.* 2019;32(3):e4058.
68. Porges EC, Woods AJ, Edden RA, et al. Frontal Gamma-Aminobutyric Acid Concentrations Are Associated With Cognitive Performance in Older Adults. *Biological psychiatry Cognitive neuroscience and neuroimaging.* 2017;2(1):38-44.
69. Gao F, Edden RA, Li M, et al. Edited magnetic resonance spectroscopy detects an age-related decline in brain GABA levels. *Neuroimage.* 2013;78:75-82.
70. van Veenendaal TM, Backes WH, van Bussel FCG, et al. Glutamate quantification by PRESS or MEGA-PRESS: Validation, repeatability, and concordance. *Magnetic Resonance Imaging.* 2018;48:107-114.

71. Choi IY, Lee SP, Merkle H, Shen J. In vivo detection of gray and white matter differences in GABA concentration in the human brain. *Neuroimage.* 2006;33(1):85-93.
72. Geramita M, van der Veen JW, Barnett AS, et al. Reproducibility of prefrontal γ-aminobutyric acid measurements with J-edited spectroscopy. *NMR in Biomedicine.* 2011;24(9):1089-1098.
73. Bhattacharyya PK, Phillips MD, Stone LA, Lowe MJ. In vivo magnetic resonance spectroscopy measurement of gray-matter and white-matter gamma-aminobutyric acid concentration in sensorimotor cortex using a motion-controlled MEGA point-resolved spectroscopy sequence. *Magnetic Resonance Imaging.* 2011;29(3):374-379.
74. Harris AD, Puts NA, Edden RA. Tissue correction for GABA-edited MRS: Considerations of voxel composition, tissue segmentation, and tissue relaxations. *Journal of Magnetic Resonance Imaging.* 2015;42(5):1431-1440.
75. Mikkelsen M, Singh KD, Brealy JA, Linden DE, Evans CJ. Quantification of gamma-aminobutyric acid (GABA) in 1-H MRS volumes composed heterogeneously of grey and white matter. *NMR in Biomedicine.* 2016;29(11):1644-1655.
76. Porges EC, Woods AJ, Lamb DG, et al. Impact of tissue correction strategy on GABA-edited MRS findings. *Neuroimage.* 2017;162:249-256.
77. Gasparovic C, Chen H, Mullins PG. Errors in 1 H-MRS estimates of brain metabolite concentrations caused by failing to take into account tissue-specific signal relaxation. *NMR in Biomedicine.* 2018;31(6):e3914.
78. Petroff OA, Spencer DD, Alger JR, Prichard JW. High-field proton magnetic resonance spectroscopy of human cerebrum obtained during surgery for epilepsy. *Neurology.* 1989;39(9):1197-1202.
79. Jensen J, Frederick B, Renshaw P. Grey and white matter GABA level differences in the human brain using two-dimensional, J-resolved spectroscopic imaging. *NMR in biomedicine.* 2005;18:570-576.
80. Porges EC, Jensen G, Foster B, Edden RA, Puts NA. The trajectory of cortical GABA across the lifespan, an individual participant data meta-analysis of edited MRS studies. *eLife.* 2021;10.
81. Aufhaus E, Weber-Fahr W, Sack M, et al. Absence of changes in GABA concentrations with age and gender in the human anterior cingulate cortex: a MEGA-PRESS study with symmetric editing pulse frequencies for macromolecule suppression. *Magnetic resonance in medicine.* 2013;69(2):317-320.
82. Maes C, Hermans L, Pauwels L, et al. Age-related differences in GABA levels are driven by bulk tissue changes. *Human Brain Mapping.* 2018;39(9):3652-3662.
83. Marenco S, Meyer C, van der Veen JW, et al. Role of gamma-amino-butyric acid in the dorsal anterior cingulate in age-associated changes in cognition. *Neuropsychopharmacology : official publication of the American College of Neuropsychopharmacology.* 2018;43(11):2285-2291.
84. Simmonite M, Carp J, Foerster BR, et al. Age-Related Declines in Occipital GABA are Associated with Reduced Fluid Processing Ability. *Academic Radiology.* 2019;26(8):1053-1061.
85. Saleh MG, Near J, Alhamud A, Van Der Kouwe AJW, Meintjes EM. Effects of tissue and gender on macromolecule suppressed gamma-aminobutyric acid. *International Journal of Imaging Systems and Technology.* 2017;27(2):144-152.
86. O'Gorman RL, Michels L, Edden RA, Murdoch JB, Martin E. In vivo detection of GABA and glutamate with MEGA-PRESS: reproducibility and gender effects. *Journal of magnetic resonance imaging : JMRI.* 2011;33(5):1262-1267.
87. Bhagwagar Z, Wylezinska M, Taylor M, Jezzard P, Matthews PM, Cowen PJ. Increased brain GABA concentrations following acute administration of a selective serotonin reuptake inhibitor. *The American journal of psychiatry.* 2004;161(2):368-370.
88. Petroff OA, Rothman DL, Behar KL, Collins TL, Mattson RH. Human brain GABA levels rise rapidly after initiation of vigabatrin therapy. *Neurology.* 1996;47(6):1567-1571.
89. Petroff OA, Rothman DL, Behar KL, Lamoureux D, Mattson RH. The effect of gabapentin on brain gamma-aminobutyric acid in patients with epilepsy. *Annals of Neurology.* 1996;39(1):95-99.
90. Rothman DL, Petroff OA, Behar KL, Mattson RH. Localized 1H NMR measurements of gamma-aminobutyric acid in human brain in vivo. *Proceedings of the National Academy of Sciences of the United States of America.* 1993;90(12):5662-5666.
91. Licata SC, Jensen JE, Penetar DM, Prescot AP, Lukas SE, Renshaw PF. A therapeutic dose of zolpidem reduces thalamic GABA in healthy volunteers: a proton MRS study at 4 T. *Psychopharmacology.* 2009;203(4):819-829.
92. Cai K, Nanga RP, Lamprou L, et al. The impact of gabapentin administration on brain GABA and glutamate concentrations: a 7T 1H-MRS study. *Neuropsychopharmacology.* 2012;37(13):2764-2771.

93. Myers JF, Evans CJ, Kalk NJ, Edden RA, Lingford-Hughes AR. Measurement of GABA using J-difference edited 1H-MRS following modulation of synaptic GABA concentration with tiagabine. *Synapse (New York, NY).* 2014;68(8):355-362.
94. Schulte MHJ, Kaag AM, Wiers RW, et al. Prefrontal Glx and GABA concentrations and impulsivity in cigarette smokers and smoking polysubstance users. *Drug and alcohol dependence.* 2017;179:117-123.
95. Epperson CN, O'Malley S, Czarkowski KA, et al. Sex, GABA, and nicotine: the impact of smoking on cortical GABA levels across the menstrual cycle as measured with proton magnetic resonance spectroscopy. *Biological psychiatry.* 2005;57(1):44-48.
96. Oeltzschner G, Zöllner HJ, Jonuscheit M, Lanzman RS, Schnitzler A, Wittsack H-J. J-difference-edited MRS measures of γ-aminobutyric acid before and after acute caffeine administration. *Magnetic resonance in medicine.* 2018.
97. Epperson CN, Haga K, Mason GF, et al. Cortical gamma-aminobutyric acid levels across the menstrual cycle in healthy women and those with premenstrual dysphoric disorder: a proton magnetic resonance spectroscopy study. *Archives of general psychiatry.* 2002;59(9):851-858.
98. De Bondt T, De Belder F, Vanhevel F, Jacquemyn Y, Parizel PM. Prefrontal GABA concentration changes in women-Influence of menstrual cycle phase, hormonal contraceptive use, and correlation with premenstrual symptoms. *Brain research.* 2015;1597:129-138.
99. Kreis R. The trouble with quality filtering based on relative Cramér-Rao lower bounds. *Magnetic resonance in medicine.* 2016;75(1):15-18.
100. Bolliger CS, Boesch C, Kreis R. On the use of Cramer-Rao minimum variance bounds for the design of magnetic resonance spectroscopy experiments. *NeuroImage.* 2013;83:1031-1040.
101. Kreis R, Boer V, Choi I-Y, et al. Terminology and concepts for the characterization of in vivo MR spectroscopy methods and MR spectra: Background and experts' consensus recommendations. *NMR in biomedicine.* 2021;34(5):e4347.
102. Edden RA, Barker PB. Spatial effects in the detection of gamma-aminobutyric acid: improved sensitivity at high fields using inner volume saturation. *Magnetic resonance in medicine.* 2007;58(6):1276-1282.
103. van der Veen JW, Marenco S, Berman KF, Shen J. Retrospective correction of frequency drift in spectral editing: The GABA editing example. *NMR in Biomedicine.* 2017;30(8).
104. Edden RA, Puts NA, Harris AD, Barker PB, Evans CJ. Gannet: A batch-processing tool for the quantitative analysis of gamma-aminobutyric acid-edited MR spectroscopy spectra. *Journal of magnetic resonance imaging : JMRI.* 2014;40(6):1445-1452.
105. Near J, Edden R, Evans CJ, Paquin R, Harris A, Jezzard P. Frequency and phase drift correction of magnetic resonance spectroscopy data by spectral registration in the time domain. *Magnetic resonance in medicine.* 2015;73(1):44-50.
106. Cleve M, Gussew A, Wagner G, Bar KJ, Reichenbach JR. Assessment of intra- and inter-regional interrelations between GABA+, Glx and BOLD during pain perception in the human brain - A combined (1)H fMRS and fMRI study. *Neuroscience.* 2017;365:125-136.
107. Wiegers EC, Philips BWJ, Heerschap A, van der Graaf M. Automatic frequency and phase alignment of in vivo J-difference-edited MR spectra by frequency domain correlation. *Magma.* 2017;30(6):537-544.
108. Tapper S, Tisell A, Helms G, Lundberg P. Retrospective artifact elimination in MEGA-PRESS using a correlation approach. *Magnetic resonance in medicine.* 2019;81(4):2223-2237.

### Supplementary Material

#### Supplement 1: Level of evidence modified from NHMRC (1999, 2009)

| | MODIFIED EVIDENCE HIERARCHY | | ORIGINAL EVIDENCE HIERARCHY | |
|---|---|---|---|---|
| **Level** | **Design** | **Justification** | **Design** | **Justification** |
| **1** | Consensus Document | Traditionally a systematic review of the most appropriate study design is considered Level 1 evidence. In this case we consider expert consensus documents as Level 1 because akin to systematic reviews in other fields, these consensus documents draw on the most appropriate study designs to inform the parameters required to run a MEGA-PRESS study. All consensus documents included within this review had a panel of authors from multiple institutions across multiple countries. They also benefit from recency, with 7/9 included consensus being published in 2020/2021. | Systematic review | In line with the NHMRC recommendations (NHMRC, 2009) a systematic review of Level 2 studies will be considered Level 1. In this case meta-analysis of the studies will likely improve precision of the results. In cases where systematic reviews are of lower levels of evidence, they will be considered the same level as the studies they include, as they may increase the chance of bias (NHMRC, 2009). |
| | Seminal texts | Where core principles of physics are required to inform the guideline, seminal text of these fundamental physical properties are also considered the highest level of text. | | |
| **2** | Systematic Review | Systematic reviews are considered Level 2 evidence as they pool together results from methodological publications which have been specifically designed to test parameters required to run a MEGA-PRESS study. However, the methodological publications typically have small sample sizes, and limitations and suffer from publication bias. | Randomised Control Trial | In order to investigate the impact of a confounder a randomized control trial would be considered the best design to address the research question. |
| | Large multi-site studies | Large multi-site studies provide the most information on applying parameters in a clinical context; however, the purpose of such trials is rarely to investigate a single parameter required to run a MEGA-PRESS study. | | |
| **3** | Methodological publications | For the purpose of this study, methodological publications were considered as any study that had a specific aim to investigate a parameter required to run a MEGA-PRESS study. These might include studies on humans, phantoms, or simulations. These did not include animal studies. These methodological publications will often test a specific parameter required to run a MEGA-PRESS study and directly inform this guideline. However, these studies are typically performed using small samples, and are often tested on healthy subjects in non-clinical environments. | i) Comparative study with concurrent controls<br><br>ii) Comparative study without concurrent controls | Studies designs that investigate a condition compared to a control group, or situation are considered Level 3 evidence, as they have the potential for bias. |
| | | | Consensus document | Consensus documents are considered Level 3 when investigating confounders, as these research questions are better answered using a scientifically rigorous design, and therefore a consensus document is potentially biased. |
| **4** | Narrative Reviews | Narrative reviews are commonly published in the field of 1H-MRS spectroscopy but must be interpreted with caution due to the high risk of bias and personal opinion. | Case series | Case series are considered Level 4 due to being underpowered to answer these research questions, with no control for comparison. |
